# Neuropsychiatric Disturbances in Mild Cognitive Impairment: A Scientometric Analysis

**DOI:** 10.1101/2023.08.26.23294661

**Authors:** Arisara Amrapala, Michel Sabé, Marco Solmi, Michael Maes

**Affiliations:** Department of Psychiatry, Faculty of Medicine, Chulalongkorn University, Bangkok, Thailand; (A.A.); (M.M.); Cognitive Fitness and Biopsychiatry Technology Research Unit, Faculty of Medicine, Chulalongkorn University, Bangkok, Thailand; Center of Excellence in Digital and AI for Mental Health, Faculty of Engineering, Chulalongkorn University, Bangkok, Thailand; Division of Adult Psychiatry, Department of Psychiatry, University Hospitals of Geneva, Thonex, Switzerland; (M.S.); Department of Psychiatry, University of Ottawa, Ottawa, Ontario, Canada; (M.S.); Department of Mental Health, The Ottawa Hospital, Ottawa, Ontario, Canada; Ottawa Hospital Research Institute (OHRI) Clinical Epidemiology Program University of Ottawa, Ottawa, Ontario, Canada; Department of Child and Adolescent Psychiatry, Charite Universitatsmedizin, Berlin, Germany; Cognitive Impairment and Dementia Research Unit, Faculty of Medicine, Chulalongkorn University, Bangkok, Thailand; Department of Psychiatry, Medical University of Plovdiv and Technological Center for Emergency Medicine, Plovdiv, Bulgaria; Kyung Hee University, Seoul, Republic of Korea; Mental Health Center, University of Electronic Science and Technology of China, Chengdu, China; Research Institute, Medical University of Plovdiv, Plovdiv, Bulgaria

**Keywords:** Neuropsychiatric symptoms, Mild Cognitive Impairment, Scientometrics, Evidence synthesis, CiteSpace

## Abstract

Behavioral and psychological symptoms of dementia (BPSD) have been extensively studied in dementia than its prodromal stage, known as mild cognitive impairment (MCI). A scientometric study on BPSD in MCI would be valuable in synthesizing the existing body of research and provide insights into the trends, networks, and influencers within this area. We searched for related literature in the Web of Science database and extracted complete text and citation records of each publication. The primary objective was to map the research evolution of BPSD in MCI and highlight dominant research themes. The secondary objective was to identify research network characteristics (authors, journals, countries, and institutions) and abundances. A total of 12,369 studies published between 1980 to 2022 were included in the analysis. We found 51 distinct clusters from the co-cited reference network that were highly credible with significant modularity (Q = 0.856) and silhouette scores (S = 0.932). Five major research domains were identified: symptoms, diagnosis, brain substrates, biochemical pathways, and interventions. Within recent years, the research focus in this area is on gut microbiota, e-health, COVID-19, cognition, and delirium. Collectively, findings from this scientometric analysis can help clarify the scope and direction of future research and clinical practices.

## 1. Introduction

Emotional, behavioral, and perceptual disturbances are often present in those suffering from dementia. These disturbances can emerge at any point across the entire disease course - from prodromal stages, including the mild cognitive impairment (MCI) phase, all the way to severe dementia (Savva et al., 2009). These neuropsychiatric symptoms were first collectively described by the International Psychogeriatric Association in 1996 as ‘behavioral and psychological symptoms of dementia’ (BPSD) (Finkel et al., 1997).

Generally, BPSD can be classified into five domains: (1) emotional symptoms - depression/dysphoria, anxiety, euphoria, apathy/indifference, and irritability/lability; (2) cognitive symptoms - delusions and hallucinations; (3) verbal symptoms - agitation/aggression and disinhibition; (4) motor symptoms - aberrant motor activity/wandering; and (5) vegetative symptoms - night-time behavioral disturbances and appetite/eating abnormalities (Cloak and Al Khalili, 2022). BPSD are prevalent in up to 99.0% of dementia patients, with depression, agitation, apathy, anxiety, and irritability being the most common symptoms. Notably, half of these patients were observed to suffer from at least four BPSD at once (Frisoni et al., 1999). For those with MCI, an estimated 12.8% to 66.0% of individuals exhibit some type of BPSD, with the most frequent symptoms being depression (median prevalence of 29.8%), sleep disturbances (median prevalence of 18.3%), and apathy (median prevalence of 15.2%) (Köhler et al., 2016). BPSD can cause significant distress to both the affected individual and their caregivers (Pinyopornpanish et al., 2022), making it essential to identify the individual’s neuropsychiatric symptoms to ensure better prognosis, prevention, and management of their dementia.

Presently, BPSD have gained more worldwide recognition to the extent that starting from the Fifth Edition of the Diagnostic and Statistical Manual of Mental Disorders (DSM-5) onwards, clinicians are now required to determine the type and severity of behavioral and psychological disturbances in addition to the routine neurocognitive diagnosis. Despite these advancements, our understanding of the association between BPSD and MCI remains vague.

Throughout the years, countless comprehensive systematic reviews, meta-analyses, and guidelines have provided extensive knowledge on behavioral and psychological symptoms in various contexts. Among these, tens of thousands of research papers have explored BPSD-related elements in mild cognitive impairment. Given the large volume of literature available on the topic, it is necessary to employ new methods to identify underlying relationships and trends while omitting as little domain-related publications as possible. Accordingly, big data visualization, text mining, and network analysis techniques are key for achieving such a goal. More recently, a new approach by Nakagawa et al. (2019) called ‘research weaving’ was proposed, which enables the research synthesis of both evidence and influence by combining (1) systematic mapping and (2) bibliometric analysis methods. The former method provides a visual overview of the current state of knowledge in the field of interest and its evolution from the past (James et al., 2016), whilst the latter measures the interconnections between scientific evidence and the impact of authors within the field (Zupic and Čater, 2015). Together, this combinatorial technique known as ‘scientometric analysis’ provides investigators and clinicians with an exhaustive summary of a specific research field, its history and dynamics throughout time, and the different levels of networks formed. More importantly, gaps, trends, abundancies, limitations, and factors that potentially influence or cause bias in the research area can also be detected. From these in-depth results, investigators and clinicians can further make better informed decisions in implementing future research studies and health care services.

The scientometric approach is suitable within the field of neuropsychiatry (Sabe et al., 2022), especially for BPSD in MCI due to their multidimensional and dynamic nature, giving rise to a broad range of research spanning from studies on each neuropsychiatric symptom (Lövheim et al., 2008; McKeith and Cummings, 2005), to intervention studies (Kandiah et al., 2019; Orgeta et al., 2015), as well as association studies with other factors such as COVID-19 (Kuroda et al., 2022b) and malnutrition (Kimura et al., 2019). Despite the available systematic reviews, meta-analyses, and bibliometric analyses in this area, there lacks an exhaustive, large-scale analysis of the overlying trends and influences in this topic.

Accordingly, we conducted a scientometric analysis on the BPSD in MCI individuals by combining systematic mapping and bibliometric analysis techniques to fill this knowledge gap. To the best of our knowledge, we are the first to implement such a study.

Our main objective was to produce a systematic map of the evolution of research on BPSD in MCI as well as produce co-cited reference networks and co-occurring keywords networks to highlight the dominant themes in the field throughout time. Our secondary objective was to identify: (1) research network characteristics by looking at measures such as authors, journals, countries, and institutions, and (2) research abundances.

## 2. Methods

### 2.1 Database search and data collection

We conducted an advance search in the Web of Science Core Collection (WOSCC) database using search terms that combined both Medical Subject Headings (MeSH) and keywords (e.g., ‘mild cognitive*’ and ‘behavioral and psychological symptoms of dementia’). The full search terms used can be found in the **Electronic Supplementary File**. Publications were not limited to language and time, however, only those that were classified as articles or reviews were included in the study and the sources were limited to Science Citation Index Expanded.

After the search, the full records until the present search time (July 2022) with the abstract and complete cited references of each paper were extracted from the WOSCC database into tab-delimited plain text files for analysis. Duplicates were eliminated with CiteSpace. Further screening was carried out using Python to eliminate additional publications that were outside the scope of the study. The study flowchart can be found in **Supplementary Figure 1** and the inclusion and exclusion criteria can be found in the **Supplementary Information**.

### 2.2 Data analysis

CiteSpace (6.1.R3) (Chen, 2006), Bibliometrix R packages (3.1.4) (Aria and Cuccurullo, 2017), and VOSviewer (1.6.16) (van Eck and Waltman, 2010) were used to conduct the analyses. The author, institution, country, journal, reference, and keyword information were the units of measurement.

Bibliometric measures in this study were the number of citations, co-citations, and co-occurrences (Boyack and Klavans, 2010). Co-citation refers to the frequency with which two publications are cited together by other articles published afterwards (Small, 1973), whilst co-occurrence is defined by how frequently variables occur together (Zhou et al., 2022). Systematic mapping outcomes were both the directed (direct citation networks) and undirected graphs (co-citation and co-occurrence networks) produced and the co-citation/co-occurrence clusters. The cluster labels were enhanced and interpreted using the automatic cluster labeling and summarization feature provided by CiteSpace (Chen et al., 2010).

Metrics of significance were calculated by CiteSpace, including structural metrics (e.g., betweenness centrality, silhouette score, and modularity), temporal metrics (e.g., citation burstness), and the sigma metric, which is a combination of both structural and temporal matrices.

A node is considered one variable, such as one publication, one author, one country, or one institution.

Betweenness centrality is a measure that captures the extent to which a node lies on the shortest path between other nodes (Hansen et al., 2020). The higher the betweenness centrality, the higher connectivity between different clusters therefore implying that it is a major hub.

Silhouette (S score) is a method to evaluate the performance of the clustering algorithm by measuring the similarity of a sample to its own cluster compared to other clusters. The S score gives a value between -1 to +1, with values closer to 1 indicating that the sample is more compact within the cluster it currently belongs to and further apart from other clusters (i.e., greatly separated clusters). Values closer to 0 imply overlapping clusters while negative values mean that the samples may have been assigned to the incorrect cluster (Belyadi and Haghighat, 2021; Rousseeuw, 1987).

Modularity (Q score) is a measure of the strength of which a network has been divided into clusters or modules using data clustering or community detection methods (Fornito et al., 2016). High modularity networks have denser connections between nodes within the same cluster but sparse connections between nodes in other clusters. The Q score ranges between 0 to 1, with values closer to 1 indicating a strong community structure and a value of 0 implying a community division that is not better than random.

Sigma is a measure combining betweenness centrality and citation burstness through the formula (centrality + 1)^burstness^ (Chen et al., 2010) whereby higher values imply research that has higher influential potential. Citation burstness refers to the extent to which the individual publication is associated with a surge of citations.

When analyzing citation burstnesss, the citations referring to sources that define and classify diseases and disorders (e.g., ICD-10, DSM, etc.) were excluded. Redundant nodes (e.g. Smith J and Smith JA) were merged together where deemed appropriate. Journal impact factors were obtained from the 2022 Journal Citation Reports by extracting the data from WOSCC in plain text files.

Cluster labels were generated from the keyword lists of cited articles within each cluster using the likelihood ratio statistic (p < 0.001). The keyword with the highest association for each cluster is automatically chosen as the cluster name by the software. The cluster names were re-labelled by the authors according to their expertise where it was deemed appropriate.

Information on authors and journals were retrieved using the Bibliometrix R package, whilst network maps of most-cited journals and the co-occurring author keywords network were obtained using VOSviewer. Co-citation analysis (co-cited reference networks and author clusters), co-occurrence analysis (co-occurring author keywords network), and collaboration networks (between countries and institutions) were computed using CiteSpace. All calculations used the g-index, a metric representing an author’s citation score that is more sensitive than the h-index as it gives more weight to highly cited articles (higher g scores indicate higher author citations) (Egghe, 2006). CiteSpace was also used to conduct burst analyses for all units of measure and the parameters can be found in the **Supplementary Information**. Scale factor k = 25 was used for all analyses and timeframe reduction was achieved by CiteSpace via the removal of empty time intervals which optimizes time slicing.

## 3. Results

### 3.1 Co-cited reference networks

A map of co-cited references was generated for the timeframe 1980 to 2022 (**Figure 1)**. The first identified article related to BPSD in MCI was a German article published by Plaum (1980) on cognitive disorders in endogenous depression and manic psychosis.

**Figure 1.**
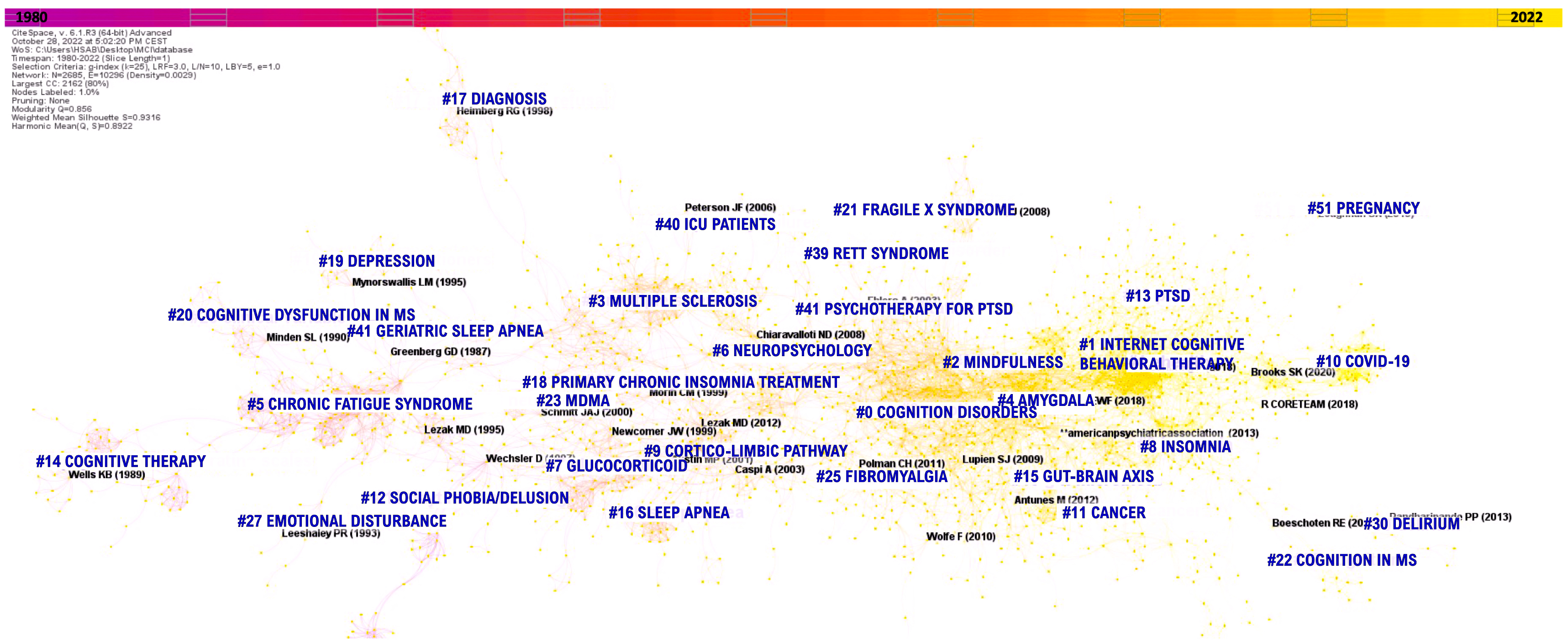
Co-citation reference network between the year 1980 to 2022. This network displays the computed clusters and shows the burstness of hotspots. Each node in this analysis corresponds to one publication and the node size is proportional to the co­citation number of that publication. Burstness is depicted through red tree rings.

From this analysis, 51 distinct clusters were found with significant modularity (Q = 0.856) and silhouette scores (S = 0.932), implying that the resulting clusters are highly credible. We report below the cluster mean year of co-citation, size, and S score in the following format (year;size;score). Cluster 0 (2011;246;0.888) is the largest cluster that has ‘emotion’, ‘fmri’, ‘prefrontal cortex’, ‘insomnia’, and ‘attention bias’ as the top keywords. This cluster was renamed to ‘Cognitive Disorders’. The second largest, Cluster 1 (2018;241;0.889) had the keywords ‘ehealth’, ‘telemedicine’, ‘mhealth’, ‘mental health’ and ‘e-mental health’. This cluster was renamed to ‘internet-derived cognitive behavioral therapy’ (ICBT). Cluster 2 (2011;167;0.867) was the third largest and had ‘internet’, ‘depression’, ‘cognitive therapy’, ‘mindfulness’, and ‘mindfulness-based cognitive therapy’ as the keywords. This cluster was renamed ‘Mindfulness’. Further information regarding all clusters can be found in **Supplementary Figure 2** and **Supplementary Table 1**.

Looking at the evolution of the network (from left to right), the research on BPSD in MCI gained traction in the 1990s with general studies on psychometric tests, which evolved into research in chronic fatigue syndrome, clinical variables, and animal models. In the early 2000s, the research focus shifted to glucocorticoids, sleep apnea, insomnia, multiple sclerosis, and chemotherapy. Later, there was the emergence of research in emotion, internet, gut microbiota, electronic health (e-health), and insomnia in the field, while more recently from 2019 onwards, COVID-19, cognition, systematic review, and delirium became the main research area.

To further examine newer trends of BPSD and MCI research, we generated another map of co-cited references from 2020 to 2022 along with its corresponding clusters (**Figure 2**) and extracted the top 5 most co-cited references (**Table 1**). The highest co-cited publication was a systematic review and meta-analysis by Carlbring et al. (2018) in *Cognitive Behaviour Therapy* with 42 co-citations in our network and 1,031 citations in the literature. From the co-citation network map, 19 highly credible major clusters were retrieved with significant modularity (Q = 0.887) and silhouette scores (S = 0.937). Detailed information regarding each cluster can be found in **Supplementary Table 1**. Results indicate that the biggest focus at present is on self-management, telemedicine, COVID-19, cognitive behavioral therapy, and e-health (Cluster 0; mean year = 2020; size = 123; S = 0.842), as they are the top keywords associated with the cluster. This is the central cluster where a majority of other clusters share hotspots with (both directly and indirectly).

**Table 1.** The 5 most co-cited references between 2020-2022.

| Number of Network Citations | Number of Literature Citations | Cited Reference | Year | Source | Vol | Page | Title | DOI | Type of Paper | Cluster Number (in Fig 2) |
| --- | --- | --- | --- | --- | --- | --- | --- | --- | --- | --- |
| 41 | 1,031 | Carlbring P | 2018 | Cogn Behav Ther | 47 | 1-18 | Internet-based vs. face-to-face cognitive behavior therapy for psychiatric and somatic disorders: an updated systematic review and meta-analysis | 10.1080/16506073.2017.1401115 | Systematic Review and Meta-analysis | 0 |
| 28 | 772 | Andrews G | 2018 | J Anxiety Disord | 55 | 70-78 | Computer therapy for the anxiety and depression disorders is effective, acceptable and practical health care: An updated meta-analysis | 10.1016/j.janxdis.2018.01.001 | Meta-analysis | 0 |
| 20 | 1,573 | Riemann D | 2017 | J Sleep Res | 26 | 675-700 | European guideline for the diagnosis and treatment of insomnia | 10.1111/jsr.12594 | Practice Guideline | 3 |
| 20 | 1,435 | Qaseem A | 2016 | Ann Intern Med | 165 | 125-133 | Management of Chronic Insomnia Disorder in Adults: A Clinical Practice Guideline From the American College of Physicians | 10.7326/M15-2175 | Practice Guideline | 3 |
| 18 | 5,065 | James SL | 2018 | Lancet | 392 | 1789-1858 | Global, regional, and national incidence, prevalence, and years lived with disability for 354 diseases and injuries for 195 countries and territories, 1990–2017: a systematic analysis for the Global Burden of Disease Study 2017 | 10.1016/s0140-6736(18)32279-7 | Systematic Review | 2 |

**Figure 2.**
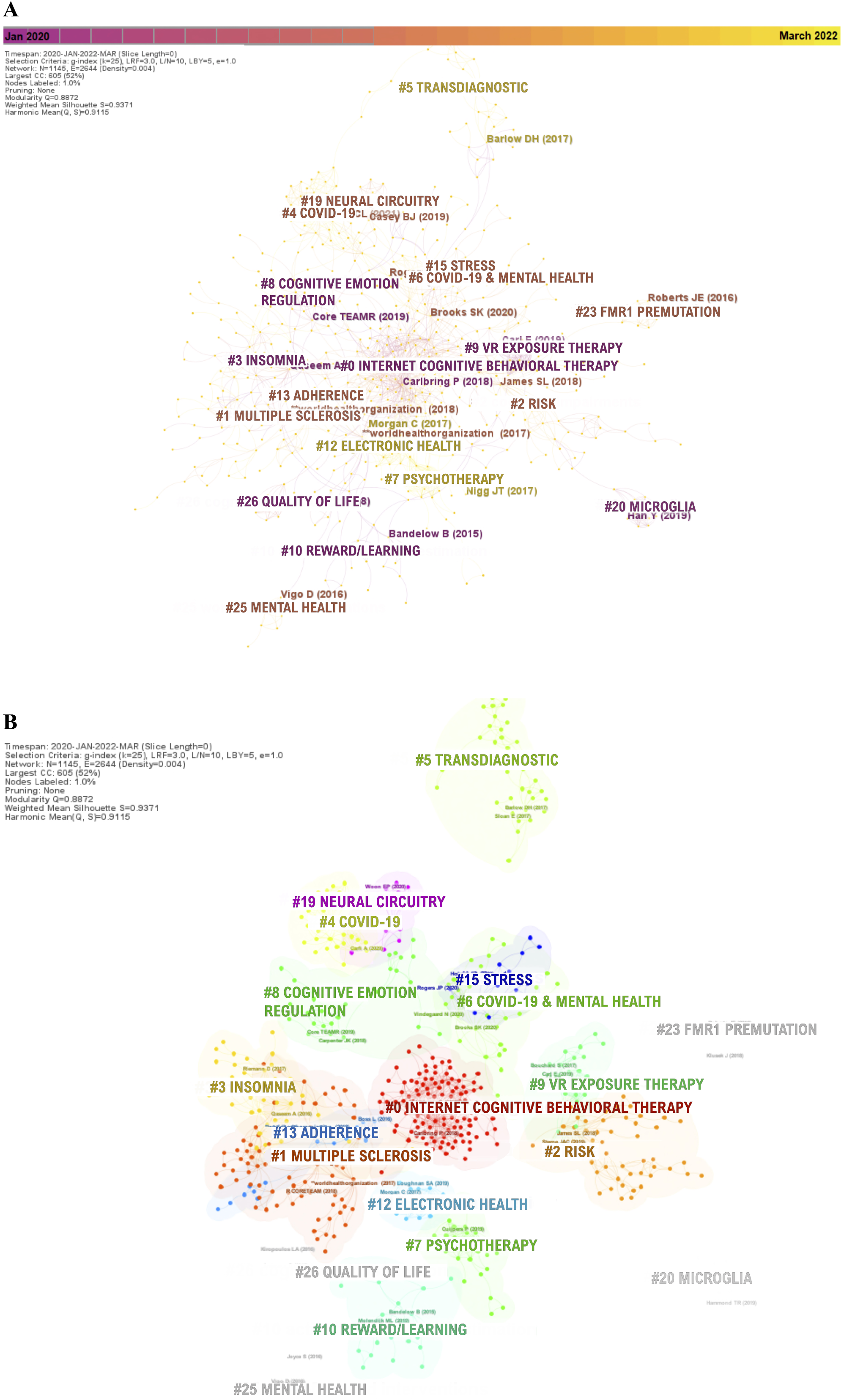
Co-citation reference network between the year 2020 to 2022 (A) and its corresponding clusters (B). This network displays the computed clusters and shows the burstness of hotspots. Each node in this analysis corresponds to one publication and the node size is proportional to the co-citation number of that publication. Citation burstness is depicted through red tree rings.

Broadly considering the co-cited reference networks, the observable major research trends in the field of BPSD in MCI can be grouped into five domains: 1) Symptoms - broad range of symptoms that are related to or make up BPSD, such as depression and anxiety; 2) Diagnosis - associated comorbidities and clinical conditions, such as multiple sclerosis, COVID-19, and fibromyalgia; 3) Brain Substrates - neurological pathways and brain regions related to mental processing, such as the amygdala and hippocampus; 4) Biochemical Pathways - involved metabolic pathways, such as the gut microbiota and glucocorticoids; and 5) Interventions - related pharmaceutical and non-pharmaceutical treatments, such as cognitive therapy and monoamine oxidase inhibitors (MAOIs).

### 3.2 Co-occurring author keywords network

The most used keywords between 2017 to 2022 were analyzed to find recent research trends and hotspots. The findings from this analysis are displayed as a timeline of the co-occurring author keywords network (**Figure 3**).

**Figure 3.**
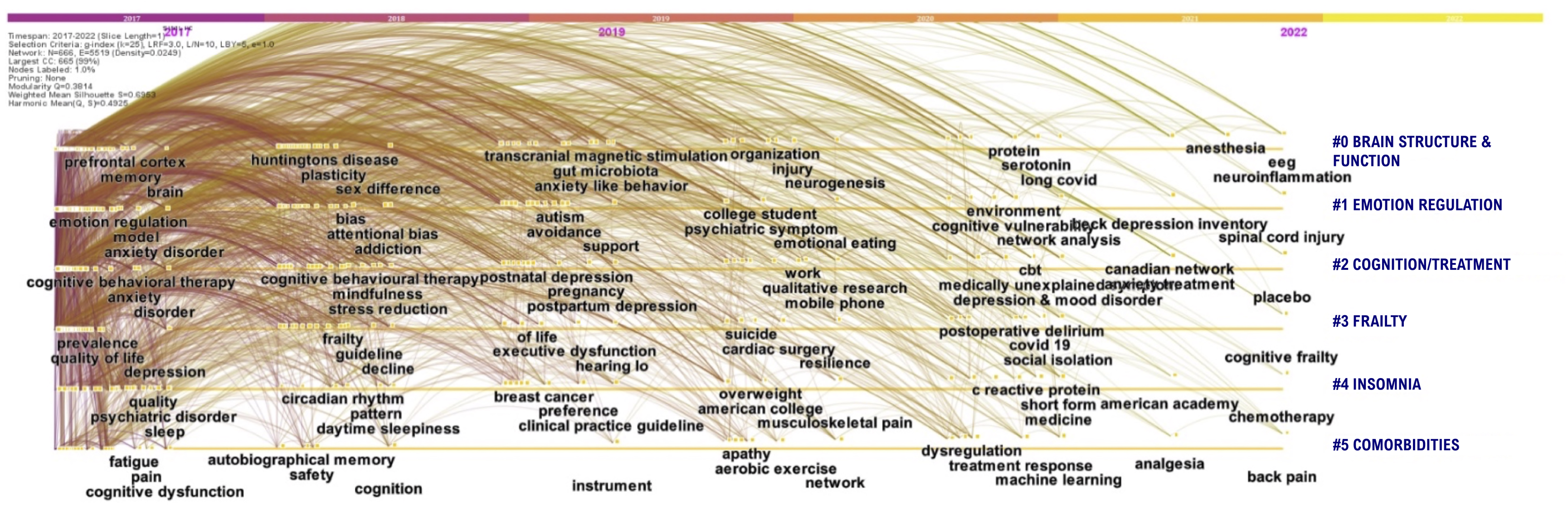
Timeline of co-occurring author keywords networks between the year 2017 and 2022. Each node in this analysis corresponds to one keyword and the timeline at the top is the average publication year of each node. The size of the dot is proportional to the burstness of keyword co-occurrence. This network is generated based on the total link strength across different keyword nodes and the average publication years. The blue labels on the right are the clusters labels.

Six significant clusters of keywords were extracted. The ‘brain structure and function’ keywords cluster was the most important one, followed by ‘emotion regulation’, ‘cognition/treatment’, ‘frailty’, ‘insomnia’, and ‘comorbidities’. The network had an acceptable modularity score (Q > 0.3) and significant silhouette score (S > 0.6).

According to the citation counts, the top five keywords were ‘depression’ (1,215 counts), ‘disorder’ (955), ‘anxiety’ (699), ‘cognitive behavioral therapy (667), and ‘quality of life’ (486). Analysis of burstness found that the three most cited keywords (by the beginning year of citation bursts - 2017) were ‘randomized controlled trial’, ‘attention deficit/hyperactivity disorder’, and ‘event related potential’. When ranked by burst strength, the top three keywords were ‘randomized controlled trial’ with the largest strength of 6.11, followed by ‘functional connectivity’ (5.02) and ‘attention deficit/hyperactivity disorder’ (4.97).

### 3.3 Publication and Journal Characteristics

Our original dataset contained 22,417 publications from the initial search. Through the filtering and selection process (see protocol details in the **Supplementary Information**), this number was further reduced to a final dataset consisting of 12,369 publications.

Of the publications included in this study, 10,320 were articles, 1,913 were reviews, and 165 were editorial materials produced between the year 1980 to 2022. The research was written in 20 different languages with the majority being in English (n = 11,828), followed by German (n = 189) and French (n = 151).

These works were published in 2,084 unique journals and the top 10 journals with the most literature relating to BPSD in MCI can be found in **Table 2**. The *Journal of Affective Disorders* (Publisher: Elsevier; Year Established: 1979) had the highest number of literature with 290 publications, followed by *PLOS One* (Publisher: PLOS; Year Established: 2006) with 224 publications, and *Frontiers in Psychiatry* (Publisher: Frontiers; Year Established: 2010) with 159 publications.

**Table 2.** The top 10 journals with the most articles on BPSD in MCI (1980-2022).

| Journal | Impact Factor<br>(2022) | Number of Articles | Percentage of Articles<br>(%) |
| --- | --- | --- | --- |
| Journal of Affective Disorders | 6.6 | 290 | 2.3 |
| PLOS One | 3.7 | 224 | 1.8 |
| Frontiers in Psychiatry | 4.7 | 159 | 1.3 |
| Psychological Medicine | 6.9 | 136 | 1.1 |
| BMC Psychiatry | 4.4 | 113 | 0.9 |
| Psychiatry Research | 11.3 | 113 | 0.9 |
| Journal of Psychosomatic Research | 4.7 | 112 | 0.9 |
| BMJ Open | 2.9 | 97 | 0.8 |
| Journal of Medical Internet Research | 7.4 | 95 | 0.8 |
| Trials | 2.5 | 94 | 0.8 |

### 3.4 Co-cited author clusters

Collaborative networks between authors (i.e., those who co-authored publications) who conducted research on BPSD in MCI between 2000 to 2022 was generated using information on co-authorship frequency (**Figure 4**).

**Figure 4.**
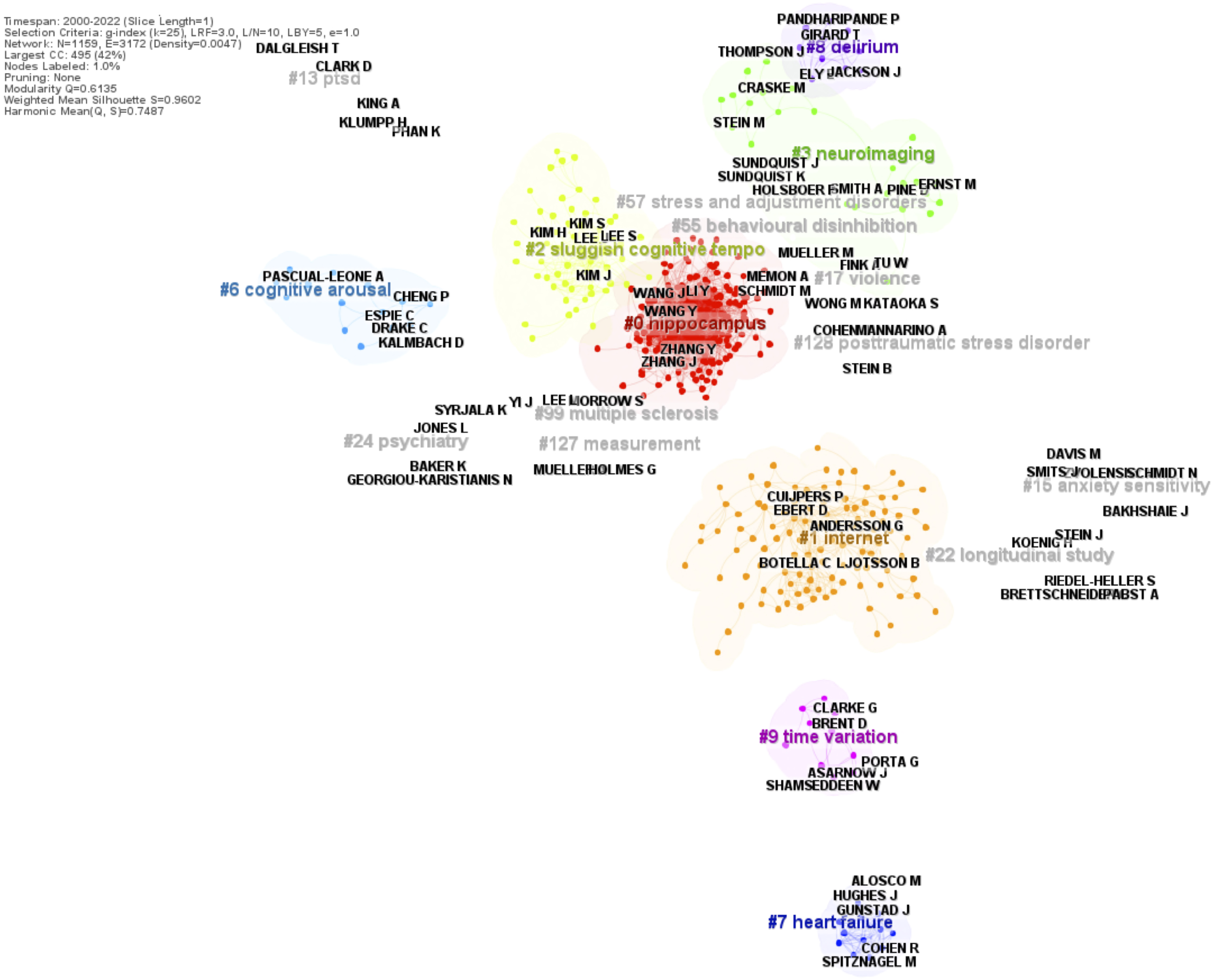
Clusters of the co-authorship network between the year 2000 to 2022. The clusters illustrate research collaborations between authors as calculated by co-authorship frequency. Each node in this analysis corresponds to one author and co-authorship is represented by the links between authors.

The resulting network had highly credible clusters with a high modularity (Q = 0.614) and silhouette score (S = 0.960). Eight major clusters were identified from the network, namely ‘hippocampus’ (cluster size = 213), ‘internet’ (108), ‘sluggish cognitive tempo’ (60), ‘neuroimaging’ (26), ‘cognitive arousal’ (14), ‘heart failure’ (12), ‘delirium’ (11), and ‘time variation’ (10). In the past 22 years, the top five co-authors with the strongest citation burst as ranked by the starting year of bursts were Hickie I (burst strength = 4.65; duration = 2000 - 2015), Arnett P (6.85;2001-2009), Ancoli-Israel S (4.46;2001-2009), Benedict R (7.41;2002 -2015), and Bleijenberg G (5.33;2002-2012); the top five co-authors with the highest citation counts were Zhang Y (126 counts), Wang Y (124), Zhang J (94), Li Y (93), and Wang J (93); the top five co-authors with the strongest citation bursts were Hedman E (burst strength = 8.41), Rapee R (7.46), Benedict R (7.41), Lindeforns N (7.08), and Arnett P (6.85); and the top five co-authors with the highest sigma scores were Cuijpers P (sigma = 1.59), Hollon S (1.14), Andersson G (1.14), Wang L (1.11), and Hughes J (1.10).

### 3.5 Co-cited institutions and countries

Collaborative networks between institutions and countries were generated for the years 2000 to 2022 using information from co-authorship institution frequency and can be seen in **Figure 5**.

**Figure 5.**
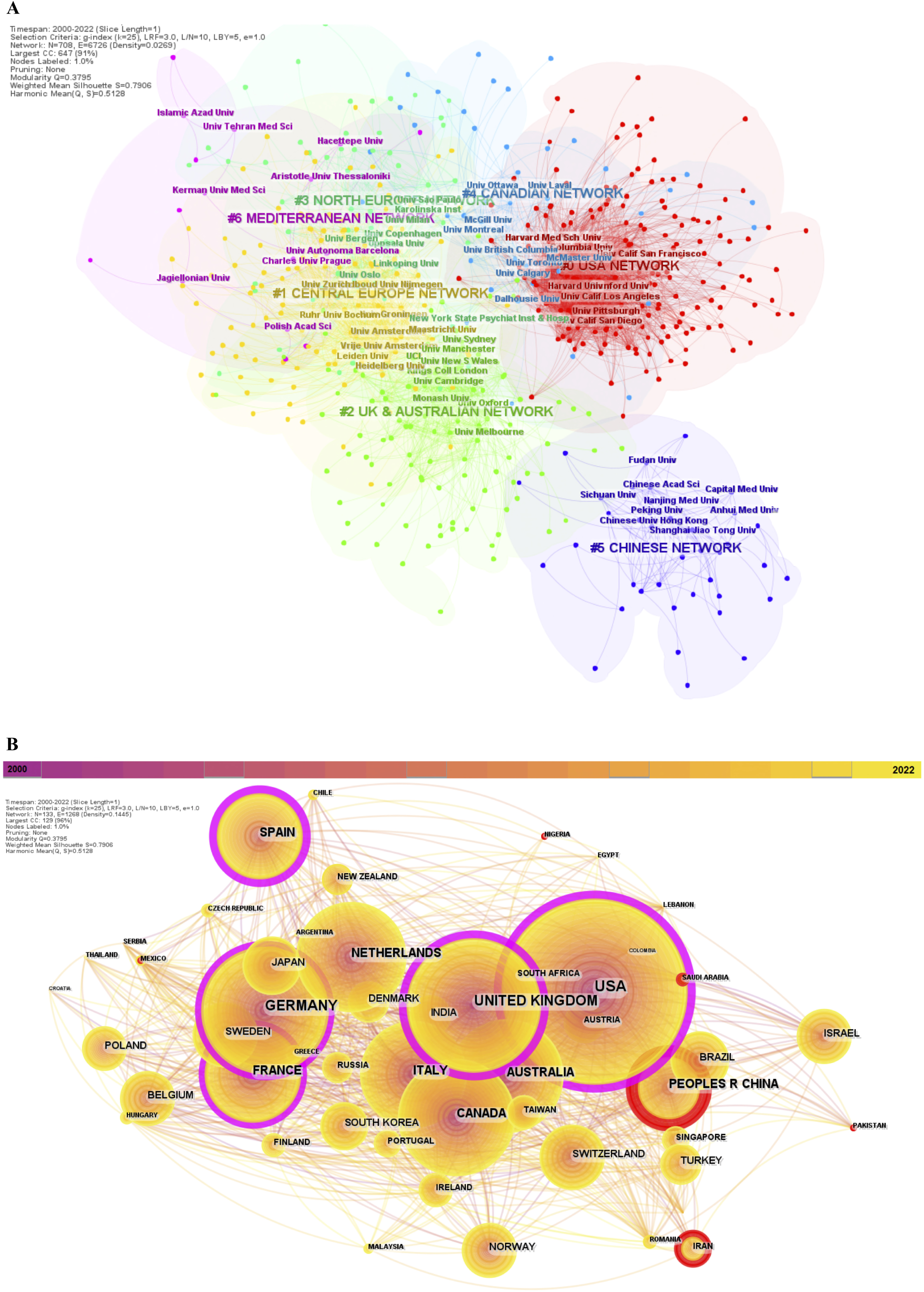
Co-authors’ institutions (A) and countries (B) network between the year 2000 to 2022. (A) Identified clusters from co-authors’ institutions network. Each node in this analysis corresponds to one authors’ institution and the links between the nodes represent co-authoring institutions. (B) Collaborative network between countries. Each node in this analysis corresponds to one authors’ country and the co-authoring countries are represented by the links between the nodes. Nodes with high betweenness centrality (i.e., those with significant influence on the network) generally have connections with two or more large groups of nodes. Citation burstness is depicted through red tree rings, whereby ring thickness is proportional to burst strength.

Seven major institutional network clusters were discovered from the analysis (ranked from the largest to smallest): 1) United States of America (USA) network, 2) Central Europe network, 3) United Kingdom (UK) and Australian network, 4) North Europe network, 5) Canadian network, 6) Mediterranean network, and 7) Chinese network. The network had an acceptable modularity score (Q > 0.3) and significant silhouette score (S > 0.6).

In the past 22 years, the top five institutions with the highest centrality all belonged to the USA network, namely Harvard University, University of California - Los Angeles, Yale University, Emory University, and University of North Carolina. The top five institutes with the strongest citation burst as ranked by the starting year of bursts was the University of Iowa (burst strength = 15.95; duration = 2000 - 2011), Cornell University (14.99; 2000-2010), University of Munich (13.58;2000-2016), Duke University (12.12;2000-2011), and Harvard University (41.33;2001-2015); the top five institutions with the highest citation counts were King’s College London (414 counts), University of Toronto (357), University of Pittsburgh (321), Karolinska Institute (292), and University of California, Los Angeles (280); the top five institutes with the strongest citation bursts were Harvard University (burst strength = 41.33), Harvard Medical School (35.07), University of New South Wales (29.73), University of Texas (21.49), and Sapienza University of Rome (16.13); and the top five institutes with the highest sigma scores were Harvard University (sigma = 30.0), Duke University (1.64), University of Pittsburgh (1.54), Columbia University (1.51), and Yale University (1.48).

The co-cited country network clusters had an acceptable modularity score (Q > 0.3) and significant silhouette score (S > 0.6). USA had the highest centrality (0.29), followed by the UK (0.25) and Spain (0.22). The top five countries with the strongest citation burst as ranked by the starting year of bursts was the USA (burst strength = 9.43; duration = 2002 - 2002), the UK (7.41;2007-2007), New Zealand (3.57;2008-2009), Argentina (3.67;2013- 2014), and Columbia (3.21;2017-2020); the top five with the highest citation counts was the USA (7,497 counts), the UK (2,266), Germany (1,903), Australia (1,484), and the People’s Republic of China (PRC) (1,390); the top five countries with the strongest citation bursts was the PRC (burst strength = 90.0), Iran (19.24), USA (9.43), UK (7.41), and Saudi Arabia and the top five countries with the highest sigma scores were the PRC (sigma = 37.71), Mexico (1.61), Saudi Arabia (1.19), Iran (1.11), and Argentina (1.04).

## 4. Discussion

### 4.1 Summary of main findings

An extensive overview of the development and evolution of research on BPSD in MCI since the first related article in 1980 until the year 2022 was produced using CiteSpace, VOSviewer, and Bibliometrix R package. In this study we identified the major hotspots and trends, as well as the leading keywords, authors, institutions, and countries in the research field.

Publications related to BPSD in MCI significantly increased from the 1990s. The most cited keyword for the literature was ‘depression’, followed by ‘disorder’ and ‘anxiety’, and the *Journal of Affective Disorders* had the highest number of publications in this field. The 5 highest co-cited authors were Zhang Y, Wang Y, Zhang J, Li Y, and Wang J, while the top 5 co-authors with highest influential potential were Cuijpers P, Hollon S, Andersson G, Wang L, and Hughes J. The USA had the highest centrality and citations, followed by the UK. More recently, the PRC exhibited notable growth, achieving a place in the top 5 countries with the highest citation counts, and has the strongest citation burst and the highest influential potential, surpassing other countries. Harvard University was the institute with the highest centrality, strongest citation burst, and highest influential potential, while King’s College London was the most cited institute of all time.

The co-cited reference network generated in this study captures the evolution and connection between 51 research clusters. Five observable major research trends were derived from the network: 1) Symptoms; 2) Diagnosis; 3) Brain Substrates; 4) Biochemical Pathways; and 5) Interventions. Throughout the years, research on BPSD in MCI progressed from studies on psychometric tests, chronic fatigue syndrome, and animal models in the 1990s/early 2000s to gut microbiota, e-health, COVID-19, cognition, and delirium within recent years.

### 4.2 Symptoms Domain

The ‘Symptoms’ domain refers to a broad range of individual features that make up BPSD or are closely related to those symptoms. Research clusters on emotions, emotional disturbances, depression, anxiety, sleep, and delirium derived from the analysis fall into this category.

Studies have focused on the correlation between emotion recognition and BPSD prevalence in MCI as past literature have suggested that these impairments are correlated with isolation, difficulties in interpersonal relationships, higher levels of depression (Chiu et al., 2006; Liu et al., 2022), and psychological and behavior disturbances (Shimokawa et al., 2001). However, these associations remain unclear as a recent systematic review found conflicts between the literature - some studies reported emotion-specific impairments in MCI individuals, whilst others did not find any differences between MCI and normal controls (Morellini et al., 2022).

Regarding individual BPSD, it has been reported that the onset of each symptom differs between disease stages, and the earliest ones that manifest at the aMCI stage are apathy, depression, irritability, sleep disturbance, and agitation, whilst the rest occur later (Chen et al., 2021). These findings corroborate the presence of the research clusters in our analysis and most likely explain the absence of clusters of the other neuropsychiatric symptoms.

### 4.3 Diagnosis Domain

The ‘Diagnosis’ domain relates to the comorbidities and clinical conditions associated with BPSD exhibited in MCI individuals. This domain is considered the largest as it encompasses a majority of the research hotspots from the analysis, such as multiple sclerosis, social anxiety disorder, insomnia, COVID-19, cancer, post-traumatic stress disorder (PTSD), autism, fragile X syndrome, fibromyalgia, respiratory failure, bariatric surgery, pregnancy, traumatic brain injury (TBI), and amyotrophic lateral sclerosis (ALS).

MCI individuals generally exhibit multiple comorbidities. It has been proposed that these coexisting conditions may significantly affect the conversion of MCI into dementia and contribute to the severity of neuropsychiatric symptoms. Therefore, comorbidities are an important factor to consider in both research and clinical practice. Some frequently observed comorbidities found in those with MCI include, but are not limited to, cardiovascular diseases, TBI, diabetes, hypertension, depression, and obesity (Katabathula et al., 2023). Furthermore, authors of the same study found that when MCI participants were computationally clustered into subtypes according to their comorbidities and features, those who had a ‘poor’ prognosis either stayed at the MCI stage or progressed to AD, whereas those with the ‘best’ prognosis never developed AD and either reverted to a normal cognitive stage or remained in the MCI stage.

Interestingly, a new research area that emerged within the last few years is the relationship between COVID-19, BPSD, and MCI. Articles exploring this area have documented higher frequencies of cognitive impairment and increased BPSD prevalence and severity after COVID-19 infection (Henneghan et al., 2022; Kuroda et al., 2022a; Tavares-Junior et al., 2022). Nevertheless, the long-term effects of COVID-19 are still to be determined.

### 4.4 Brain Substrate Domain

The ‘Brain Substrate’ domain encompasses neurological pathways and brain regions related to mental and emotional processing. Research on the prefrontal cortex, amygdala, hippocampus, anterior cingulate cortex, and ventral tegmental area relate to this domain.

Anatomical brain changes may contribute to cognitive impairment and neuropsychiatric symptoms, and conversely, the same may apply. Neuroimaging studies have observed changes in multiple brain regions of individuals with MCI, with the most common being attenuated hippocampal volume compared to controls (Ries et al., 2008). The same review also reported attenuated medial temporal lobe volume in the amygdala, parahippocampal gyri, fusiform gyrus, and entorhinal cortex in MCI.

Regarding the relationship between neuropsychiatric symptoms and brain morphology in MCI, recent studies found that (1) depressive symptoms were associated to thickness of the left superior temporal sulcus banks and accumulation of amyloid-β in the frontal lobe; (2) psychosis had a positive correlation with left post-central volume; and (3) elation symptoms were related to thicker right anterior cingulate cortex (Chung et al., 2015; Siafarikas et al., 2021).

Apart from understanding the extent of clinical outcomes from brain changes, atrophy rate in particular brain regions may also serve as a good predictor of disease progression (Ries et al., 2008).

### 4.5 Biochemical Pathways Domain

The ‘Biochemical Pathways’ domain includes the metabolic pathways and biochemical compounds related to BPSD in MCI: serotonin, kynurenine pathway, microbiota gut-brain axis, brain-derived neurotrophic factor (BDNF), neurotoxicity, neurotransmitters, microglia, and the glucocorticoid receptor.

The pathophysiological changes that accompany MCI comprise multiple cellular pathways. One study reported up to 342 and 351 significantly altered plasma and cerebrospinal fluid (CSF) metabolites, respectively, translating to 23 altered canonical pathways in plasma and 20 in CSF (Trushina et al., 2013). Some of the disturbed pathways include neurotransmitter and amino acid metabolism, Krebs cycle, lysine metabolism, tryptophan metabolism, cholesterol and sphingolipids transport, energy metabolism, and mitochondrial function. These disturbances may not only affect the severity of cognitive impairment but could also influence neuropsychiatric symptoms, for example in serotonin neurotransmission deficits (Rodríguez et al., 2012).

### 4.6 Interventions Domain

Pharmaceutical and non-pharmaceutical interventions for neuropsychiatric symptoms are encompassed within the ‘Interventions’ domain. These treatment options include both patient- and caregiver-focused interventions. Research clusters in this domain comprise cognitive therapy (mindfulness-based), cognitive behavioral therapy (for insomnia, digital), antidepressants (MAOIs, imipramine), psychotherapy, exposure therapy (virtual reality), digital interventions, self-help (online), group counseling, cognitive emotion regulation strategies, and work-focused interventions.

Both types of interventions have been extensively studied. A systematic review looking at eight non-pharmacological interventions and seven pharmacological interventions reported that global BPSD seemed to be relatively improved with functional analysis-based interventions, music therapy (some uncertainty due to low-quality evidence), analgesics, donepezil, galantamine, and atypical antipsychotics (Dyer et al., 2018).

Although pharmaceutical options are quite effective, it also causes higher rates of adverse events (Seibert et al., 2021), thus, clinical guidelines generally recommend the use of non-pharmacological approaches as the first line of treatment. With the advancement of technology, newer options have incorporated internet-based solutions, such as internet-delivered cognitive behavioral therapy (ICBT), which may increase accessibility and ease of use. Nevertheless, more research is required to gauge whether online tools are equivalent to face-to-face therapy (Carlbring et al., 2018).

### 4.7 Strengths of Scientometric Studies

To the best of our knowledge, this is the first scientometric study on the BPSD in MCI. The scientometric approach offers a comprehensive insight into the research field that cannot be obtained from systematic reviews, meta-analyses, and guidelines alone. Networks retrieved from this type of analysis enables the visualization of significant research focus areas and connections between co-authors and co-authoring countries and institutions. There are multiple ways in which investigators can utilize this information to help with current and future research as well as clinical practices. One example is that investigators can decide the direction of their future research by using the co-cited reference networks, information on articles with the highest centrality, and the co-occurring author keywords network to identify saturated research hotspots (such as emotion, amygdala, and glucocorticoids) and newer topics of research that are starting to gain traction (such as COVID-19, delirium, and cognition). Moreover, knowledge of the top co-cited authors, institutes, and countries from the analysis enables investigators to further recognize leading individuals and research bodies in the field who may be promising candidates for any prospective collaborations.

An alternative example is the use of reference and keywords networks in the clinical setting to look at related comorbidities and clinical conditions to help the physician decide whether these conditions should be addressed as part of the patient’s care to reduce the severity of neuropsychiatric symptoms or cognitive impairment. Furthermore, in the scenario where a patient presents an unusual clinical profile, physicians can determine those with the greatest domain knowledge from the network of co-cited authors, institutes, and countries, and contact them for more information or to initiate a collaborative case study. Regardless of its use case, it is recommended that the first step in any research is to conduct a scientometric analysis in the field of interest to help clarify the research scope and direction.

### 4.8 Limitations and Future Research

The database search in this study was only conducted in WOSCC as it is one of the largest accessible citation databases that provides full text and citation information. Furthermore, information from other databases is provided in a different format, making it complicated to merge datasets. Nevertheless, the use of only one database entails that certain significant publications related to BPSD in MCI may have been excluded from the study. Once the desired features become available in other databases and the datasets can be merged seamlessly, an updated scientometric study should be conducted to ensure study completeness.

Additionally, the following should also be considered: (1) influence of all authors may not have been adequately characterized as the co-citation analysis only looks at first authors; (2) the newest trends may not have been reported as more recent publications take time to be adequately cited; and (3) some keywords may have been unintentionally excluded as they may have multiple variations.

Regarding the scientometric approach, a major limitation is that it relies on citation-related indicators, which entails that there is a certain level of citation bias. To elaborate, particular references could be cited to increase the value and credibility of the manuscript regardless of its relevance and quality, decreasing the recognition of other work that may be more significant. Factors such as self-citation, author authority, institution, reviewer suggestions, and journal all contribute to citation bias (Urlings et al., 2021).

Bias against novelty and publication bias are two other potential sources of bias. In research, there is bias against novelty as highly novel papers have delayed recognition, higher citation variance, may get published in journals with lower Impact Factor, and tend to be significantly cited in “foreign” fields than their “home” fields (Wang et al., 2017). For publication bias, studies with significant results are favored in publication over non-significant ones by editors, reviewers, authors, and institutions for reasons such as lack of interest or conflicting results that are unaligned with specific agenda (DeVito and Goldacre, 2019). This bias could further lead to the dismissal of non-significant results and exaggeration of significant results, impacting the quality of subsequent studies that cite them.

Although the biases cannot be prevented, these can still be limited and accounted for in future analyses by combining newer approaches to scientometrics such as webometrics, infometrics, and altmetrics (Vakhrushev, 2019) to achieve more accurate metrics.

## 5. Conclusion

Using the scientometric approach, our study provides a comprehensive overview of the research evolution, trends, and hotspots within the field of BPSD in MCI. The first paper relating to this field was published in 1980 and the number of publications significantly increased throughout the subsequent years. The largest co-cited reference network cluster was on cognitive disorders and five research domains were derived from the network: symptoms, diagnosis, brain substrates, biochemical pathways, and interventions. The most cited keyword among all publications was ‘depression’ and the current research focus is on gut microbiota, e-health, COVID-19, cognition, and delirium. The USA and UK had the highest influence and citations, whilst the PRC exhibited notable growth with increasing influence. Altogether, this study can help investigators and clinicians gain a better understanding of the research body on BPSD in MCI to make better informed decisions and contribute to refining the quality of work in this area.

## Data Availability

The dataset will be available from M.M. upon reasonable request and once the data has been fully analyzed by the authors.

## 6. Declarations

### 6.2 Declaration of Generative AI and AI-assisted Technologies in the Writing Process

None

## 7 Acknowledgements

None

