## Supplementary Information for "Neuropsychiatric Disturbances in Mild Cognitive Impairment: A Scientometric Analysis"

### 1. Methods

*1.1 Database Selection*

Complete publication records including the keywords and full citations are needed to conduct a scientometric study. A handful of electronic databases can provide the information required (e.g., Web of Sciences and Scopus). However, the format, extent of information, and year range of publications differ between these databases, making it difficult to combine publication information from difference sources. Accordingly, we chose Web of Science (WOS) for this study as it is one of the largest accessible citation databases, with some publications dating back to the 1900s, and has a slightly lower frequency of incorrect DOIs compared to Scopus (Pranckutė, 2021).

*1.2 Database Search and Further Filtering*

An extensive search was done for all literature related to terms and phrases associated with both behavioral and psychological symptoms of dementia (BPSD) and mild cognitive impairment (MCI). The following search terms, with the combination of keywords and MeSH terms, were selected and chosen for the WOSCC search:

(mild cognitive* OR cognitive impairment OR neurocognitive deficit OR cognitive deficit OR cognitive disorder OR minor neurocognitive disorder OR minor neurocognitive deficit) AND ("behavioural and psychological symptoms of dementia" OR "behavioral and psychological symptoms of dementia" OR delusion OR hallucination OR agitation OR depression OR anxiety OR apathy OR irritability OR euphoria OR disinhibition OR aberrant motor* OR night time disturbance OR sleep disturbance OR eating abnormalities* OR abnormal eating) NOT (major depressive OR bipolar OR schizoaffective OR schizophrenia OR post traumatic* OR generalized anxiety disorder OR panic* OR obsessive compulsive*) NOT (Alzheimer* OR stroke OR hungtington OR parkinson* OR dementia) NOT (comment OR commentary OR letter) NOT TITLE (animal* OR aethiop* OR amphibian* OR alligator* OR antilope* OR aplysia OR armadillo* OR anuran* OR "aedes albopictus" OR ape* OR bat* OR badger* OR baboon* OR bear* OR beaver* OR beagle* OR buffalo* OR bovine* OR *bird* OR broiler* OR buzzard* OR binturong* OR budgerigar* OR carp* OR camel* OR canine* OR cat* OR calve* OR chinchilla* OR callithrix OR cow* OR captiv* OR chamois OR chimpanzee* OR chick* OR crab* OR crustac* OR coyote* OR crocodyl* OR croaker* OR duck* OR drosophil* OR deer* OR duiker* OR dog* OR eagle* OR elk* OR ewe* OR elasmobranch* OR equine* OR elephant* OR egg* OR fox* OR *fish* OR fowl* OR feline* OR foal* OR ferret* OR frog* OR fundulu* OR galliwasp OR gastropod* OR gazelle* OR geese OR goat* OR gilthead* OR gecko* OR greyhound* OR hawk* OR horse* OR hemiptera* OR hamster* OR honeybee OR heterocapsa* OR heteropt* OR impala* OR insect* OR ibex* OR kitten* OR leopard* OR "lepus europaeus" OR llama* OR larva* OR lamb* OR lymnaea* OR lion* OR lizard* OR lemur* OR lateolabrax* OR jackal* OR ostriche* OR otter* OR gorilla* OR pup* OR mice* OR mouse* OR macaque* OR marmoset* OR mollusk* OR macropod* OR "mugil cephalus" OR marte* OR monkey* OR meloidogyne* OR nile OR "nonhuman primate*" OR "non-human primate*" OR nematode* OR "nacobbus aberrans" OR otolemur* OR rabbit* OR swine* OR opossum* OR psyllid* OR ruminant* OR reindeer* OR reptile* OR tetranychu* OR tilapia* OR turtle* OR thripidae* OR ungulate* OR otariid* OR opossum* OR panther* OR periwinkle OR pteropus OR pseudococcidae* OR periplaneta* OR possum* OR prawn* OR parrot* OR panda* OR poni* OR pagrus* OR *pig* OR primate* OR peacock* OR porcin* OR rat* OR rodent* OR serval* OR snake* OR specie* OR seal* OR sterlet OR salmon* OR snail* OR suricat* OR slider* OR solea* OR sheep* shrimp* OR sturgeon* OR springbok* OR squirrel* OR "scophthalmus rhombus" OR swainson* OR tapir* OR tasmanian* OR tortoise* OR tiger* OR tephritidae* OR veterinar* OR wild* OR *wolf* OR wolv* OR xenop* OR zebra*)

The dataset from WOSCC was extracted in a plain text file containing full references and citations.

To ensure that the dataset only contained literature relating to BPSD in dementia-related MCI in humans, Python (3.9.14) was used for further removal of duplicates and unrelated publications. Studies that had the following terms in their title were further removed:
‘rat', 'rats', 'mouse', 'mice', 'murine', 'monkey', 'monkeys', 'marsupial', 'adhd', 'attention deficit', 'attention-deficit', 'drosophila', 'autistic', 'autism', 'oncology', 'cancer', 'tumour', 'tumor', 'tumours', 'tumors', 'animal', 'major depressive', 'bipolarity', 'schizophrenia', 'ptsd', 'anxiety disorder', 'anxiety disorders', 'obsessive compulsive', 'pregnant', 'pregnancy', 'maternal', 'menstrual', 'premenstrual syndrome', 'traumatic brain injury', 'tbi', 'diabetes', 'concussion', 'multiple sclerosis', 'ms', 'fibromyalgia', 'acromegaly', 'behcet', 'bechet', 'dependence', 'icu', 'hiv', 'alcohol', 'alcoholic', 'alcoholics', 'alcoholism', 'substance', 'glioma', 'pulmonary disease', 'bowel syndrome', 'epilepsy', 'epileptic', 'seizure', 'seizures', 'psoriasis', 'trichotillomania', 'copd', 'ataxia', 'lupus', 'lyme', 'haemorrhage', 'hemorrhage', 'major depression', 'prenatal', 'infant', 'infants', 'fetal', 'fetus', 'foetal', 'foetus', 'baby', 'babies', "baby's", 'children', 'childhood', 'child', 'youth', 'adolescent', 'adolescence', 'adolescences', 'pediatric', 'paediatric', 'pediatrics', 'paediatrics', 'girl', 'girls', 'boy', 'boys', 'restless legs', 'restless leg', 'abuse', 'abusive', 'abusers',

'posttraumatic stress', 'personality disorder', 'eating disorder', 'police', 'dystrophy', 'phenylketonuria', 'anorexia nervosa', 'bulimia', 'motor neuron disease', 'nurse', 'nurses', 'concussion', 'postconcussion', 'head injury', 'head injuries', 'head trauma', 'hypoxic-ischaemic encephalopathy', 'inflammatory bowel disease', 'ibd', 'seasonal affective', 'craniotomy', 'neurosurgery', 'posterior fossa surgery', 'intracranial aneurysm', 'radiotherapy', 'prison', 'prisoner', 'gamblers', 'mdma', 'university', 'student', 'students', 'ecstasy', 'ketamine', 'stutter', 'stuttering'.

All highly cited literature from the resulting dataset were examined in greater detail to check for relevance and homogeneity. Additionally, the authors also selected a random representative sample to confirm the dataset quality.

*1.3 Inclusion and Exclusion Criteria*

There were no limitations on the total dataset size, and the time of publication, language, and population of the literature included in this scientometric study. Articles, reviews, editorial materials, and proceeding papers related to BPSD in MCI (in line with the search terms above) were included.

Unrelated literature or those considered outside the scope of the study (according to the search terms and expertise of the authors) were excluded from the study. Commentaries, letters, magazines, and monographs were excluded.

*1.4 Software and Data Analysis*

Bibliometrix R packages (3.1.4) is an open-source tool and was used for quantitative research. More specifically, publication outputs from the dataset are analyzed using this tool.

CiteSpace (6.1.R3) is a Java application that was used to produce and analyze visual maps and patterns in the literature. Co-citation references networks, the co-occurrence authors keywords network, the co-authorship network, the co-authorship institution network, and the co-authorship country network were produced using CiteSpace. VOSviewer (1.6.16) is an application that constructs and visualizes bibliometric networks. The application is used in this study to develop the co-occurring authors keywords network and the co-authorship network.

#### *1.5 CiteSpace Parameters*

The CiteSpace paramters used for the analysis in this study were the following:

1. Link retaining factor = 3.0
2. Look back years = -1
3. Timespan = 1980 – 2022 or 2020 – 2022 with one slice per year
4. Links
    a. Strength = cosine
    b. Scope = within slices
5. Selection Criteria
    a. g-index scale factor k = 25
    b. Maximum of selected items per slice = 100
6. Minimum duration = 5 years

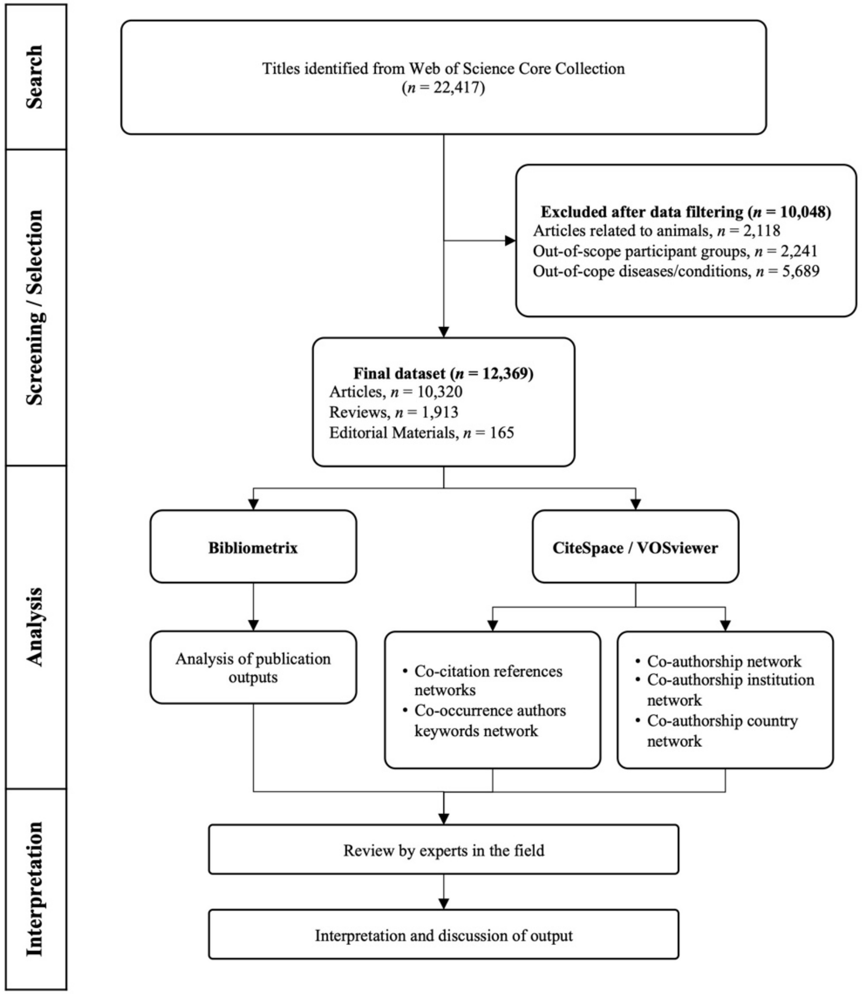

**Supplementary Figure 1.** Scientometric study flowchart

**Supplementary Figure 2.** Details of the major 16 clusters in the co-cited reference network (1980-2022) ranked by cluster size
*The chosen cluster name and top five extract terms (with its corresponding log-likelihood ratio and p-level) is reported for each cluster. Extent of burstness is represented by red tree-rings around the nodes. Nodes with citations bursts or are highly cited are considered important nodes. The color scale from pink to yellow represents the time slices from 1980 to 2022, respectively.*

**Cluster 0 • "Cognitive Disorders"**: emotion (46.96, 1.0E-4); fmri (37.17, 1.0E-4); prefrontal cortex (30.76, 1.0E-4); insomnia (30.31, 1.0E-4); attention bias (29.64, 1.0E-4)

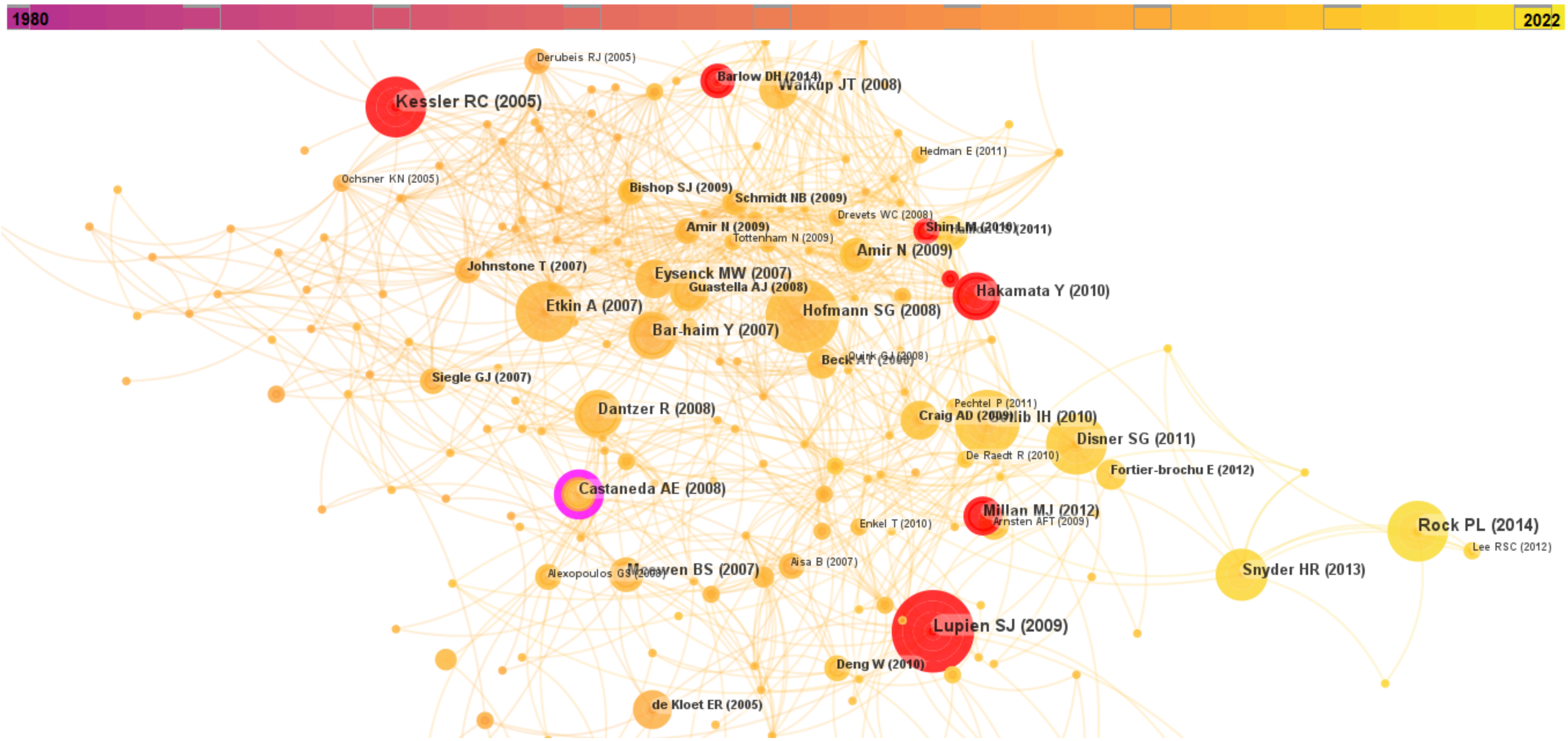

**Cluster 1 • "Internet-based Cognitive Behavioral Therapy"**: ehealth (59.08, 1.0E-4); telemedicine (50.5, 1.0E-4); mhealth (46.61, 1.0E-4); mental health (35.34, 1.0E-4); e-mental health (31.04, 1.0E-4)

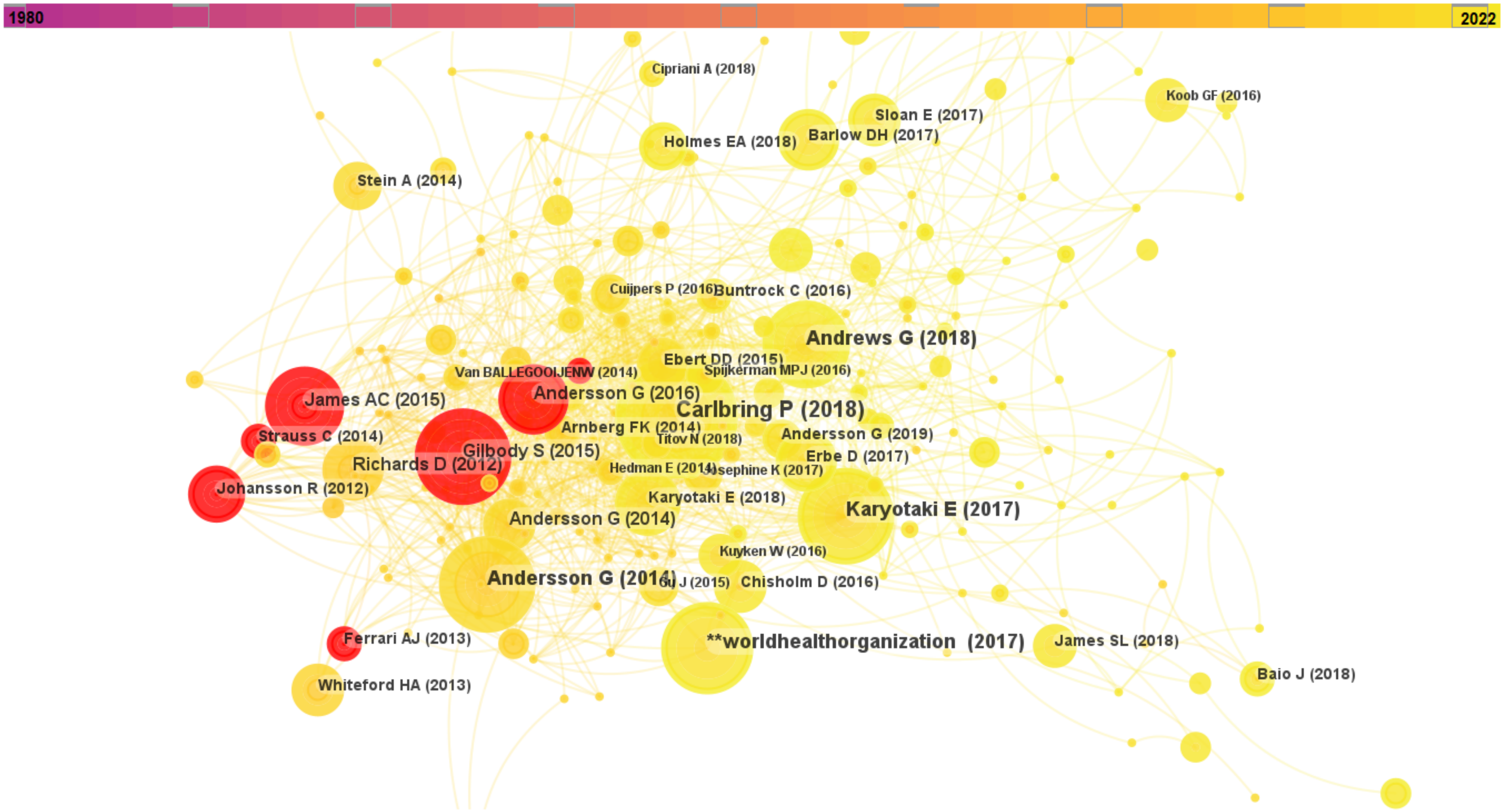

**Cluster 2 • “Mindfulness”**: internet (42.85, 1.0E-4); depression (30.62, 1.0E-4); cognitive therapy (29.06, 1.0E-4); mindfulness (27.75, 1.0E-4); mindfulness-based cognitive therapy (18.37, 1.0E-4)

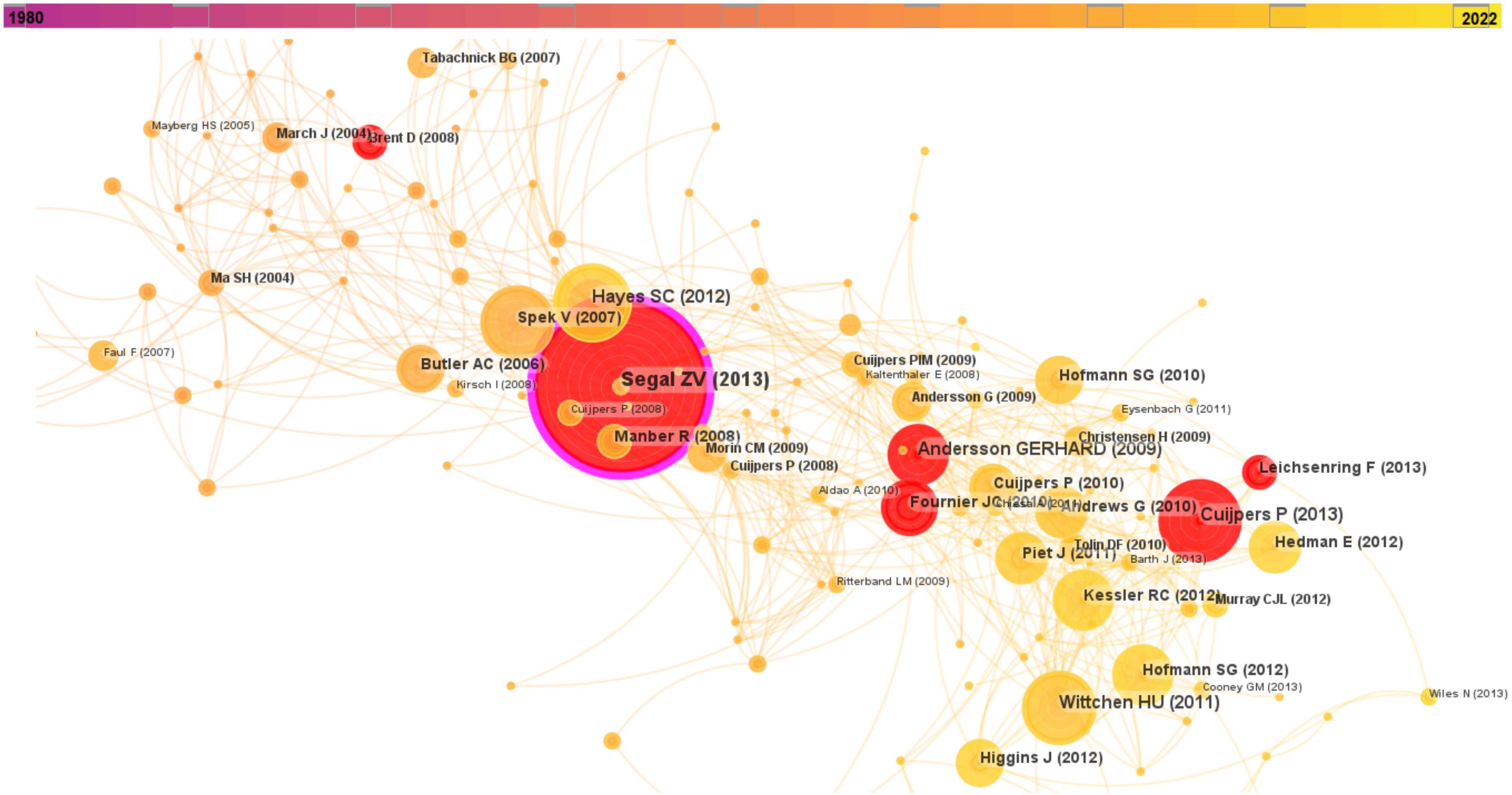

**Cluster 3 • “Multiple Sclerosis”**: multiple sclerosis (132.53, 1.0E-4); disability (23.03, 1.0E-4); cognition (18.17, 1.0E-4); fatigue (15.48, 1.0E-4); longitudinal study (13.62, 0.001)

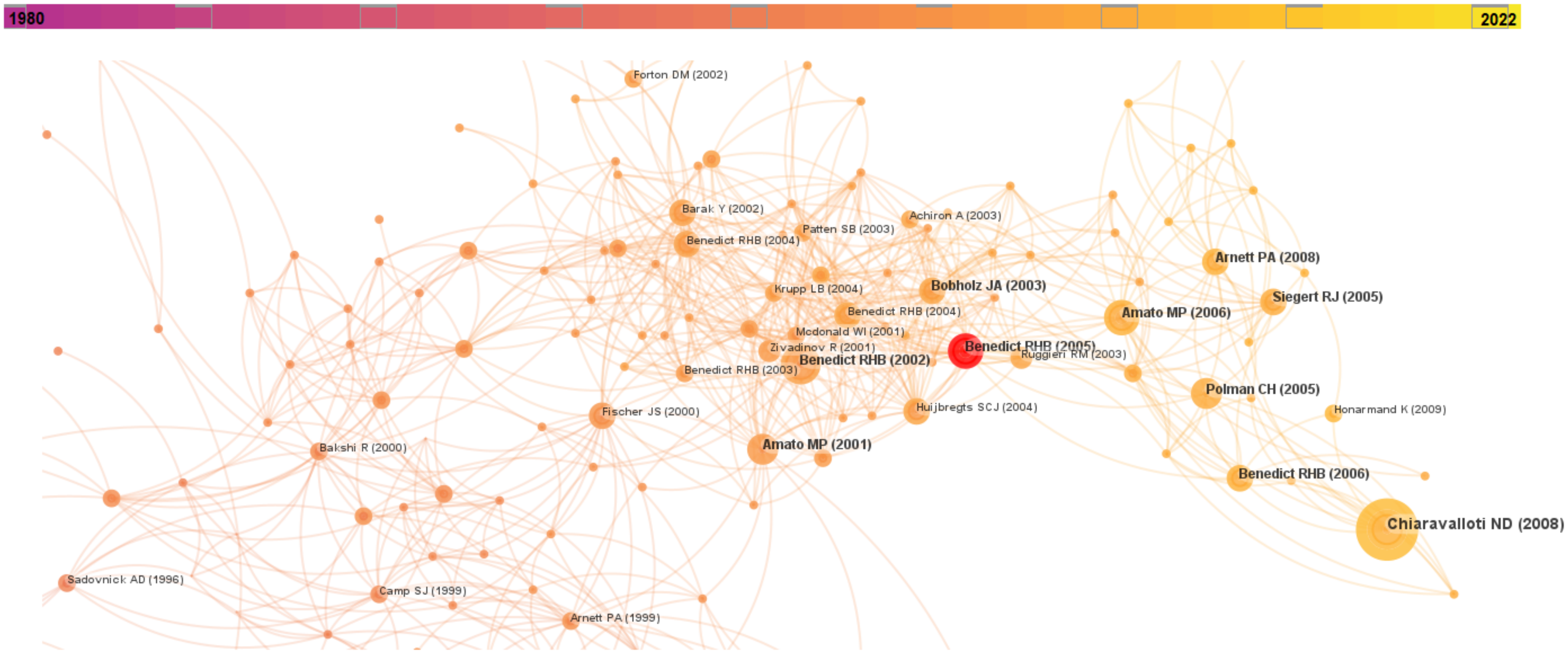

**Cluster 4 • "Amygdala"**: amygdala (38.33, 1.0E-4); fmri (32.39, 1.0E-4); social anxiety disorder (28.99, 1.0E-4); emotion regulation (27.13, 1.0E-4); chronic pain (25.83, 1.0E-4)

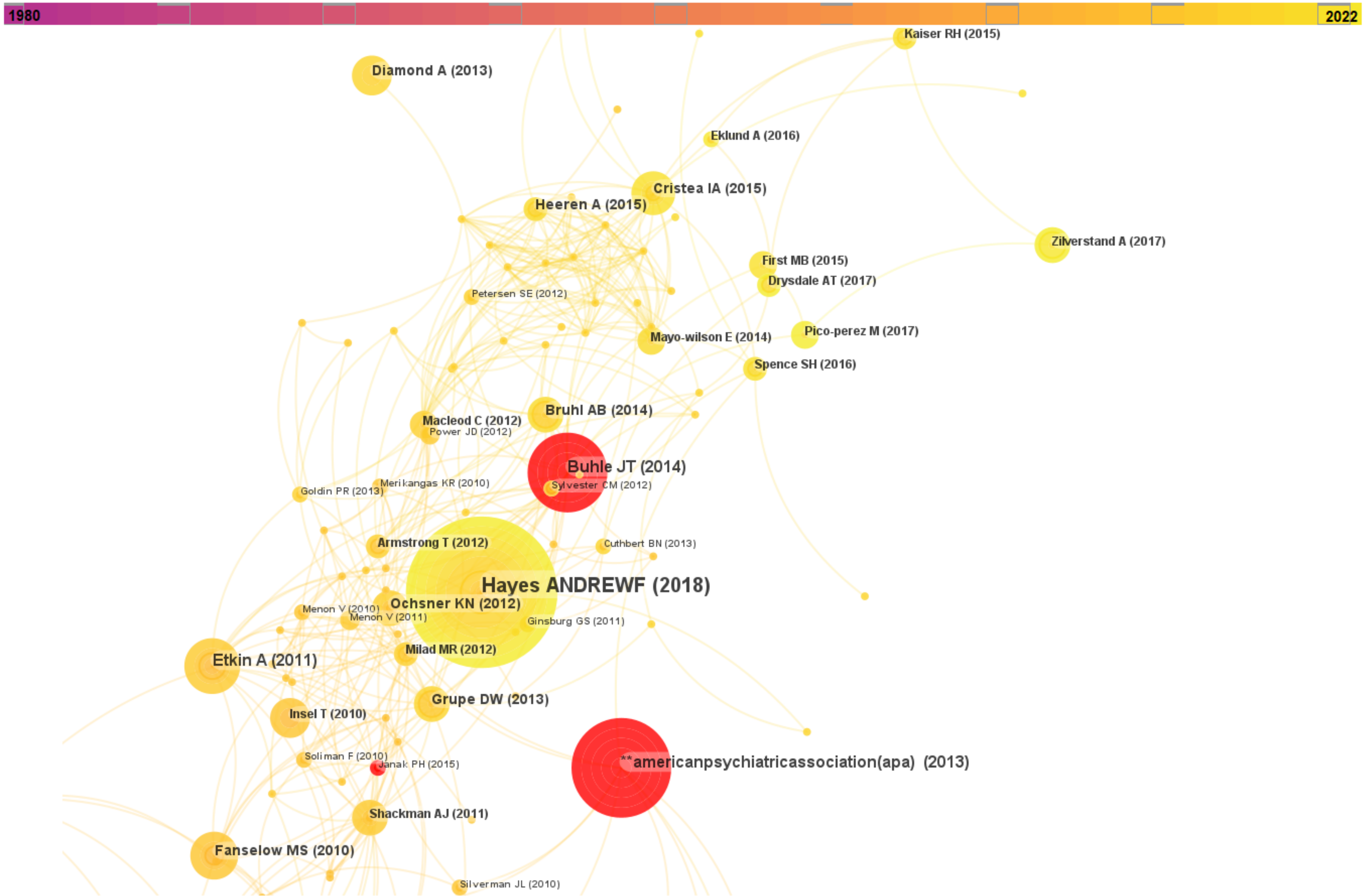

**Cluster 5 • “Chronic Fatgiue Syndrome”**: chronic fatigue syndrome (83.63, 1.0E-4); affective disorders (11.46, 0.001); cognition disorders (11.46, 0.001); cognitive deficits (9.86, 0.005); neuropsychological impairments (9.05, 0.005)

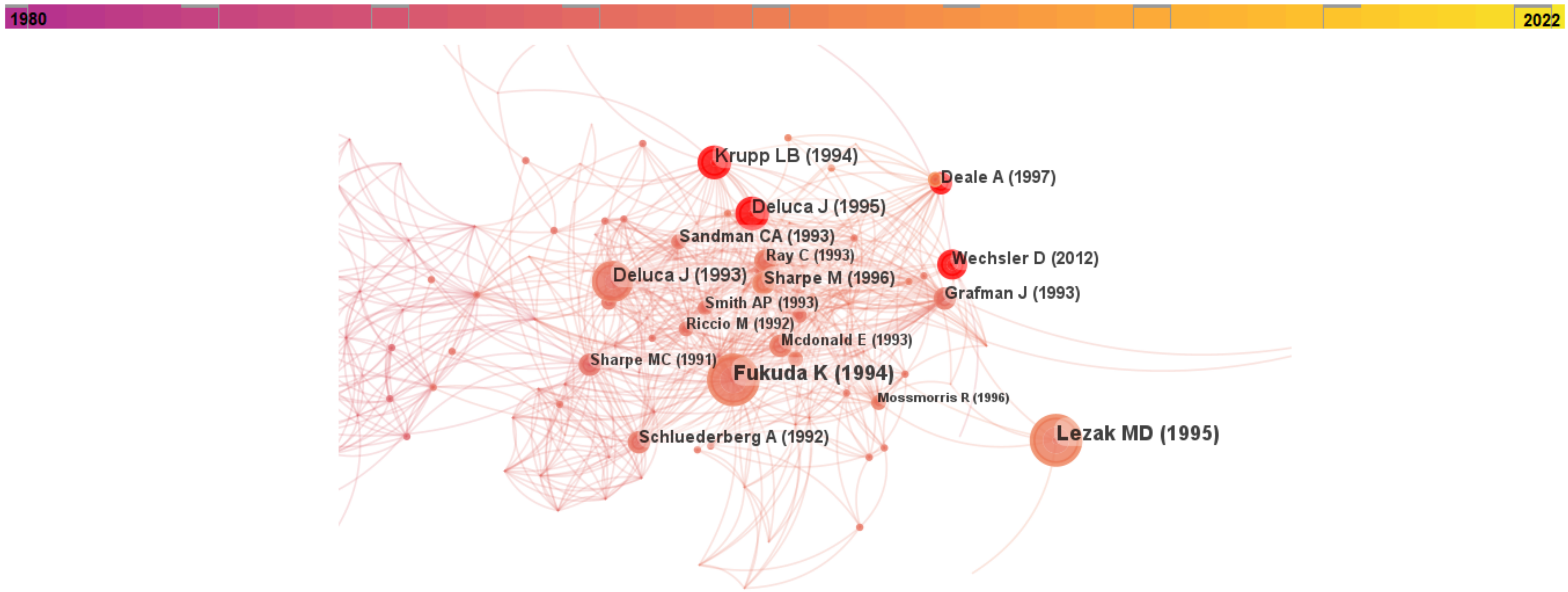

**Cluster 6 • "Neuropsychology"**: chemotherapy (24.15, 1.0E-4); anxiety (20.11, 1.0E-4); tbi (16.73, 1.0E-4); cognitive function (15.7, 1.0E-4); rehabilitation (14.67, 0.001)

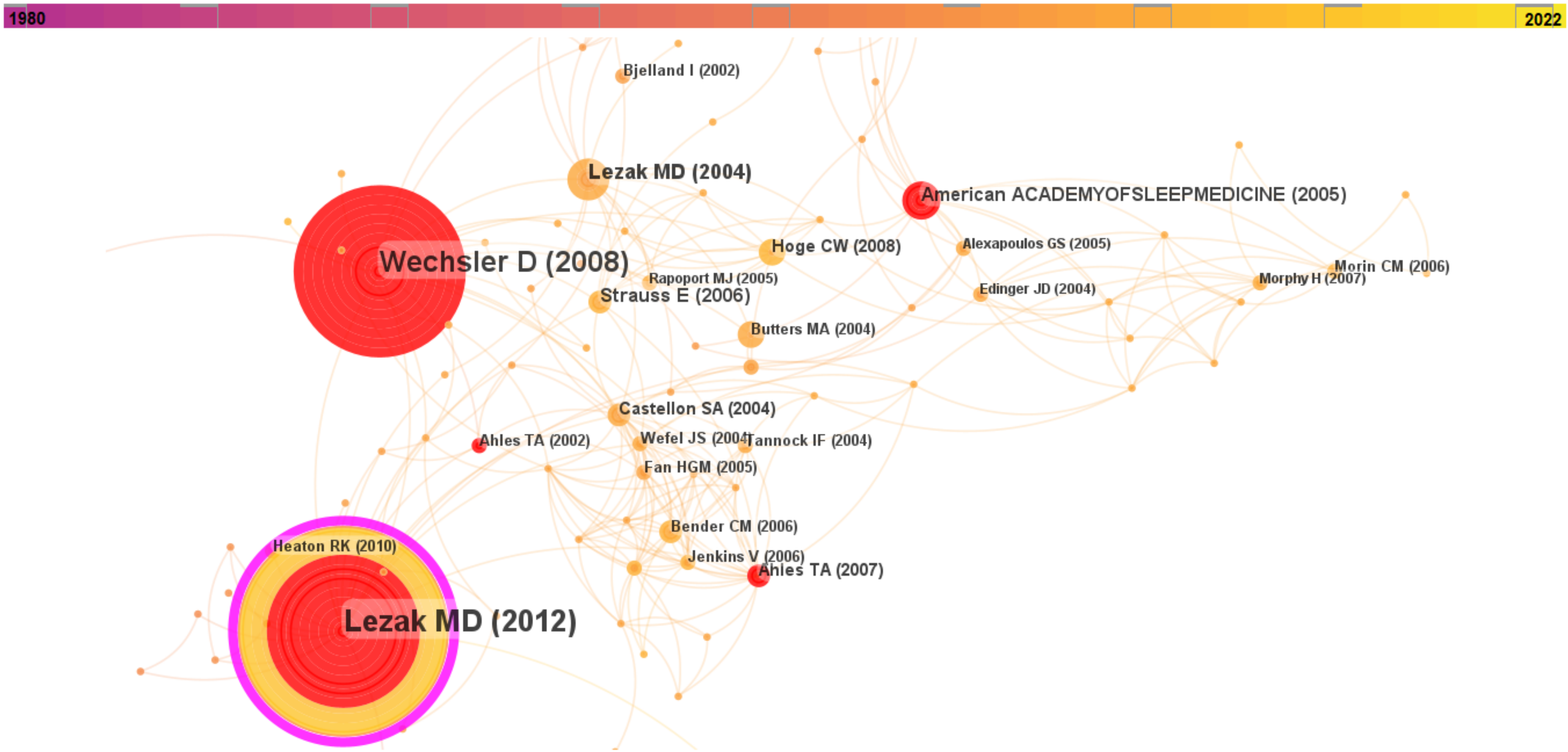

**Cluster 7 • "Glucocorticoid"**: glucocorticoid (23.58, 1.0E-4); hippocampus (17.8, 1.0E-4); glucocorticoids (16.93, 1.0E-4); human (9.1, 0.005); anxiety (8.59, 0.005)

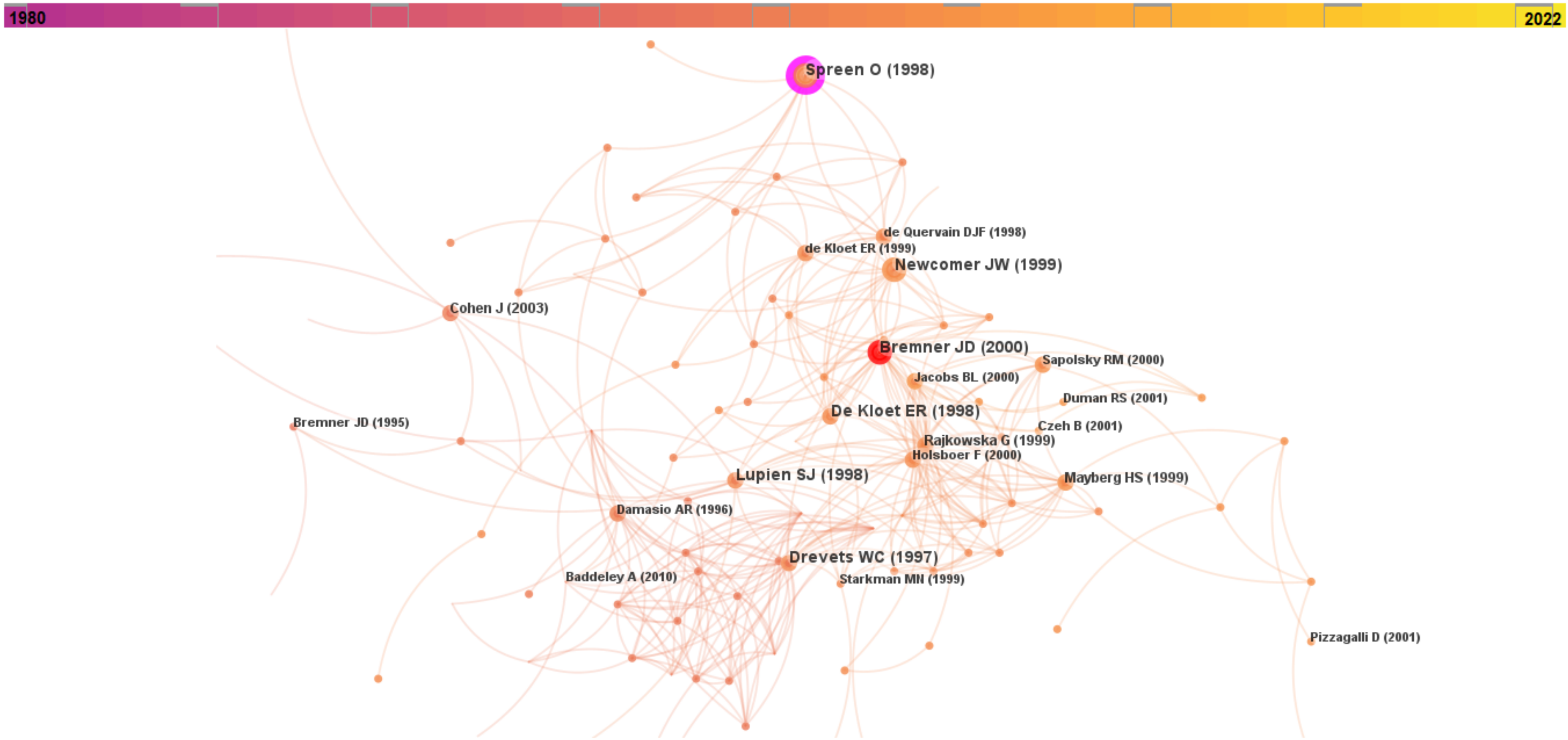

**Cluster 8 • "Insomnia"**: insomnia (273.46, 1.0E-4); sleep (112.28, 1.0E-4); cbt-i (54.89, 1.0E-4); chronic insomnia (24.07, 1.0E-4); sleep disorders (20.01, 1.0E-4)

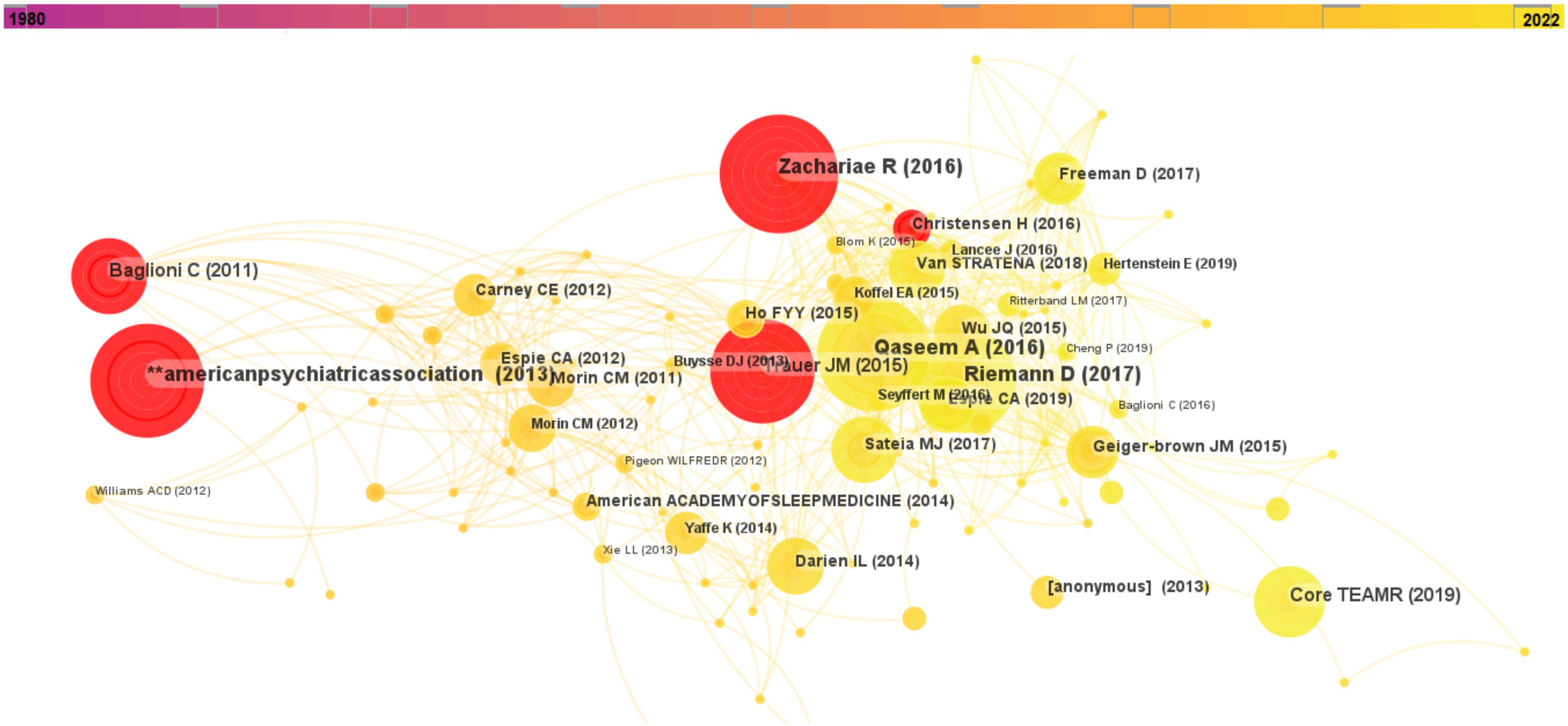

**Cluster 9 • "Cortico-limbic Pathway"**: mdma (33.65, 1.0E-4); ecstasy (24.42, 1.0E-4); serotonin (23.27, 1.0E-4); tryptophan (10.84, 0.001); inhibition (9.16, 0.005)

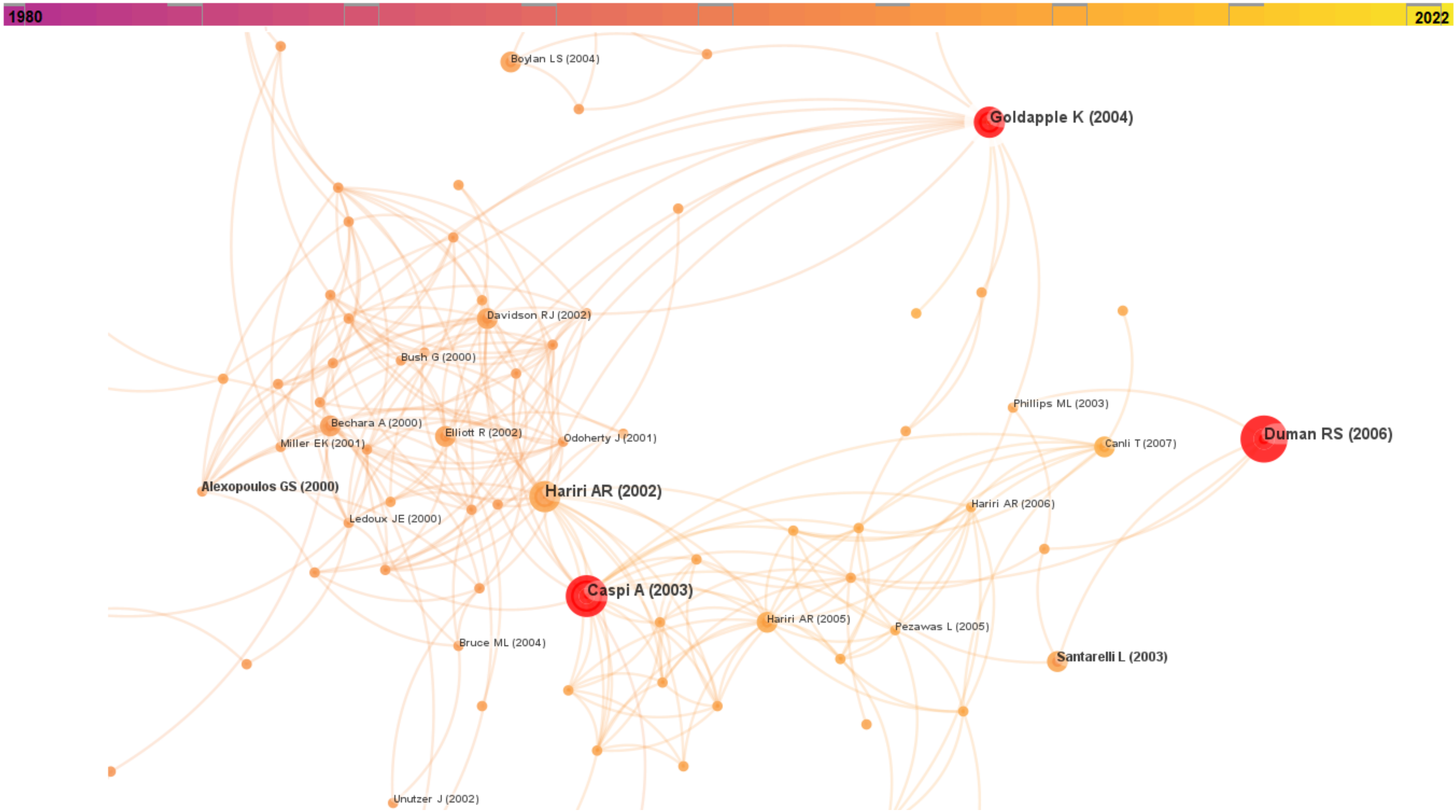

**Cluster 10 • "COVID-19"**: covid-19 (174.61, 1.0E-4); sars-cov-2 (57.73, 1.0E-4); long covid (44.86, 1.0E-4); covid-19 pandemic (20.67, 1.0E-4); post-covid-19 (19.18, 1.0E-4)

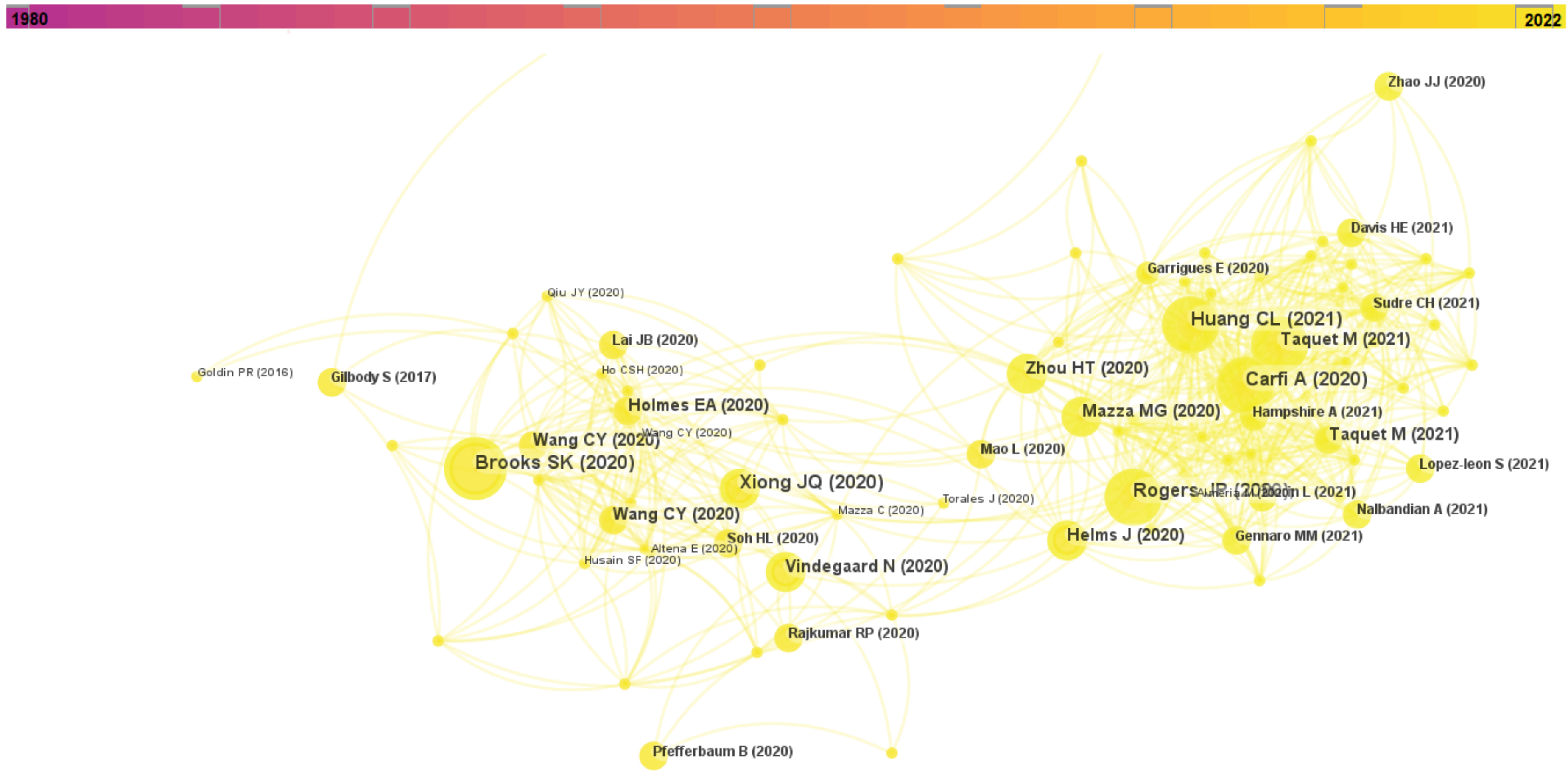

**Cluster 11 • “Cancer”**: breast cancer (51.63, 1.0E-4); cancer (49.22, 1.0E-4); chemotherapy (30.78, 1.0E-4); chemobrain (30.29, 1.0E-4); oncology (28.92, 1.0E-4)

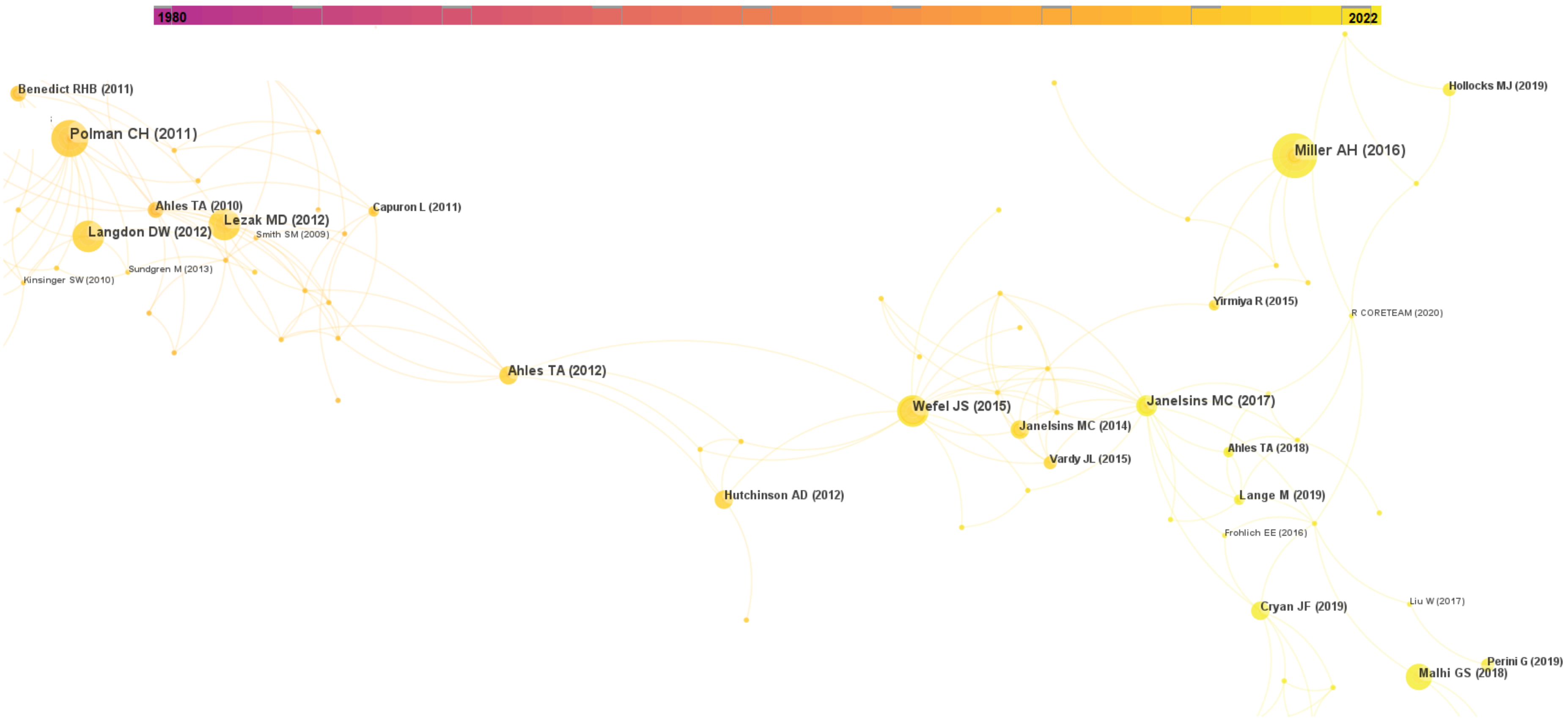

**Cluster 12 • "Social Phobia/Delusion"**: daytime functioning (20.02, 1.0E-4); sleepiness (13.33, 0.001); behaviour therapy (9.99, 0.005); wake (9.99, 0.005); maois (9.99, 0.005)

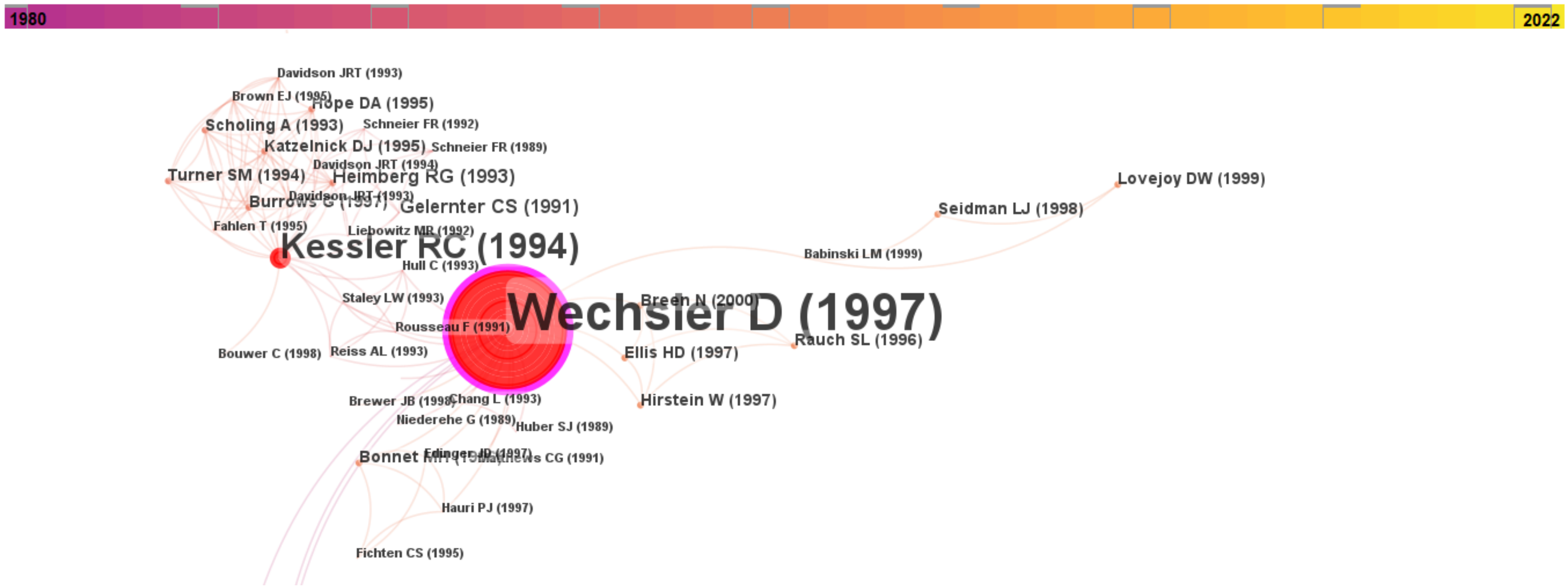

**Cluster 13 • "Post-traumatic Stress Disorder"**: ptsd (40.69, 1.0E-4); exposure therapy (39.88, 1.0E-4); posttraumatic stress disorder (31.58, 1.0E-4); veterans (24.99, 1.0E-4); virtual reality (23.24, 1.0E-4)

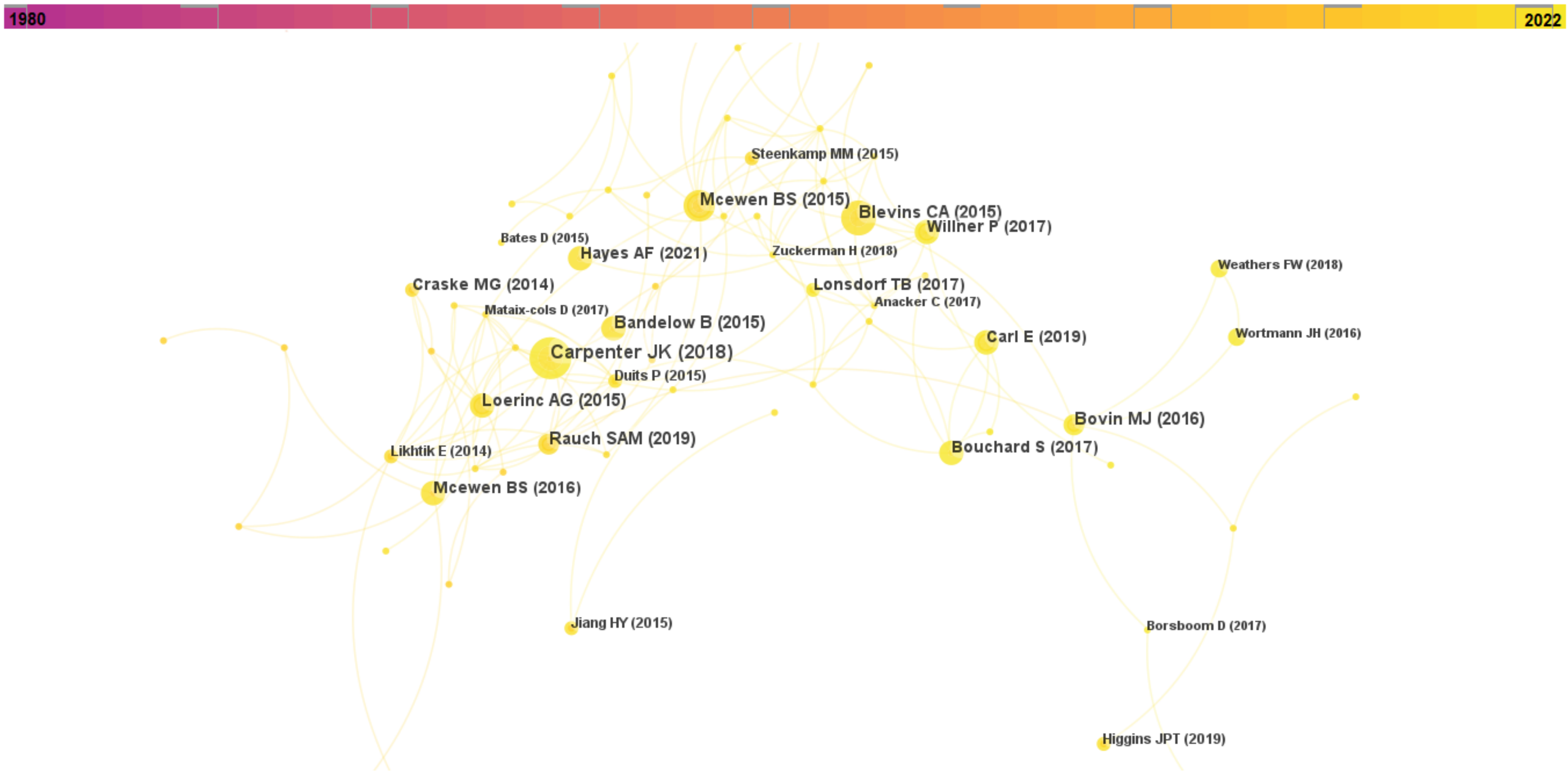

**Cluster 14 • "Cognitive Therapy"**: psychiatric status rating scales (12.55, 0.001); dysthymia (12.55, 0.001); imipramine (12.55, 0.001); cognitive behavioral (12.55, 0.001); cognitive therapy (11.55, 0.001)

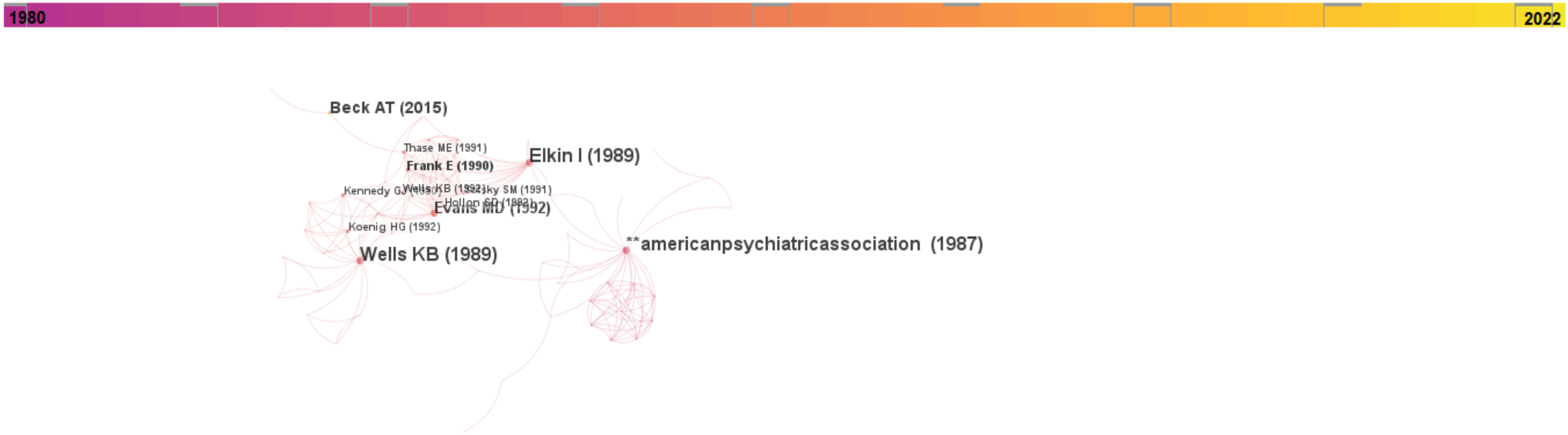

**Cluster 15 • "Gut-brain Axis"**: gut microbiota (23.09, 1.0E-4); microbiota-gut-brain axis (21.1, 1.0E-4); microbiota (21.1, 1.0E-4); early-life stress (21.1, 1.0E-4); bdnf (18.37, 1.0E-4)

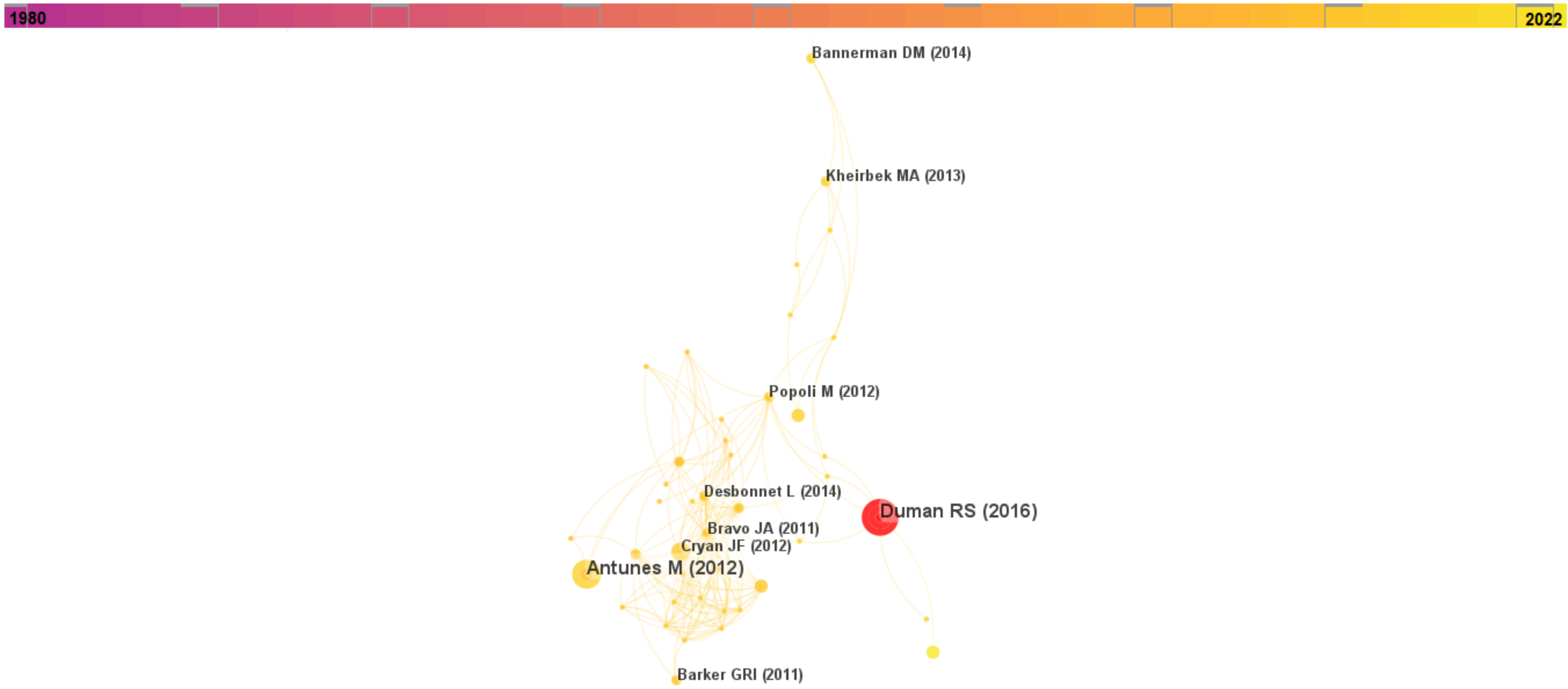

**Cluster 16 • “Sleep Apnea”**: sleep apnea (19.55, 1.0E-4); sleep-disordered breathing (19.33, 1.0E-4); hypoxia (13.82, 0.001); sleepiness (12.65, 0.001); excessive daytime somnolence (9.65, 0.005)

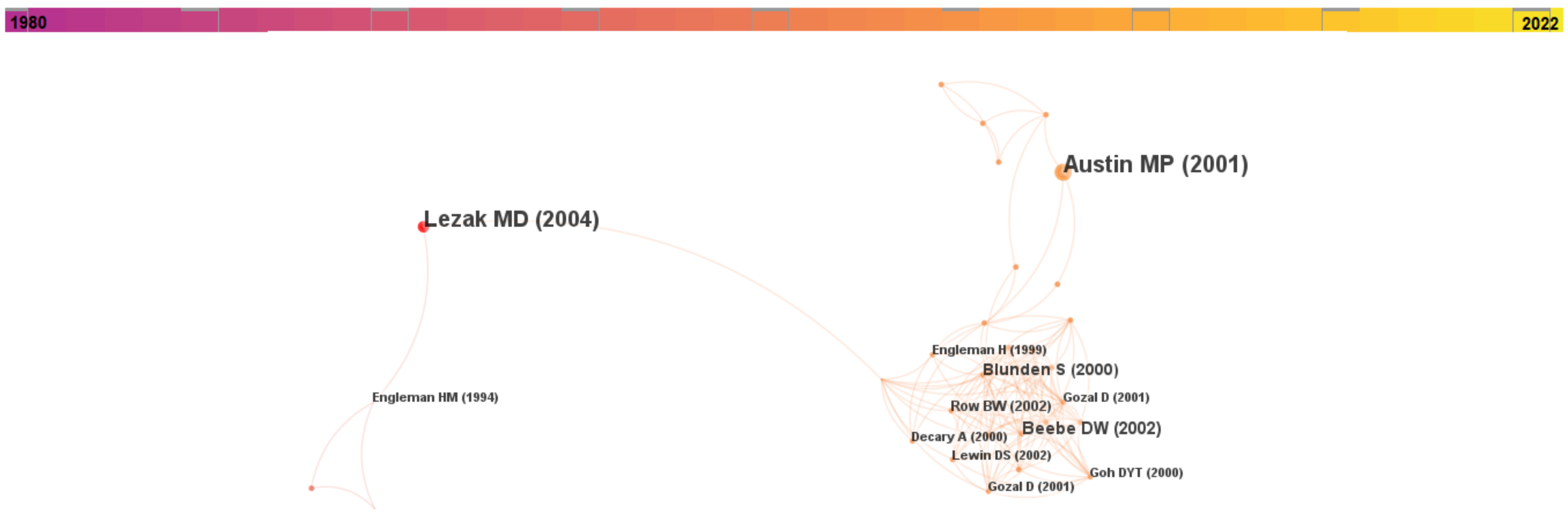

**Supplementary Figure 3.** The co-citation reference network (1980 to 2022) (A), and the co-citation reference network (2020 to 2022) (B) and its corresponding clusters (C) with the original, autogenerated cluster labels by CiteSpace prior to re-labelling

# A

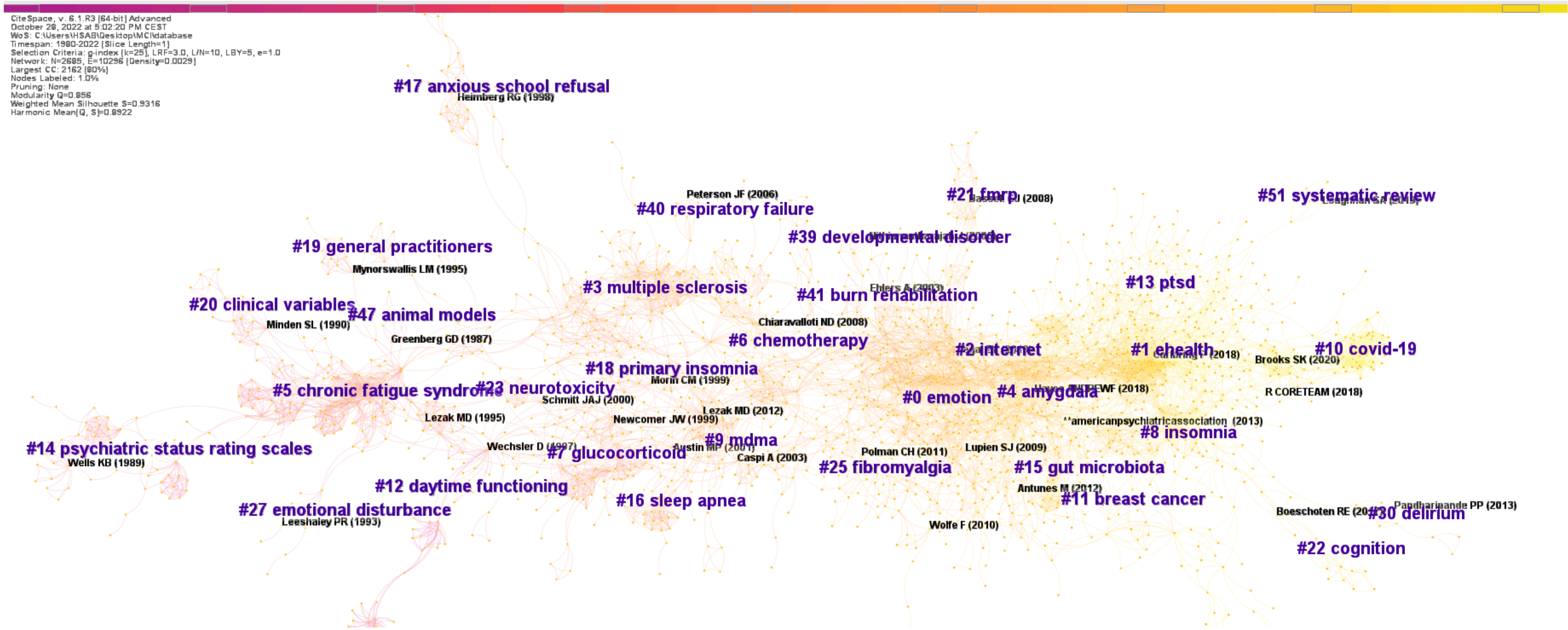

.

B

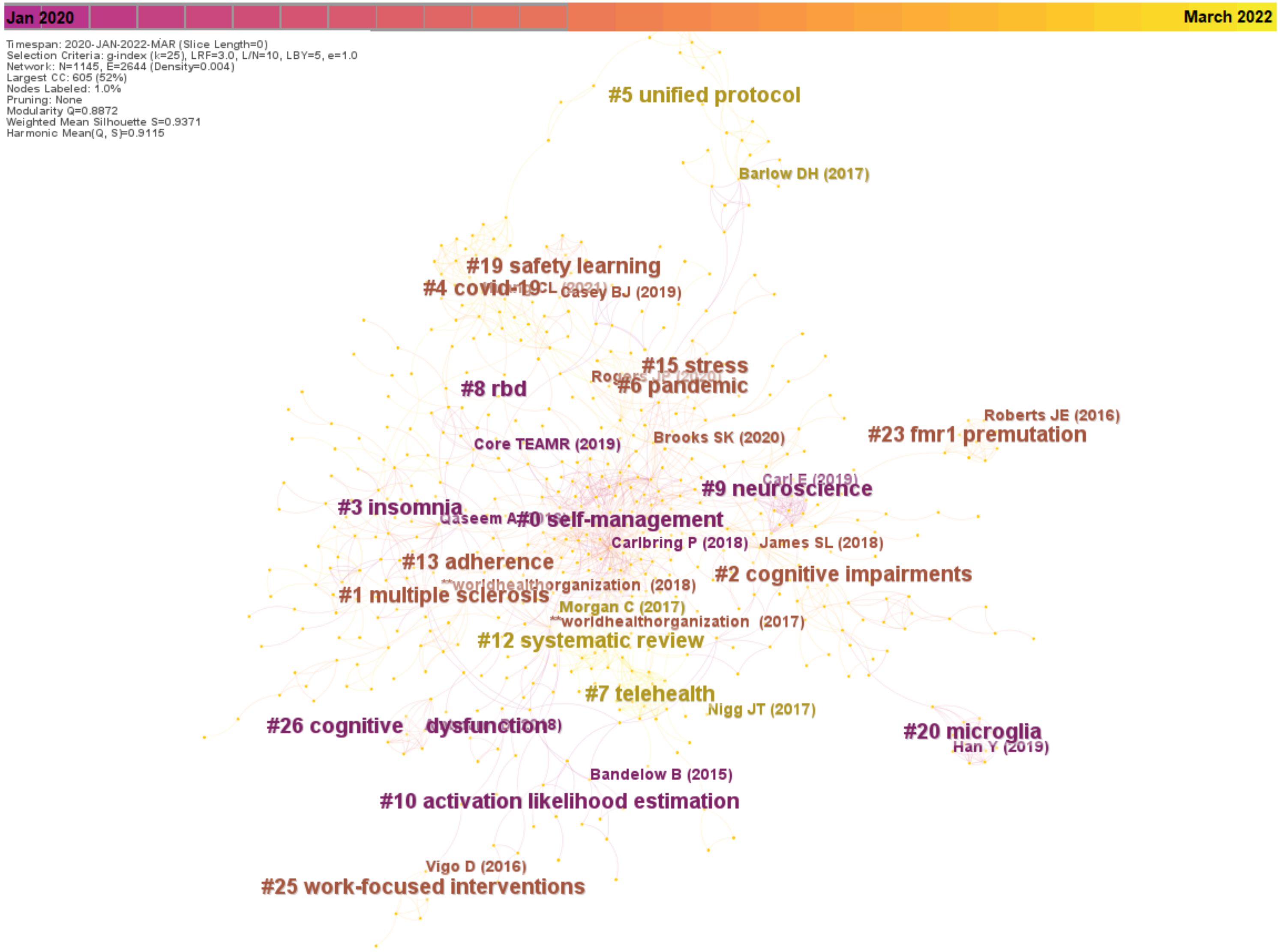

Jan 2020
March 2022
Timespan: 2020-JAN-2022-MAR (Slice Length=0)
Selection Criteria: g-index (k=25), LRF=3.0, L/N=10, LBY=5, e=1.0
Network: N=1145, E=2644 (Density=0.004)
Largest CC: 605 (52%)
Nodes Labeled: 1.0%
Pruning: None
Modularity Q=0.8872
Weighted Mean Silhouette S=0.9371
Harmonic Mean(Q, S)=0.9115
#5 unified protocol
Barlow DH (2017)
#19 safety learning
#4 covid-19
Casey BJ (2019)
#15 stress
#6 pandemic
#8 rbd
Roberts JE (2016)
#23 fmr1 premutation
Core TEAMR (2019)
Brooks SK (2020)
#9 neuroscience
#3 insomnia
#0 self-management
Carlbring P (2018)
James SL (2018)
#13 adherence
#2 cognitive impairments
**worldhealthorganization (2018)
#1 multiple sclerosis
Morgan C (2017)
**worldhealthorganization (2017)
#12 systematic review
#7 telehealth
Nigg JT (2017)
#26 cognitive dysfunction
#20 microglia
Han Y (2019)
Bandelow B (2015)
#10 activation likelihood estimation
Vigo D (2016)
#25 work-focused interventions

**C**

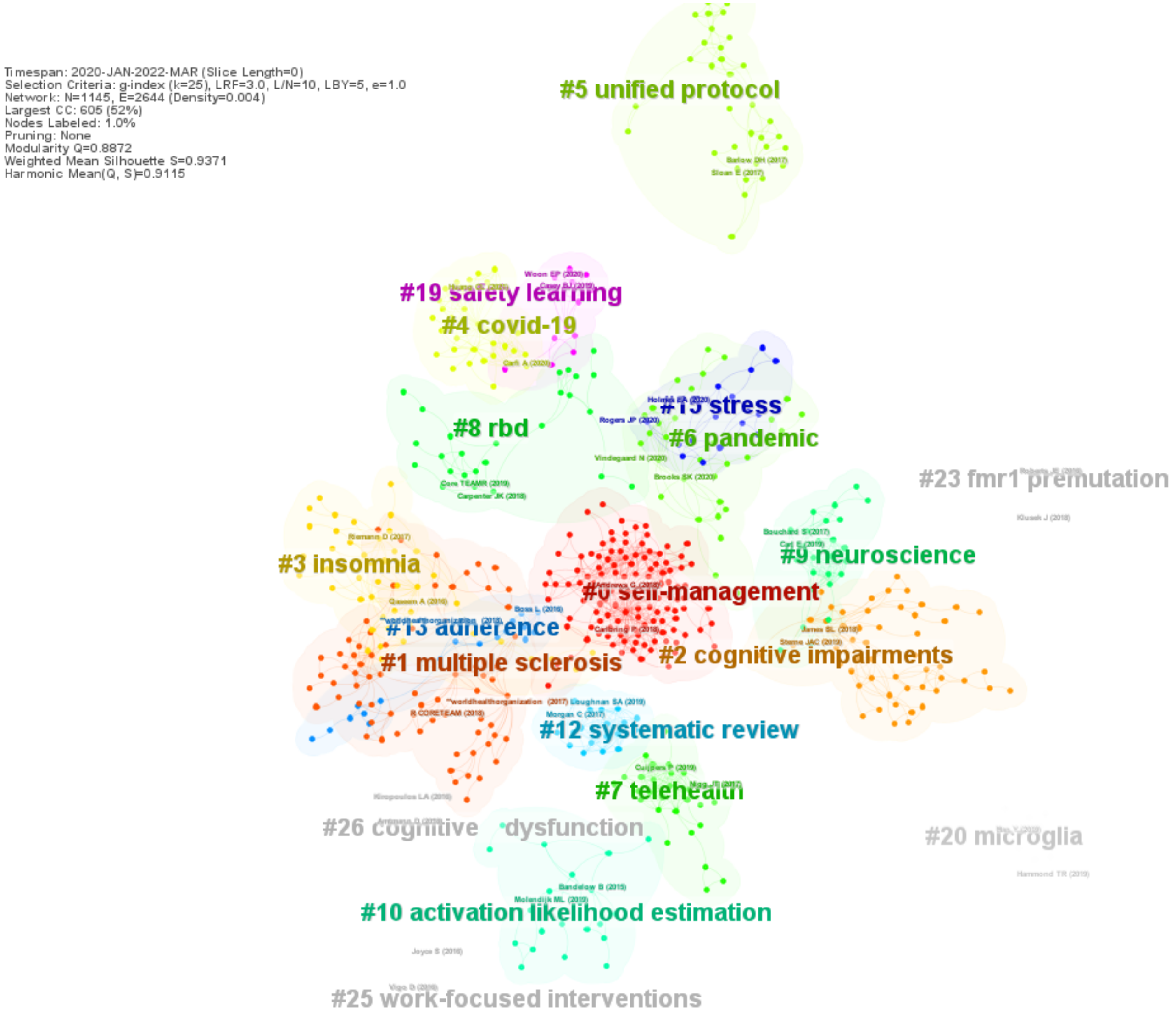

Timespan: 2020-JAN-2022-MAR (Slice Length=0)
Selection Criteria: g-index (k=25), LRF=3.0, L/N=10, LBY=5, e=1.0
Network: N=1145, E=2644 (Density=0.004)
Largest CC: 605 (52%)
Nodes Labeled: 1.0%
Pruning: None
Modularity Q=0.8872
Weighted Mean Silhouette S=0.9371
Harmonic Mean(Q, S)=0.9115
#5 unified protocol
#19 safety learning
#4 covid-19
#15 stress
#6 pandemic
#8 rbd
#23 fmr1 premutation
#9 neuroscience
#3 insomnia
#0 self-management
#13 adherence
#2 cognitive impairments
#1 multiple sclerosis
#12 systematic review
#7 telehealth
#26 cognitive dysfunction
#20 microglia
#10 activation likelihood estimation
#25 work-focused interventions

**Supplementary Table 1**. Detailed information on the clusters in the co-cited reference networks (1980-2022 network and 2020-2022 network)

| **Information of the largest clusters in the co-cited reference network (1980 to 2022)** | | | | | |
|---|---|---|---|---|---|
| **Cluster #** | **Cluster Size** | **Silhouette Score** | **Mean Year** | **Cluster Label and Top Five Extracted Terms (Log-likelihood ratio; p-value)** | **References with Highest Centrality** |
| 0 | 248 | 0.888 | 2011 | **Cognition Disorders**<br>emotion (46.96, 1.0E-4); fmri (37.17, 1.0E-4); prefrontal cortex (30.76, 1.0E-4); insomnia (30.31, 1.0E-4); attention bias (29.64, 1.0E-4) | Lupien SJ, McEwen BS, Gunnar MR, Heim C. Effects of stress throughout the lifespan on the brain, behaviour and cognition. Nature Reviews Neuroscience. 2009;10(6):434–45. doi:10.1038/nrn2639 |
| 1 | 241 | 0.889 | 2018 | **Internet-derived Cognitive Behavioral Therapy**<br>ehealth (59.08, 1.0E-4); telemedicine (50.5, 1.0E-4); mhealth (46.61, 1.0E-4); mental health (35.34, 1.0E-4); e-mental health (31.04, 1.0E-4) | Carlbring P, Andersson G, Cuijpers P, Riper H, Hedman-Lagerlöf E. Internet-based vs. face-to-face cognitive behavior therapy for psychiatric and somatic disorders: An updated systematic review and meta-analysis. Cognitive Behaviour Therapy. 2017;47(1):1–18. doi:10.1080/16506073.2017.1401115 |
| 2 | 167 | 0.867 | 2011 | **Mindfulness**<br>internet (42.85, 1.0E-4); depression (30.62, 1.0E-4); cognitive therapy (29.06, 1.0E-4); mindfulness (27.75, 1.0E-4); mindfulness-based cognitive therapy (18.37, 1.0E-4) | Segal ZV, G. WJM, Teasdale JD. Mindfulness-based cognitive therapy for Depression. 2nd ed. New York: Guilford Press; 2013. |
| 3 | 161 | 0.937 | 2004 | **Multiple Sclerosis**<br>multiple sclerosis (132.53, 1.0E-4); disability (23.03, 1.0E-4); cognition (18.17, 1.0E-4); fatigue (15.48, 1.0E-4); longitudinal study (13.62, 0.001) | Chiaravalloti ND, DeLuca J. Cognitive impairment in multiple sclerosis. The Lancet Neurology. 2008;7(12):1139–51. doi:10.1016/s1474-4422(08)70259-x |
| 4 | 144 | 0.879 | 2015 | **Amygdala**<br>amygdala (38.33, 1.0E-4); fmri (32.39, 1.0E-4); social anxiety disorder (28.99, 1.0E-4); emotion regulation (27.13, 1.0E-4); chronic pain (25.83, 1.0E-4) | Etkin A, Egner T, Kalisch R. Emotional processing in anterior cingulate and medial prefrontal cortex. Trends in Cognitive Sciences. 2011;15(2):85–93. doi:10.1016/j.tics.2010.11.004 |
| 5 | 128 | 0.947 | 1995 | **Chronic Fatigue Syndrome**<br>chronic fatigue syndrome (83.63, 1.0E-4); affective disorders (11.46, 0.001); cognition disorders (11.46, 0.001); cognitive deficits (9.86, 0.005); neuropsychological impairments (9.05, 0.005) | Lezak MD. Neuropsychological assessment. 3rd ed. New York: Oxford University Press; 1995. |
| 6 | 111 | 0.937 | 2007 | **Neuropsychology**<br>chemotherapy (24.15, 1.0E-4); anxiety (20.11, 1.0E-4); tbi (16.73, 1.0E-4); cognitive function (15.7, 1.0E-4); rehabilitation (14.67, 0.001) | Lezak MD. Neuropsychological assessment. 5th ed. Oxford: Oxford University Press; 2012. |
| 7 | 106 | 0.959 | 2001 | **Glucocorticoid**<br>glucocorticoid (23.58, 1.0E-4); hippocampus (17.8, 1.0E-4); glucocorticoids (16.93, 1.0E-4); human (9.1, 0.005); anxiety (8.59, 0.005) | Newcomer JW. Decreased memory performance in healthy humans induced by stress-level cortisol treatment. Archives of General Psychiatry. 1999;56(6):527–33. doi:10.1001/archpsyc.56.6.527 |

| | | | | | |
|---|---|---|---|---|---|
| 8 | 102 | 0.922 | 2017 | **Insomnia**<br><br>insomnia (273.46, 1.0E-4); sleep (112.28, 1.0E-4); cbt-i (54.89, 1.0E-4); chronic insomnia (24.07, 1.0E-4); sleep disorders (20.01, 1.0E-4) | Qaseem A, Kansagara D, Forciea MA, Cooke M, Denberg TD. Management of chronic insomnia disorder in adults: A clinical practice guideline from the American College of Physicians. Annals of Internal Medicine. 2016;165(2):125. doi:10.7326/m15-2175 |
| 9 | 92 | 0.963 | 2005 | **Cortico-limbic Pathway**<br><br>mdma (33.65, 1.0E-4); ecstasy (24.42, 1.0E-4); serotonin (23.27, 1.0E-4); tryptophan (10.84, 0.001); inhibition (9.16, 0.005) | Caspi A, Sugden K, Moffitt TE, Taylor A, Craig IW, Harrington H, et al. Influence of life stress on depression: Moderation by a polymorphism in the 5-htt gene. Science. 2003;301(5631):386–9. doi:10.1126/science.1083968 |
| 10 | 88 | 0.984 | 2021 | **COVID-19**<br><br>covid-19 (174.61, 1.0E-4); sars-cov-2 (57.73, 1.0E-4); long covid (44.86, 1.0E-4); covid-19 pandemic (20.67, 1.0E-4); post-covid-19 (19.18, 1.0E-4) | Brooks SK, Webster RK, Smith LE, Woodland L, Wessely S, Greenberg N, et al. The psychological impact of quarantine and how to reduce it: Rapid review of the evidence. The Lancet. 2020 Feb 26;395(10227):912–20. doi:10.1016/s0140-6736(20)30460-8 |
| 11 | 86 | 0.944 | 2017 | **Cancer**<br><br>breast cancer (51.63, 1.0E-4); cancer (49.22, 1.0E-4); chemotherapy (30.78, 1.0E-4); chemobrain (30.29, 1.0E-4); oncology (28.92, 1.0E-4) | Polman CH, Reingold SC, Banwell B, Clanet M, Cohen JA, Filippi M, et al. Diagnostic criteria for multiple sclerosis: 2010 revisions to the McDonald Criteria. Annals of Neurology. 2011;69(2):292–302. doi:10.1002/ana.22366<br><br>Janelsins MC, Heckler CE, Peppone LJ, Kamen C, Mustian KM, Mohile SG, et al. Cognitive complaints in survivors of breast cancer after chemotherapy compared with age-matched controls: An analysis from a nationwide, Multicenter, prospective longitudinal study. Journal of Clinical Oncology. 2017;35(5):506–14. doi:10.1200/jco.2016.68.5826 |
| 12 | 75 | 0.989 | 1991 | **Social Phobia/Delusion**<br><br>daytime functioning (20.02, 1.0E-4); sleepiness (13.33, 0.001); behaviour therapy (9.99, 0.005); wake (9.99, 0.005); maois (9.99, 0.005) | Gelernter CS. Cognitive-behavioral and pharmacological treatments of social phobia. Archives of General Psychiatry. 1991;48(10):938. doi:10.1001/archpsyc.1991.01810340070009 |
| 13 | 69 | 0.933 | 2019 | **Post-traumatic Stress Disorder**<br><br>ptsd (40.69, 1.0E-4); exposure therapy (39.88, 1.0E-4); posttraumatic stress disorder (31.58, 1.0E-4); veterans (24.99, 1.0E-4); virtual reality (23.24, 1.0E-4) | Carpenter JK, Andrews LA, Witcraft SM, Powers MB, Smits JA, Hofmann SG. Cognitive behavioral therapy for anxiety and related disorders: A meta-analysis of randomized placebo-controlled trials. Depression and Anxiety. 2018;35(6):502–14. doi:10.1002/da.22728 |
| 14 | 56 | 0.995 | 1992 | **Cognitive Therapy**<br><br>psychiatric status rating scales (12.55, 0.001); dysthymia (12.55, 0.001); imipramine (12.55, 0.001); cognitive behavioral (12.55, 0.001); cognitive therapy (11.55, 0.001) | Elkin I. National Institute of Mental Health Treatment of Depression Collaborative Research Program. Archives of General Psychiatry. 1989;46(11):971. doi:10.1001/archpsyc.1989.01810110013002 |
| 15 | 43 | 0.973 | 2015 | **Gut-brain Axis**<br><br>gut microbiota (23.09, 1.0E-4); microbiota-gut-brain axis (21.1, 1.0E-4); microbiota (21.1, 1.0E-4); early-life stress (21.1, 1.0E-4); bdnf (18.37, 1.0E-4) | Antunes M, Biala G. The novel Object recognition memory: Neurobiology, test procedure, and its modifications. Cognitive Processing. 2011;13(2):93–110. doi:10.1007/s10339-011-0430-z<br><br>Bravo JA, Forsythe P, Chew MV, Escaravage E, Savignac HM, Dinan TG, et al. Ingestion of lactobacillus strain regulates emotional behavior and central GABA receptor expression in a mouse via the vagus nerve. Proceedings of the National Academy of Sciences. 2011;108(38):16050–5. doi:10.1073/pnas.1102999108 |

| | | | | | |
|---|---|---|---|---|---|
| 16 | 31 | 0.986 | 2002 | **Sleep Apnea**<br><br>sleep apnea (19.55, 1.0E-4); sleep-disordered breathing (19.33, 1.0E-4); hypoxia (13.82, 0.001); sleepiness (12.65, 0.001); excessive daytime somnolence (9.65, 0.005) | Austin M-P, Mitchell P, Goodwin GM. Cognitive deficits in depression: Possible implications for functional neuropathology. British Journal of Psychiatry. 2001;178(3):200–6. doi:10.1192/bjp.178.3.200<br><br>Beebe DW, Gozal D. Obstructive sleep apnea and the prefrontal cortex: Towards a comprehensive model linking nocturnal upper airway obstruction to daytime cognitive and behavioral deficits. Journal of Sleep Research. 2002;11(1):1–16. doi:10.1046/j.1365-2869.2002.00289.x |
| 17 | 29 | 0.999 | 2002 | **Diagnosis**<br><br>anxious school refusal (12.03, 0.001); hispanic (12.03, 0.001); childhood anxiety disorders (12.03, 0.001); family functioning (12.03, 0.001); pharmacological treatment (12.03, 0.001) | Heimberg RG, Liebowitz MR, Hope DA, Schneier FR, Holt CS, Welkowitz LA, et al. Cognitive Behavioral Group therapy vs Phenelzine therapy for social phobia. Archives of General Psychiatry. 1998;55(12):1133. doi:10.1001/archpsyc.55.12.1133 |
| 18 | 28 | 0.998 | 2002 | **Primary Chronic Insomnia Treatment**<br><br>primary insomnia (21.17, 1.0E-4); self-medication (10.56, 0.005); measures (10.56, 0.005); primary care (10.56, 0.005); sleep hygiene (10.56, 0.005) | Smith MT, Perlis ML, Park A, Smith MS, Pennington J, Giles DE, et al. Comparative meta-analysis of pharmacotherapy and behavior therapy for persistent insomnia. American Journal of Psychiatry. 2002;159(1):5–11. doi:10.1176/appi.ajp.159.1.5 |
| 19 | 22 | 0.998 | 1999 | **Depression**<br><br>general practitioners (12.55, 0.001); psychological management (12.55, 0.001); outcome prediction (12.55, 0.001); primary care review (12.55, 0.001); psychological therapy (12.55, 0.001) | Mynors-Wallis LM, Gath DH, Lloyd-Thomas AR, Tomlinson D. Randomised controlled trial comparing problem solving treatment with amitriptyline and placebo for major depression in primary care. BMJ. 1995;310(6977):441–5. doi:10.1136/bmj.310.6977.441 |
| 20 | 19 | 0.997 | 1993 | **Cognitive Dysfunction in Multiple Sclerosis**<br><br>clinical variables (15.25, 1.0E-4); neuropsychological performance (15.25, 1.0E-4); multiple sclerosis (4.41, 0.05); depression (1.56, 0.5); anxiety (0.24, 1.0) | Minden SL, Schiffer RB. Affective disorders in multiple sclerosis review and recommendations for clinical research. Archives of Neurology. 1990;47(1):98–104. doi:10.1001/archneur.1990.00530010124031 |
| 21 | 18 | 0.999 | 2009 | **Fragile X Syndrome**<br><br>fmrp (19.13, 1.0E-4); synapse (19.13, 1.0E-4); autism (13.92, 0.001); fragile x syndrome (12.45, 0.001); mental retardation (9.54, 0.005) | Bassell GJ, Warren ST. Fragile X syndrome: Loss of local mrna regulation alters synaptic development and function. Neuron. 2008;60(2):201–14. doi:10.1016/j.neuron.2008.10.004 |
| 22 | 17 | 0.995 | 2019 | **Cognition in MS**<br><br>multiple sclerosis (102.08, 1.0E-4); cognition (31.81, 1.0E-4); cognitive impairment (20.77, 1.0E-4); cognitive rehabilitation (18.15, 1.0E-4); neuropsychology (15.61, 1.0E-4) | Boeschoten RE, Braamse AMJ, Beekman ATF, Cuijpers P, van Oppen P, Dekker J, et al. Prevalence of depression and anxiety in multiple sclerosis: A systematic review and meta-analysis. Journal of the Neurological Sciences. 2017;372:331–41. doi:10.1016/j.jns.2016.11.067 |
| 23 | 15 | 0.995 | 2000 | **MDMA**<br><br>mdma (21.44, 1.0E-4); neurotoxicity (17.99, 1.0E-4); ecstasy (14.03, 0.001); (s)-mde (11.71, 0.001); former chronic (11.71, 0.001) | Curran HV, Travill RA. Mood and cognitive effects of 6 3,4-methylenedioxymethamphetamine (MDMA, 'ecstasy'): Week-end "high" followed by mid-week low. Addiction. 1997;92(7):821–31. doi:10.1046/j.1360-0443.1997.9278215.x |
| 25 | 13 | 0.996 | 2009 | **Fibromyalgia** | Wolfe F, Clauw DJ, Fitzcharles M-A, Goldenberg DL, Katz RS, Mease P, et al. The American College of Rheumatology preliminary diagnostic criteria for fibromyalgia and measurement of symptom severity. Arthritis Care & Research. 2010;62(5):600–10. doi:10.1002/acr.20140 |

| | | | | | |
|---|---|---|---|---|---|
| | | | | fibromyalgia (44.54, 1.0E-4); pain (13.65, 0.001); stress axis (10.07, 0.005); emotional decision making (10.07, 0.005); postoperative pain (10.07, 0.005) | |
| 27 | 11 | 1 | 1997 | **Emotional Disturbance**<br><br>emotional disturbance (13.72, 0.001); amnesia (13.72, 0.001); neurotransmitters (13.72, 0.001); brain damage (13.72, 0.001); tests of personality and emotional status (13.72, 0.001) | Burt DB, Zembar MJ, Niederehe G. Depression and memory impairment: A meta-analysis of the association, its pattern, and specificity. Psychological Bulletin. 1995;117(2):285–305. doi:10.1037/0033-2909.117.2.285 |
| 30 | 10 | 1 | 2015 | **Delirium**<br><br>delirium (86.2, 1.0E-4); critical care (43.97, 1.0E-4); sedation (30.21, 1.0E-4); guidelines (26.36, 1.0E-4); abcdef bundle (17.55, 1.0E-4) | Pandharipande PP, Girard TD, Jackson JC, Morandi A, Thompson JL, Pun BT, et al. Long-term cognitive impairment after critical illness. New England Journal of Medicine. 2013;369(14):1306–16. doi:10.1056/nejmoa1301372 |
| 39 | 7 | 1 | 2010 | **Rett Syndrome**<br><br>developmental disorder (11.61, 0.001); synaptopathy (11.61, 0.001); neurodevelopmental disorders (11.61, 0.001); metabolic compromise (11.61, 0.001); ultrasound vocalizations (11.61, 0.001) | Nithianantharajah J, Hannan AJ. Enriched environments, experience-dependent plasticity and disorders of the nervous system. Nature Reviews Neuroscience. 2006;7(9):697–709. doi:10.1038/nrn1970<br><br>Kondo M, Gray LJ, Pelka GJ, Christodoulou J, Tam PP, Hannan AJ. Environmental enrichment ameliorates a motor coordination deficit in a mouse model of Rett syndrome mecp2 gene dosage effects and BDNF expression. European Journal of Neuroscience. 2008;27(12):3342–50. doi:10.1111/j.1460-9568.2008.06305.x |
| 40 | 7 | 1 | 2007 | **ICU Patients**<br><br>delirium (12.69, 0.001); respiratory failure (12.4, 0.001); analgesia (12.4, 0.001); posttraumatic stress disorder (12.4, 0.001); protocols (12.4, 0.001) | Peterson JF, Pun BT, Dittus RS, Thomason JW, Jackson JC, Shintani AK, et al. Delirium and its motoric subtypes: A study of 614 critically ill patients. Journal of the American Geriatrics Society. 2006;54(3):479–84. doi:10.1111/j.1532-5415.2005.00621.x |
| 41 | 7 | 0.997 | 2006 | **Psychotherapy for PTSD**<br><br>burn rehabilitation (14.01, 0.001); cognitive-behavioural therapy (14.01, 0.001); natural recovery (14.01, 0.001); early intervention (11.24, 0.001); outcomes (10.2, 0.005) | Ehlers A, Clark DM, Hackmann A, McManus F, Fennell M, Herbert C, et al. A randomized controlled trial of cognitive therapy, a self-help booklet, and repeated assessments as early interventions for posttraumatic stress disorder. Archives of General Psychiatry. 2003;60(10):1024. doi:10.1001/archpsyc.60.10.1024 |
| 47 | 6 | 0.998 | 1992 | **Geriatric Sleep Apnea**<br><br>animal models (NaN, 1.0); default mode network (dmn) (NaN, 1.0); pad bundle (NaN, 1.0); prolonged exposure therapy (NaN, 1.0); benzodiazepine receptor agonist (NaN, 1.0) | Walker A. Markers of HIV infection in the Concorde trial. Concorde Co-ordinating Committee. QJM. 1998;91(6):423–38. doi:10.1093/qjmed/91.6.423<br><br>1. Berry DT, Phillips BA, Cook YR, Schmitt FA, Gilmore RL, Patel R, et al. Sleep-disordered breathing in healthy aged persons: Possible daytime sequelae. Journal of Gerontology. 1987;42(6):620–6. doi:10.1093/geronj/42.6.620 |
| 51 | 5 | 0.999 | 2022 | **Pregnancy**<br><br>systematic review (18.19, 1.0E-4); digital interventions (17.26, 1.0E-4); pregnancy (12.42, 0.001); digital cognitive behavioral therapy (11.35, 0.001); antenatal (11.35, 0.001) | Loughnan SA, Joubert AE, Grierson A, Andrews G, Newby JM. Internet-delivered psychological interventions for clinical anxiety and depression in perinatal women: A systematic review and meta-analysis. Archives of Women's Mental Health. 2019;22(6):737–50. doi:10.1007/s00737-019-00961-9 |

| Information of the largest clusters in the co-cited reference network (2020 to 2022) | | | | | |
|---|---|---|---|---|---|
| **Cluster #** | **Cluster Size** | **Silhouette Score** | **Mean Year** | **Cluster Label and Top Five Extracted Terms (Log-likelihood ratio; p-value)** | **References with Highest Centrality** |
| 0 | 123 | 0.842 | 2020 | **Internet-based Cognitive Behavioral Therapy**<br><br>self-management (10.24, 0.005); telemedicine (7.72, 0.01); covid-19 (6.88, 0.01); cognitive behavior therapy (6.83, 0.01); ehealth (6.83, 0.01) | Carlbring P, Andersson G, Cuijpers P, Riper H, Hedman-Lagerlöf E. Internet-based vs. face-to-face cognitive behavior therapy for psychiatric and somatic disorders: An updated systematic review and meta-analysis. Cognitive Behaviour Therapy. 2017;47(1):1–18. doi:10.1080/16506073.2017.1401115 |
| 1 | 71 | 0.959 | 2020 | **Multiple Sclerosis**<br><br>multiple sclerosis (55.01, 1.0E-4); cognition (8.49, 0.005); neuropsychology (7.72, 0.01); posttraumatic stress disorder (7.72, 0.01); employment (7.72, 0.01) | Benedict RH, DeLuca J, Phillips G, LaRocca N, Hudson LD, Rudick R. Validity of the symbol digit modalities test as a cognition performance outcome measure for multiple sclerosis. Multiple Sclerosis Journal. 2017;23(5):721–33. doi:10.1177/1352458517690821 |
| 2 | 53 | 0.947 | 2021 | **Risk**<br><br>cognitive impairments (9.57, 0.005); antidepressant (9.57, 0.005); physical activity (9.57, 0.005); hippocampus (5.94, 0.05); meta-analysis (5.58, 0.05) | James SL, Abate D, Abate KH, Abay SM, Abbafati C, Abbasi N, et al. Global, regional, and national incidence, prevalence, and years lived with disability for 354 diseases and injuries for 195 countries and territories, 1990–2017: A systematic analysis for the global burden of disease study 2017. The Lancet. 2018;392(10159):1789–858. doi:10.1016/s0140-6736(18)32279-7 |
| 3 | 49 | 0.942 | 2020 | **Insomnia**<br><br>insomnia (38.38, 1.0E-4); sleep (18.65, 1.0E-4); cbt-i (14.26, 0.001); sleep disorders (9.49, 0.005); traumatic brain injury (9.49, 0.005) | Riemann D, Baglioni C, Bassetti C, Bjorvatn B, Dolenc Groselj L, Ellis JG, et al. European guideline for the diagnosis and treatment of insomnia. Journal of Sleep Research. 2017;26(6):675–700. doi:10.1111/jsr.12594 |
| 4 | 36 | 0.949 | 2021 | **COVID-19**<br><br>covid-19 (22.48, 1.0E-4); functional decline (6.26, 0.05); post-covid-19 syndrome (6.26, 0.05); post-acute covid syndrome (6.26, 0.05); screening (6.26, 0.05) | Carfì A, Bernabei R, Landi F. Persistent symptoms in patients after acute COVID-19. JAMA. 2020;324(6):603. doi:10.1001/jama.2020.12603 |
| 5 | 35 | 0.988 | 2021 | **Transdiagnostic**<br><br>unified protocol (20.71, 1.0E-4); transdiagnostic (6.65, 0.01); mood disorders (6.65, 0.01); animal models (5.14, 0.05); anterior cingulate cortex (5.14, 0.05) | Barlow DH, Farchione TJ, Bullis JR, Gallagher MW, Murray-Latin H, Sauer-Zavala S, et al. The unified protocol for transdiagnostic treatment of emotional disorders compared with diagnosis-specific protocols for anxiety disorders. JAMA Psychiatry. 2017;74(9):875. doi:10.1001/jamapsychiatry.2017.2164 |
| 6 | 34 | 0.912 | 2021 | **COVID-19 & Mental Health**<br><br>covid-19 (15.58, 1.0E-4); pandemic (13.75, 0.001); loneliness (12.06, 0.001); college students (12.06, 0.001); trauma (6.01, 0.05) | Brooks SK, Webster RK, Smith LE, Woodland L, Wessely S, Greenberg N, et al. The psychological impact of quarantine and how to reduce it: Rapid review of the evidence. The Lancet. 2020 Feb 26;395(10227):912–20. doi:10.1016/s0140-6736(20)30460-8 |
| 7 | 27 | 0.977 | 2021 | **Psychotherapy**<br><br>telehealth (9.1, 0.005); online self-help (6.39, 0.05); group counseling (6.39, 0.05); caregiver (6.39, 0.05); family (6.39, 0.05) | US Preventive Services Task Force. Interventions to Prevent Perinatal Depression: US Preventive Services Task Force Recommendation Statement. JAMA. 2019;321(6):580. doi:10.1001/jama.2019.0007 |
| 8 | 26 | 0.935 | 2020 | **Cognitive Emotion Regulation** | Carpenter JK, Andrews LA, Witcraft SM, Powers MB, Smits JA, Hofmann SG. Cognitive behavioral therapy for anxiety and related disorders: A meta- |

| | | | | | |
|---|---|---|---|---|---|
| | | | | rbd (12.3, 0.001); cognitive emotion regulation strategies (12.3, 0.001); rem sleep behavior disorder (12.3, 0.001); emotion regulation (8.58, 0.005); nightmares (6.94, 0.01) | analysis of randomized placebo-controlled trials. Depression and Anxiety. 2018;35(6):502–14. doi:10.1002/da.22728 |
| 9 | 25 | 0.999 | 2020 | **Virtual Reality Exposure Therapy**<br><br>neuroscience (7.26, 0.01); virtual social worlds (7.26, 0.01); extinction (7.26, 0.01); virtual reality exposure therapy (7.26, 0.01); conditioning (7.26, 0.01) | Carl E, Stein AT, Levihn-Coon A, Pogue JR, Rothbaum B, Emmelkamp P, et al. Virtual reality exposure therapy for anxiety and related disorders: A meta-analysis of randomized controlled trials. Journal of Anxiety Disorders. 2019;61:27–36. doi:10.1016/j.janxdis.2018.08.003 |
| 10 | 21 | 0.947 | 2020 | **Reward/Learning**<br><br>activation likelihood estimation (6.3, 0.05); psychological therapies (6.3, 0.05); hrv (6.3, 0.05); ventral tegmental area (6.3, 0.05); computational neuroscience (6.3, 0.05) | Molendijk ML, de Kloet ER. Coping with the forced swim stressor: Current state-of-the-art. Behavioural Brain Research. 2019;364:1–10. doi:10.1016/j.bbr.2019.02.005 |
| 12 | 19 | 0.966 | 2021 | **Electronic Health**<br><br>systematic review (20.98, 1.0E-4); pregnancy (13.92, 0.001); digital interventions (10.17, 0.005); cognitive behavior therapy (6.93, 0.01); electronic health (6.93, 0.01) | Loughnan SA, Sie A, Hobbs MJ, Joubert AE, Smith J, Haskelberg H, et al. A randomized controlled trial of 'mumentum pregnancy': Internet-delivered cognitive behavioral therapy program for antenatal anxiety and depression. Journal of Affective Disorders. 2019;243:381–90. doi:10.1016/j.jad.2018.09.057 |
| 13 | 19 | 0.99 | 2021 | **Adherence**<br><br>adherence (12.06, 0.001); burnout (8.34, 0.005); medically unexplained symptoms (6.01, 0.05); self-help (6.01, 0.05); mental disorder (6.01, 0.05) | Boß L, Lehr D, Reis D, Vis C, Riper H, Berking M, et al. Reliability and validity of assessing user satisfaction with web-based health interventions. Journal of Medical Internet Research. 2016;18(8). doi:10.2196/jmir.5952 |
| 15 | 17 | 0.989 | 2021 | **Stress**<br><br>stress (9.88, 0.005); cognitive dysfunction (7.58, 0.01); behavioral outcomes (7.58, 0.01); network activity (7.58, 0.01); healthcare workers (7.58, 0.01) | Holmes EA, O'Connor RC, Perry VH, Tracey I, Wessely S, Arseneault L, et al. Multidisciplinary research priorities for the COVID-19 pandemic: A call for action for mental health science. The Lancet Psychiatry. 2020;7(6):547–60. doi:10.1016/s2215-0366(20)30168-1 |
| 19 | 13 | 0.982 | 2021 | **Neural Circuitry**<br><br>safety learning (8.41, 0.005); lithium (8.41, 0.005); conditioned inhibition (8.41, 0.005); esketamine (8.41, 0.005); kynurenine pathway (8.41, 0.005) | Casey BJ, Heller AS, Gee DG, Cohen AO. Development of the emotional brain. Neuroscience Letters. 2019;693:29–34. doi:10.1016/j.neulet.2017.11.055 |
| 20 | 12 | 1 | 2020 | **Microglia**<br><br>microglia (9.39, 0.005); brain development (7.95, 0.005); maternal sleep deprivation (7.95, 0.005); ppar gamma (7.95, 0.005); brain-derived neurotrophic factor (7.95, 0.005) | Han Y, Zhang L, Wang Q, Zhang D, Zhao Q, Zhang J, et al. Minocycline inhibits microglial activation and alleviates depressive-like behaviors in male adolescent mice subjected to maternal separation. Psychoneuroendocrinology. 2019;107:37–45. doi:10.1016/j.psyneuen.2019.04.021 |
| 23 | 9 | 0.998 | 2021 | **FMR1 Premutation**<br><br>fmr1 premutation (10.89, 0.001); broad autism phenotype (10.89, 0.001); social cognition (7.09, 0.01); latent profile analysis (7.09, 0.01); executive function (6.42, 0.05) | Klusek J, Thurman AJ, Abbeduto L. Maternal pragmatic language difficulties in the FMR1 premutation and the broad autism phenotype: Associations with individual and family outcomes. Journal of Autism and Developmental Disorders. 2021;52(2):835–51. doi:10.1007/s10803-021-04980-3 |

| | | | | | |
|---|---|---|---|---|---|
| 25 | 8 | 0.989 | 2021 | **Mental Health**<br><br>work-focused interventions (9.01, 0.005); depression & mood disorders (9.01, 0.005); public health (9.01, 0.005); metacognitive therapy (9.01, 0.005); health policy (9.01, 0.005) | Vigo D, Thornicroft G, Atun R. Estimating the true global burden of mental illness. The Lancet Psychiatry. 2016;3(2):171–8. doi:10.1016/s2215-0366(15)00505-2 |
| 26 | 8 | 0.999 | 2020 | **Quality of Life**<br><br>cognitive dysfunction (NaN, 1.0); community referral (NaN, 1.0); animal models (NaN, 1.0); glucocorticoid receptor (NaN, 1.0); social phobia (NaN, 1.0) | Barin L, Salmen A, Disanto G, Babačić H, Calabrese P, Chan A, et al. The disease burden of multiple sclerosis from the individual and population perspective: Which symptoms matter most? Multiple Sclerosis and Related Disorders. 2018;25:112–21. doi:10.1016/j.msard.2018.07.013 |

**Supplementary Table 2**. Burstness analysis of references, keywords, authors, institutions, and countries (1980-2022 network, 2020-2022 network, 2017-2022 network, and 2000-2022 network)

### A. Top 25 References with the Strongest Citation Bursts as Ranked by Beginning Year of Burst (1980 to 2022)

| References | Year | Strength | Begin | End | 1980 - 2022 |
|---|---|---|---|---|---|
| Kessler RC, 1994, ARCH GEN PSYCHIAT, V51, P8 | 1994 | 11.2 | 1994 | 1999 | |
| Krupp LB, 1994, ARCH NEUROL-CHICAGO, V51, P705, DOI 10.1001/archneur.1994.00540190089021, DOI | 1994 | 11.34 | 1995 | 1999 | |
| Deluca J, 1995, J NEUROL NEUROSUR PS, V58, P38, DOI 10.1136/jnnp.58.1.38, DOI | 1995 | 13.85 | 1996 | 2000 | |
| Lezak MD, 2012, NEUROPSYCHOLOGICAL A, V0, P0 | 2012 | 31.15 | 2012 | 2011 | |
| Caspi A, 2003, SCIENCE, V301, P386, DOI 10.1126/science.1083968, DOI | 2003 | 10.52 | 2004 | 2008 | |
| Goldapple K, 2004, ARCH GEN PSYCHIAT, V61, P34, DOI 10.1001/archpsyc.61.1.34, DOI | 2004 | 9.78 | 2005 | 2009 | |
| Kessler RC, 2005, ARCH GEN PSYCHIAT, V62, P593, DOI 10.1001/archpsyc.62.6.593, DOI | 2005 | 19.13 | 2006 | 2010 | |
| American ACADEMYOFSLEEPMEDICINE, 2005, INT CLASS SLEEP DIS, V0, P0 | 2005 | 12.37 | 2006 | 2010 | |
| Lupien SJ, 2009, NAT REV NEUROSCI, V10, P434, DOI 10.1038/nrn2639, DOI | 2009 | 18.35 | 2010 | 2014 | |
| Andersson GERHARD, 2009, COGNITIVE BEHAVIOUR THERAPY, V38, P196, DOI 10.1080/16506070903318960, DOI | 2009 | 17.36 | 2010 | 2014 | |
| Fournier JC, 2010, JAMA-J AM MED ASSOC, V303, P47, DOI 10.1001/jama.2009.1943, DOI | 2010 | 13.07 | 2011 | 2015 | |
| Hakamata Y, 2010, BIOL PSYCHIAT, V68, P982, DOI 10.1016/j.biopsych.2010.07.021, DOI | 2010 | 12.58 | 2011 | 2015 | |
| Baglioni C, 2011, J AFFECT DISORDERS, V135, P10, DOI 10.1016/j.jad.2011.01.011, DOI | 2011 | 19.81 | 2012 | 2016 | |
| Segal ZV, 2013, MINDFULNESS BASED CO, V0, P0 | 2013 | 18.23 | 2013 | 2018 | |
| Moriarty O, 2011, PROG NEUROBIOL, V93, P385, DOI 10.1016/j.pneurobio.2011.01.002, DOI | 2011 | 15.65 | 2012 | 2016 | |
| Johansson R, 2012, EXPERT REV NEUROTHER, V12, P861 | 2012 | 14.49 | 2013 | 2017 | |
| Cuijpers P, 2013, CAN J PSYCHIAT, V58, P376, DOI 10.1177/070674371305800702, DOI | 2013 | 20.25 | 2014 | 2018 | |
| Ferrari AJ, 2013, PLOS MED, V10, P0, DOI 10.1371/journal.pmed.1001547, DOI | 2013 | 10.53 | 2014 | 2018 | |
| Barr J, 2013, CRIT CARE MED, V41, P263, DOI 10.1097/CCM.0b013e3182783b72, DOI | 2013 | 10.1 | 2014 | 2018 | |
| James AC, 2015, COCHRANE DB SYST REV, V0, P0 | 2015 | 16.36 | 2015 | 2020 | |
| Buhle JT, 2014, CEREB CORTEX, V24, P2981, DOI 10.1093/cercor/bht154, DOI | 2014 | 15.16 | 2015 | 2019 | |
| Trauer JM, 2015, ANN INTERN MED, V163, P191, DOI 10.7326/M14-2841, DOI | 2015 | 17.51 | 2016 | 2020 | |
| Gilbody S, 2015, BMJ-BRIT MED J, V351, P0, DOI 10.1136/bmj.h5627, DOI | 2015 | 16.39 | 2016 | 2020 | |
| Zachariae R, 2016, SLEEP MED REV, V30, P1, DOI 10.1016/j.smrv.2015.10.004, DOI | 2016 | 18.82 | 2017 | 2022 | |
| Andersson G, 2016, ANNU REV CLIN PSYCHO, V12, P157, DOI 10.1146/annurev-clinpsy-021815-093006, DOI | 2016 | 13.42 | 2017 | 2022 | |

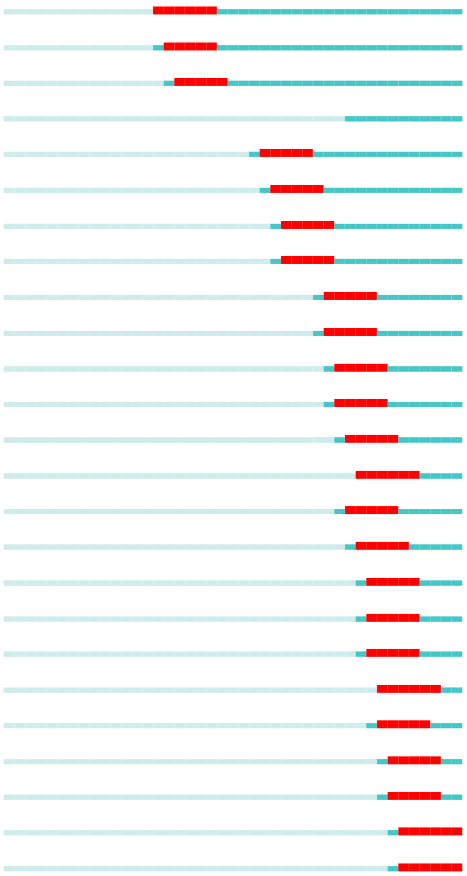

### B. Top 25 References with the Strongest Citation Bursts as Ranked by Burst Strength (1980 to 2022)

| References | Year | Strength | Begin | End | 1980 - 2022 |
|---|---|---|---|---|---|
| Lezak MD, 2012, NEUROPSYCHOLOGICAL A, V0, P0 | 2012 | 31.15 | 2012 | 2011 | |
| Cuijpers P, 2013, CAN J PSYCHIAT, V58, P376, DOI 10.1177/070674371305800702, DOI | 2013 | 20.25 | 2014 | 2018 | |
| Baglioni C, 2011, J AFFECT DISORDERS, V135, P10, DOI 10.1016/j.jad.2011.01.011, DOI | 2011 | 19.81 | 2012 | 2016 | |
| Kessler RC, 2005, ARCH GEN PSYCHIAT, V62, P593, DOI 10.1001/archpsyc.62.6.593, DOI | 2005 | 19.13 | 2006 | 2010 | |
| Zachariae R, 2016, SLEEP MED REV, V30, P1, DOI 10.1016/j.smrv.2015.10.004, DOI | 2016 | 18.82 | 2017 | 2022 | |
| Lupien SJ, 2009, NAT REV NEUROSCI, V10, P434, DOI 10.1038/nrn2639, DOI | 2009 | 18.35 | 2010 | 2014 | |
| Segal ZV, 2013, MINDFULNESS BASED CO, V0, P0 | 2013 | 18.23 | 2013 | 2018 | |
| Trauer JM, 2015, ANN INTERN MED, V163, P191, DOI 10.7326/M14-2841, DOI | 2015 | 17.51 | 2016 | 2020 | |
| Andersson GERHARD, 2009, COGNITIVE BEHAVIOUR THERAPY, V38, P196, DOI 10.1080/16506070903318960, DOI | 2009 | 17.36 | 2010 | 2014 | |
| Gilbody S, 2015, BMJ-BRIT MED J, V351, P0, DOI 10.1136/bmj.h5627, DOI | 2015 | 16.39 | 2016 | 2020 | |
| James AC, 2015, COCHRANE DB SYST REV, V0, P0 | 2015 | 16.36 | 2015 | 2020 | |
| Moriarty O, 2011, PROG NEUROBIOL, V93, P385, DOI 10.1016/j.pneurobio.2011.01.002, DOI | 2011 | 15.65 | 2012 | 2016 | |
| Buhle JT, 2014, CEREB CORTEX, V24, P2981, DOI 10.1093/cercor/bht154, DOI | 2014 | 15.16 | 2015 | 2019 | |
| Johansson R, 2012, EXPERT REV NEUROTHER, V12, P861 | 2012 | 14.49 | 2013 | 2017 | |
| Deluca J, 1995, J NEUROL NEUROSUR PS, V58, P38, DOI 10.1136/jnnp.58.1.38, DOI | 1995 | 13.85 | 1996 | 2000 | |
| Andersson G, 2016, ANNU REV CLIN PSYCHO, V12, P157, DOI 10.1146/annurev-clinpsy-021815-093006, DOI | 2016 | 13.42 | 2017 | 2022 | |
| Fournier JC, 2010, JAMA-J AM MED ASSOC, V303, P47, DOI 10.1001/jama.2009.1943, DOI | 2010 | 13.07 | 2011 | 2015 | |
| Hakamata Y, 2010, BIOL PSYCHIAT, V68, P982, DOI 10.1016/j.biopsych.2010.07.021, DOI | 2010 | 12.58 | 2011 | 2015 | |
| American ACADEMYOFSLEEPMEDICINE, 2005, INT CLASS SLEEP DIS, V0, P0 | 2005 | 12.37 | 2006 | 2010 | |
| Krupp LB, 1994, ARCH NEUROL-CHICAGO, V51, P705, DOI 10.1001/archneur.1994.00540190089021, DOI | 1994 | 11.34 | 1995 | 1999 | |
| Kessler RC, 1994, ARCH GEN PSYCHIAT, V51, P8 | 1994 | 11.2 | 1994 | 1999 | |
| Ferrari AJ, 2013, PLOS MED, V10, P0, DOI 10.1371/journal.pmed.1001547, DOI | 2013 | 10.53 | 2014 | 2018 | |
| Caspi A, 2003, SCIENCE, V301, P386, DOI 10.1126/science.1083968, DOI | 2003 | 10.52 | 2004 | 2008 | |
| Barr J, 2013, CRIT CARE MED, V41, P263, DOI 10.1097/CCM.0b013e3182783b72, DOI | 2013 | 10.1 | 2014 | 2018 | |
| Goldapple K, 2004, ARCH GEN PSYCHIAT, V61, P34, DOI 10.1001/archpsyc.61.1.34, DOI | 2004 | 9.78 | 2005 | 2009 | |

### C. Top 8 References with the Strongest Citation Bursts as Ranked by Beginning Year of Burst (2020 to 2022)

| References | Year | Strength | Begin | End | 2020 - 2022 |
|---|---|---|---|---|---|
| Andrews G, 2018, J ANXIETY DISORD, V55, P70, DOI 10.1016/j.janxdis.2018.01.001, DOI | 2018-JAN | 3.83 | 2020-JUN | 2020-JUN | |
| Qaseem A, 2016, ANN INTERN MED, V165, P125, DOI 10.7326/M15-2175, DOI | 2016-JAN | 3.98 | 2020-JUN | 2020-NOV | |
| Brooks SK, 2020, LANCET, V395, P912 | 2020-JAN | 3.97 | 2021-JAN | 2021-NOV | |
| Xiong JQ, 2020, J AFFECT DISORDERS, V277, P55, DOI 10.1016/j.jad.2020.08.001, DOI | 2020-JAN | 3.4 | 2021-JAN | 2021-JUN | |
| R CORETEAM, 2018, R LANG ENV STAT COMP, V0, P0 | 2018-JAN | 6.03 | 2021-MAY | 2022-JAN | |
| Huang CL, 2021, LANCET, V397, P220, DOI 10.1016/S0140-6736(20)32656-8, DOI | 2021-JAN | 5.48 | 2021-JUN | 2022-MAR | |
| Ouzzani M, 2016, SYST REV-LONDON, V5, P0, DOI 10.1186/s13643-016-0384-4, DOI | 2016-JAN | 3.52 | 2021-JUN | 2021-AUG | |
| Sterne JAC, 2019, BMJ-BRIT MED J, V366, P0, DOI 10.1136/bmj.l4898, DOI | 2019-JAN | 3.57 | 2021-JUN | 2022-JAN | |

### D. Top 8 References with the Strongest Citation Bursts as Ranked by Burst Strength (2020 to 2022)

| References | Year | Strength | Begin | End | 2020 - 2022 |
|---|---|---|---|---|---|
| R CORETEAM, 2018, R LANG ENV STAT COMP, V0, P0 | 2018-JAN | 6.03 | 2021-MAY | 2022-JAN | |
| Huang CL, 2021, LANCET, V397, P220, DOI 10.1016/S0140-6736(20)32656-8, DOI | 2021-JAN | 5.48 | 2021-JUN | 2022-MAR | |
| Qaseem A, 2016, ANN INTERN MED, V165, P125, DOI 10.7326/M15-2175, DOI | 2016-JAN | 3.98 | 2020-JUN | 2020-NOV | |
| Brooks SK, 2020, LANCET, V395, P912 | 2020-JAN | 3.97 | 2021-JAN | 2021-NOV | |
| Andrews G, 2018, J ANXIETY DISORD, V55, P70, DOI 10.1016/j.janxdis.2018.01.001, DOI | 2018-JAN | 3.83 | 2020-JUN | 2020-JUN | |
| Sterne JAC, 2019, BMJ-BRIT MED J, V366, P0, DOI 10.1136/bmj.l4898, DOI | 2019-JAN | 3.57 | 2021-JUN | 2022-JAN | |
| Ouzzani M, 2016, SYST REV-LONDON, V5, P0, DOI 10.1186/s13643-016-0384-4, DOI | 2016-JAN | 3.52 | 2021-JUN | 2021-AUG | |
| Xiong JQ, 2020, J AFFECT DISORDERS, V277, P55, DOI 10.1016/j.jad.2020.08.001, DOI | 2020-JAN | 3.4 | 2021-JAN | 2021-JUN | |

### E. Top 25 Keywords with the Strongest Citation Bursts as Ranked by Beginning Year of Burst (2017 to 2022)

| Keywords | Year | Strength | Begin | End | 2017 - 2022 |
|---|---|---|---|---|---|
| randomizedcontrolled trial | 2017 | 6.11 | 2017 | 2018 | |
| attention deficit/hyperactivity disorder | 2017 | 4.97 | 2017 | 2018 | |
| event related potential | 2017 | 4.59 | 2017 | 2018 | |
| in vivo | 2017 | 4.2 | 2017 | 2018 | |
| dsm iv | 2017 | 4.18 | 2017 | 2018 | |
| pituitary adrenal axi | 2017 | 4.11 | 2017 | 2019 | |
| guided self help | 2017 | 3.82 | 2017 | 2018 | |
| personality trait | 2017 | 3.68 | 2017 | 2019 | |
| self | 2017 | 3.53 | 2017 | 2018 | |
| personality disorder | 2017 | 4.71 | 2018 | 2019 | |
| task | 2017 | 4.47 | 2018 | 2019 | |
| collaborative care | 2017 | 4.35 | 2018 | 2019 | |
| perspective | 2017 | 3.98 | 2018 | 2019 | |
| spatial memory | 2017 | 3.81 | 2018 | 2019 | |
| delirium | 2017 | 3.62 | 2018 | 2019 | |
| subthreshold depression | 2017 | 3.62 | 2018 | 2019 | |
| neural mechanism | 2017 | 3.62 | 2018 | 2019 | |
| functionalconnectivity | 2017 | 5.02 | 2019 | 2020 | |
| satisfaction | 2017 | 4.02 | 2019 | 2020 | |
| support | 2017 | 3.61 | 2019 | 2022 | |
| impulsivity | 2017 | 3.59 | 2019 | 2020 | |
| work | 2017 | 4 | 2020 | 2022 | |
| network | 2017 | 3.91 | 2020 | 2022 | |
| public health | 2017 | 3.72 | 2020 | 2022 | |
| aerobic exercise | 2017 | 3.72 | 2020 | 2022 | |

### F. Top 25 Keywords with the Strongest Citation Bursts as Ranked by Burst Strength (2017 to 2022)

| Keywords | Year | Strength | Begin | End | 2017 - 2022 |
|---|---|---|---|---|---|
| randomizedcontrolled trial | 2017 | 6.11 | 2017 | 2018 | |
| functionalconnectivity | 2017 | 5.02 | 2019 | 2020 | |
| attention deficit/hyperactivity disorder | 2017 | 4.97 | 2017 | 2018 | |
| personality disorder | 2017 | 4.71 | 2018 | 2019 | |
| event related potential | 2017 | 4.59 | 2017 | 2018 | |
| task | 2017 | 4.47 | 2018 | 2019 | |
| collaborative care | 2017 | 4.35 | 2018 | 2019 | |
| in vivo | 2017 | 4.2 | 2017 | 2018 | |
| dsm iv | 2017 | 4.18 | 2017 | 2018 | |
| pituitary adrenal axis | 2017 | 4.11 | 2017 | 2019 | |
| satisfaction | 2017 | 4.02 | 2019 | 2020 | |
| work | 2017 | 4 | 2020 | 2022 | |
| perspective | 2017 | 3.98 | 2018 | 2019 | |
| network | 2017 | 3.91 | 2020 | 2022 | |
| guided self help | 2017 | 3.82 | 2017 | 2018 | |
| spatial memory | 2017 | 3.81 | 2018 | 2019 | |
| public health | 2017 | 3.72 | 2020 | 2022 | |
| aerobic exercise | 2017 | 3.72 | 2020 | 2022 | |
| personality trait | 2017 | 3.68 | 2017 | 2019 | |
| delirium | 2017 | 3.62 | 2018 | 2019 | |
| subthreshold depression | 2017 | 3.62 | 2018 | 2019 | |
| neural mechanism | 2017 | 3.62 | 2018 | 2019 | |
| support | 2017 | 3.61 | 2019 | 2022 | |
| impulsivity | 2017 | 3.59 | 2019 | 2020 | |
| self | 2017 | 3.53 | 2017 | 2018 | |

### G. Top 25 Authors with the Strongest Citation Bursts as Ranked by Beginning Year of Burst (2000 to 2022)

| Authors | Year | Strength | Begin | End | 2000 - 2022 |
|---|---|---|---|---|---|
| HICKIE I | 2000 | 4.65 | 2000 | 2015 | |
| ARNETT P | 2000 | 6.85 | 2001 | 2009 | |
| ANCOLI-ISRAEL S | 2000 | 4.46 | 2001 | 2009 | |
| BENEDICT R | 2000 | 7.41 | 2002 | 2015 | |
| BLEIJENBERG G | 2000 | 5.33 | 2002 | 2012 | |
| COHEN J | 2000 | 4.65 | 2002 | 2011 | |
| STEIN D | 2000 | 4.98 | 2003 | 2014 | |
| GOZAL D | 2000 | 4.47 | 2003 | 2012 | |
| HEATON R | 2000 | 5 | 2004 | 2011 | |
| CHALDER T | 2000 | 4.97 | 2006 | 2011 | |
| BRYANT R | 2000 | 4.29 | 2007 | 2011 | |
| ELY E | 2000 | 4.14 | 2007 | 2018 | |
| ARAYA R | 2000 | 5.67 | 2009 | 2017 | |
| CHRISTENSEN H | 2000 | 4.9 | 2009 | 2018 | |
| COHEN R | 2000 | 5.8 | 2010 | 2014 | |
| VAN STRATEN A | 2000 | 4.28 | 2010 | 2017 | |
| HEDMAN E | 2000 | 8.41 | 2011 | 2016 | |
| LINDEFORS N | 2000 | 7.08 | 2011 | 2016 | |
| LJOTSSON B | 2000 | 5.37 | 2011 | 2018 | |
| RAPEE R | 2000 | 7.46 | 2013 | 2017 | |
| BOTELLA C | 2000 | 4.52 | 2015 | 2019 | |
| LIU J | 2000 | 4.34 | 2015 | 2019 | |
| WANG L | 2000 | 4.81 | 2016 | 2020 | |
| WANG W | 2000 | 5.52 | 2017 | 2022 | |
| ZHOU X | 2000 | 4.45 | 2017 | 2022 | |

### H. Top 25 Authors with the Strongest Citation Bursts as Ranked by Burst Strength (2000 to 2022)

| Authors | Year | Strength | Begin | End | 2000 - 2022 |
|---|---|---|---|---|---|
| HEDMAN E | 2000 | 8.41 | 2011 | 2016 | |
| RAPEE R | 2000 | 7.46 | 2013 | 2017 | |
| BENEDICT R | 2000 | 7.41 | 2002 | 2015 | |
| LINDEFORS N | 2000 | 7.08 | 2011 | 2016 | |
| ARNETT P | 2000 | 6.85 | 2001 | 2009 | |
| COHEN R | 2000 | 5.8 | 2010 | 2014 | |
| ARAYA R | 2000 | 5.67 | 2009 | 2017 | |
| WANG W | 2000 | 5.52 | 2017 | 2022 | |
| LJOTSSON B | 2000 | 5.37 | 2011 | 2018 | |
| BLEIJENBERG G | 2000 | 5.33 | 2002 | 2012 | |
| HEATON R | 2000 | 5 | 2004 | 2011 | |
| STEIN D | 2000 | 4.98 | 2003 | 2014 | |
| CHALDER T | 2000 | 4.97 | 2006 | 2011 | |
| CHRISTENSEN H | 2000 | 4.9 | 2009 | 2018 | |
| WANG L | 2000 | 4.81 | 2016 | 2020 | |
| COHEN J | 2000 | 4.65 | 2002 | 2011 | |
| HICKIE I | 2000 | 4.65 | 2000 | 2015 | |
| BOTELLA C | 2000 | 4.52 | 2015 | 2019 | |
| GOZAL D | 2000 | 4.47 | 2003 | 2012 | |
| ANCOLI-ISRAEL S | 2000 | 4.46 | 2001 | 2009 | |
| ZHOU X | 2000 | 4.45 | 2017 | 2022 | |
| LIU J | 2000 | 4.34 | 2015 | 2019 | |
| BRYANT R | 2000 | 4.29 | 2007 | 2011 | |
| VAN STRATEN A | 2000 | 4.28 | 2010 | 2017 | |
| ELY E | 2000 | 4.14 | 2007 | 2018 | |

### I. Top 25 Institutes with the Strongest Citation Bursts as Ranked by Beginning Year of Burst (2000 to 2022)

| Institutions | Year | Strength | Begin | End | 2000 - 2022 |
|---|---|---|---|---|---|
| Univ Iowa | 2000 | 15.95 | 2000 | 2011 | |
| Cornell Univ | 2000 | 14.99 | 2000 | 2010 | |
| Univ Munich | 2000 | 13.58 | 2000 | 2016 | |
| Duke Univ | 2000 | 12.12 | 2000 | 2011 | |
| Harvard Univ | 2000 | 41.33 | 2001 | 2015 | |
| Univ New S Wales | 2000 | 29.73 | 2001 | 2015 | |
| Univ Texas | 2000 | 21.49 | 2001 | 2007 | |
| Mt Sinai Sch Med | 2000 | 15.71 | 2001 | 2013 | |
| Univ Pittsburgh | 2000 | 15.35 | 2001 | 2007 | |
| Med Univ S Carolina | 2000 | 13.85 | 2001 | 2015 | |
| Inst Psychiat | 2000 | 13.5 | 2001 | 2007 | |
| Univ Florence | 2000 | 9.09 | 2001 | 2014 | |
| SUNY Buffalo | 2000 | 10.07 | 2002 | 2013 | |
| Univ London Imperial Coll Sci Technol & Med | 2000 | 10.85 | 2003 | 2015 | |
| Columbia Univ | 2000 | 12.8 | 2004 | 2009 | |
| Univ Roma La Sapienza | 2000 | 16.13 | 2005 | 2015 | |
| Univ So Calif | 2000 | 11.23 | 2005 | 2015 | |
| Univ Fed Sao Paulo | 2000 | 10.07 | 2007 | 2013 | |
| Charite | 2000 | 10.55 | 2008 | 2016 | |
| Univ Marburg | 2000 | 8.7 | 2010 | 2016 | |
| Univ Paris 06 | 2000 | 13.15 | 2011 | 2015 | |
| CNRS | 2000 | 9.34 | 2012 | 2016 | |
| INSERM | 2000 | 8.97 | 2012 | 2016 | |
| Harvard Med Sch | 2000 | 35.07 | 2017 | 2022 | |
| Univ Southern Calif | 2000 | 8.93 | 2017 | 2022 | |

### J. Top 25 Institutes with the Strongest Citation Bursts as Ranked by Burst Strength (2000 to 2022)

| Institutions | Year | Strength | Begin | End | 2000 - 2022 |
|---|---|---|---|---|---|
| Harvard Univ | 2000 | 41.33 | 2001 | 2015 | |
| Harvard Med Sch | 2000 | 35.07 | 2017 | 2022 | |
| Univ New S Wales | 2000 | 29.73 | 2001 | 2015 | |
| Univ Texas | 2000 | 21.49 | 2001 | 2007 | |
| Univ Roma La Sapienza | 2000 | 16.13 | 2005 | 2015 | |
| Univ Iowa | 2000 | 15.95 | 2000 | 2011 | |
| Mt Sinai Sch Med | 2000 | 15.71 | 2001 | 2013 | |
| Univ Pittsburgh | 2000 | 15.35 | 2001 | 2007 | |
| Cornell Univ | 2000 | 14.99 | 2000 | 2010 | |
| Med Univ S Carolina | 2000 | 13.85 | 2001 | 2015 | |
| Univ Munich | 2000 | 13.58 | 2000 | 2016 | |
| Inst Psychiat | 2000 | 13.5 | 2001 | 2007 | |
| Univ Paris 06 | 2000 | 13.15 | 2011 | 2015 | |
| Columbia Univ | 2000 | 12.8 | 2004 | 2009 | |
| Duke Univ | 2000 | 12.12 | 2000 | 2011 | |
| Univ So Calif | 2000 | 11.23 | 2005 | 2015 | |
| Univ London Imperial Coll Sci Technol & Med | 2000 | 10.85 | 2003 | 2015 | |
| Charite | 2000 | 10.55 | 2008 | 2016 | |
| SUNY Buffalo | 2000 | 10.07 | 2002 | 2013 | |
| Univ Fed Sao Paulo | 2000 | 10.07 | 2007 | 2013 | |
| CNRS | 2000 | 9.34 | 2012 | 2016 | |
| Univ Florence | 2000 | 9.09 | 2001 | 2014 | |
| INSERM | 2000 | 8.97 | 2012 | 2016 | |
| Univ Southern Calif | 2000 | 8.93 | 2017 | 2022 | |
| Univ Marburg | 2000 | 8.7 | 2010 | 2016 | |

### K. Top 13 Countries with the Strongest Citation Bursts as Ranked by Beginning Year of Burst (2000 to 2022)

| Countries | Year | Strength | Begin | End | 2000 - 2022 |
|---|---|---|---|---|---|
| USA | 2000 | 9.43 | 2002 | 2002 | |
| UNITED KINGDOM | 2000 | 7.41 | 2007 | 2007 | |
| NEW ZEALAND | 2000 | 3.57 | 2008 | 2009 | |
| ARGENTINA | 2000 | 3.67 | 2013 | 2014 | |
| COLOMBIA | 2000 | 3.21 | 2017 | 2020 | |
| SAUDI ARABIA | 2000 | 6.94 | 2019 | 2022 | |
| NIGERIA | 2000 | 5.55 | 2019 | 2022 | |
| MEXICO | 2000 | 5.23 | 2019 | 2020 | |
| PEOPLES R CHINA | 2000 | 90 | 2020 | 2022 | |
| IRAN | 2000 | 19.24 | 2020 | 2022 | |
| RUSSIA | 2000 | 5.38 | 2020 | 2020 | |
| PAKISTAN | 2000 | 4.21 | 2020 | 2022 | |
| JORDAN | 2000 | 3.97 | 2020 | 2022 | |

### L. Top 13 Countries with the Strongest Citation Bursts as Ranked by Burst Strength (2000 to 2022)

| Countries | Year | Strength | Begin | End | 2000 - 2022 |
|---|---|---|---|---|---|
| PEOPLES R CHINA | 2000 | 90 | 2020 | 2022 | |
| IRAN | 2000 | 19.24 | 2020 | 2022 | |
| USA | 2000 | 9.43 | 2002 | 2002 | |
| UNITED KINGDOM | 2000 | 7.41 | 2007 | 2007 | |
| SAUDI ARABIA | 2000 | 6.94 | 2019 | 2022 | |
| NIGERIA | 2000 | 5.55 | 2019 | 2022 | |
| RUSSIA | 2000 | 5.38 | 2020 | 2020 | |
| MEXICO | 2000 | 5.23 | 2019 | 2020 | |
| PAKISTAN | 2000 | 4.21 | 2020 | 2022 | |
| JORDAN | 2000 | 3.97 | 2020 | 2022 | |
| ARGENTINA | 2000 | 3.67 | 2013 | 2014 | |
| NEW ZEALAND | 2000 | 3.57 | 2008 | 2009 | |
| COLOMBIA | 2000 | 3.21 | 2017 | 2020 | |

**Supplementary Table 3**. Detailed information on the clusters in the co-occurrence keywords network and co-authorship network (2017-2022 network and 2000-2022 network)

| **A. Information of the clusters in the co-occurrence keywords network (2017 to 2022)** | | | | | |
|---|---|---|---|---|---|
| **Cluster #** | **Cluster Size** | **Silhouette Score** | **Mean Year** | **Cluster Label** | **Top Five Extracted Terms (Log-likelihood ratio; p-value)** |
| 0 | 166 | 0.694 | 2018 | Brain Structure & Function | hippocampus (135.16, 1.0E-4); prefrontal cortex (107.66, 1.0E-4); synaptic plasticity (95.34, 1.0E-4); functional connectivity (70.1, 1.0E-4); neurogenesis (69.61, 1.0E-4) |
| 1 | 148 | 0.663 | 2018 | Emotion Regulation | emotion regulation (118.11, 1.0E-4); social anxiety (81.32, 1.0E-4); eating disorders (61.18, 1.0E-4); virtual reality (53.38, 1.0E-4); rumination (46.84, 1.0E-4) |
| 2 | 130 | 0.674 | 2018 | Cognition/Treatment | cognitive behavioral therapy (116.28, 1.0E-4); psychotherapy (75.52, 1.0E-4); cognition (68.06, 1.0E-4); cognitive impairment (66.83, 1.0E-4); mental health (66.56, 1.0E-4) |
| 3 | 112 | 0.701 | 2018 | Frailty | frailty (119.56, 1.0E-4); quality of life (96.29, 1.0E-4); cognitive impairment (90.41, 1.0E-4); elderly (63.81, 1.0E-4); older adults (59.84, 1.0E-4) |
| 4 | 59 | 0.77 | 2018 | Insomnia | insomnia (98.29, 1.0E-4); sleep (70.26, 1.0E-4); sleep disorders (52.46, 1.0E-4); sleep quality (43.67, 1.0E-4); bariatric surgery (36.48, 1.0E-4) |
| 5 | 50 | 0.747 | 2018 | Comorbidities | cognitive dysfunction (78.27, 1.0E-4); multiple sclerosis (49.56, 1.0E-4); amyotrophic lateral sclerosis (34.13, 1.0E-4); hepatitis c (23.14, 1.0E-4); concussion (17.35, 1.0E-4) |

| B. Information of the largest clusters in the co-authorship network (2000 to 2022) | | | | | |
|---|---|---|---|---|---|
| **Cluster #** | **Cluster Size** | **Silhouette Score** | **Mean Year** | **Cluster Label** | **Top Five Extracted Terms (Log-likelihood ratio; p-value)** |
| 0 | 213 | 0.956 | 2017 | Hippocampus | hippocampus (21.99, 1.0E-4); learning and memory (14.8, 0.001); neuroinflammation (14.5, 0.001); internet (14.09, 0.001); cognitive impairment (13.74, 0.001) |
| 1 | 108 | 0.955 | 2013 | Internet | internet (33.35, 1.0E-4); cognitive behavior therapy (21.5, 1.0E-4); internet-based treatment (18.42, 1.0E-4); cost-effectiveness (18.42, 1.0E-4); hippocampus (17.65, 1.0E-4) |
| 2 | 60 | 0.919 | 2014 | Sluggish Cognitive Tempo | sluggish cognitive tempo (16.21, 1.0E-4); brain (12.96, 0.001); cancer (9.71, 0.005); chemotherapy (9.71, 0.005); aged (9.71, 0.005) |
| 3 | 26 | 0.989 | 2012 | Neuroimaging | neuroimaging (17.21, 1.0E-4); behavioral inhibition (15.12, 0.001); reward (15.12, 0.001); conditioning (10.07, 0.005); dsm-v (10.07, 0.005) |
| 6 | 14 | 0.997 | 2012 | Cognitive Arousal | cognitive arousal (18.46, 1.0E-4); perinatal (9.18, 0.005); improvement (9.18, 0.005); rumination (9.18, 0.005); cognitive-behavioral therapy for insomnia (9.18, 0.005) |
| 7 | 12 | 0.997 | 2011 | Heart Failure | heart failure (46.9, 1.0E-4); cardiovascular disease (9.18, 0.005); cerebral blood flow velocity (9.18, 0.005); step count (9.18, 0.005); psychosocial outcomes (9.18, 0.005) |
| 8 | 11 | 1 | 2012 | Delirium | delirium (34.9, 1.0E-4); critical care (17.3, 1.0E-4); critical illness (17.3, 1.0E-4); neuroimaging (9.06, 0.005); survivors (8.62, 0.005) |
| 9 | 10 | 0.997 | 2009 | Time Variation | time variation (10.43, 0.005); adolescent depression (10.43, 0.005); treatment-resistant (10.43, 0.005); selective serotonin reuptake inhibitor (10.43, 0.005); abuse (10.43, 0.005) |
| 13 | 7 | 0.998 | 2012 | Post-traumatic Stress Disorder | ptsd (9.5, 0.005); record linkage (8.48, 0.005); medial prefrontal cortex (8.48, 0.005); overgenerality (8.48, 0.005); dorsomedial prefrontal cortex (8.48, 0.005) |
| 15 | 6 | 1 | 2016 | Anxiety Sensitivity | anxiety sensitivity (27.27, 1.0E-4); cannabis (9, 0.005); substance use motives (9, 0.005); substance use disorder (9, 0.005); measurement invariance (9, 0.005) |
| 17 | 6 | 0.999 | 2003 | Violence | violence (11.91, 0.001); latinos (11.91, 0.001); trauma (6.53, 0.05); treatment (6.2, 0.05); posttraumatic stress disorder (5.67, 0.05) |
| 22 | 5 | 0.999 | 2018 | Longitudinal Study | longitudinal study (17.92, 1.0E-4); oldest old (17.92, 1.0E-4); health care utilization (8.92, 0.005); loss experiences (8.92, 0.005); old age (8.92, 0.005) |
| 24 | 5 | 0.991 | 2014 | Psychiatry | psychiatry (NaN, 1.0); youth (NaN, 1.0); cognitive-behavioral therapy (cbt) (NaN, 1.0); acceptability (NaN, 1.0); metabolomics (NaN, 1.0) |
| 55 | 3 | 0.994 | 2008 | Behavioural Disinhibition | behavioural disinhibition (11.91, 0.001); visual discrimination (11.91, 0.001); glucocorticoid receptor (11.91, 0.001); gr-antisense transgene (11.91, 0.001); signal detection theory (9.14, 0.005) |

| | | | | | |
|---|---|---|---|---|---|
| 57 | 3 | 0.995 | 2017 | Stress and Adjustment Disorders | stress and adjustment disorders (11.29, 0.001); telomere length (11.29, 0.001); cytokines (11.29, 0.001); biomarkers (11.29, 0.001); inflammation (4.48, 0.05) |
| 99 | 2 | 0.992 | 2016 | Multiple Sclerosis | multiple sclerosis (11.49, 0.001); quantitative mri (10.81, 0.005); pseudo-bulbar affect (10.81, 0.005); pathological laughing and crying (10.81, 0.005); demyelination (10.81, 0.005) |
| 127 | 2 | 1 | 2014 | Measurement | measurement (10.43, 0.005); neuro-qol (10.43, 0.005); adult (10.43, 0.005); excessive exercising (10.43, 0.005); validation (10.43, 0.005) |
| 128 | 2 | 0.999 | 2003 | Post-traumatic Stress Disorder | posttraumatic stress disorder (26.81, 1.0E-4); childhood traumatic grief (19.3, 1.0E-4); child sexual abuse (19.3, 1.0E-4); trauma- and grief-focused interventions (9.59, 0.005); childhood trauma (9.59, 0.005) |

**Supplementary Table 4**. Top references, keywords, institutions, and countries ranked by degree of centrality and counts/citation counts (2017-2022 network and 2000-2022 network)

| A. Top References (2020-2022) | | | | | |
|---|---|---|---|---|---|
| Ranked by Degree of Centrality | | | Ranked by Citation Count | | |
| | References | Degree of Centrality | | References | Citation Count |
| 1 | Carlbring P, 2018, COGN BEHAV THERAPY, 47, 1 | 77 | 1 | Carlbring P, 2018, COGN BEHAV THERAPY, 47, 1 | 41 |
| 2 | Andrews G, 2018, J ANXIETY DISORD, 55, 70 | 56 | 2 | Andrews G, 2018, J ANXIETY DISORD, 55, 70 | 28 |
| 3 | Spijkerman MPJ, 2016, CLIN PSYCHOL REV, 45, 102 | 33 | 3 | Riemann D, 2017, J SLEEP RES, 26, 675 | 20 |
| 4 | Huang CL, 2021, LANCET, 397, 220 | 32 | 4 | Qaseem A, 2016, ANN INTERN MED, 165, 125 | 20 |
| 5 | Andersson G, 2018, INTERNET INTERV, 12, 181 | 26 | 5 | James SL, 2018, LANCET, 392, 1789 | 18 |
| 6 | Paganini S, 2018, J AFFECT DISORDERS, 225, 733 | 26 | 6 | Huang CL, 2021, LANCET, 397, 220 | 17 |
| 7 | Qaseem A, 2016, ANN INTERN MED, 165, 125 | 25 | 7 | R Core Team, 2018, R LANG ENV STAT COMP, 0, 0 | 16 |
| 8 | Titov N, 2018, INTERNET INTERV, 13, 108 | 25 | 8 | Brooks SK, 2020, LANCET, 395, 912 | 13 |
| 9 | James SL, 2018, LANCET, 392, 1789 | 24 | 9 | Hayes AF, 2018, INTRO MEDIATION MODE, 2nd, 0 | 13 |

| B. Top Keywords (2017-2022) | | | |
|---|---|---|---|
| | Keywords | Year | Count |
| 1 | Depression | 2017 | 1,215 |
| 2 | Disorder | 2017 | 955 |
| 3 | Anxiety | 2017 | 699 |
| 4 | Cognitive behavioral therapy | 2017 | 667 |
| 5 | Quality of life | 2017 | 486 |
| 6 | Prevalence | 2017 | 478 |
| 7 | Meta-analysis | 2017 | 459 |
| 8 | Symptom | 2017 | 446 |
| 9 | Cognitive impairment | 2017 | 368 |
| 10 | Validation | 2017 | 345 |
| 11 | Health | 2017 | 338 |
| 12 | Mental health | 2017 | 329 |
| 13 | Older adult | 2017 | 325 |
| 14 | Impairment | 2017 | 311 |
| 15 | Association | 2017 | 273 |

### C. Top Institutions (2000-2022)

| Ranked by Degree of Centrality | | | Ranked by Citation Count | | |
|---|---|---|---|---|---|
| | Institution | Degree of Centrality | | Institution | Citation Count |
| 1 | Harvard University | 117 | 1 | King's College London | 414 |
| 2 | Emory University | 106 | 2 | University of Toronto | 357 |
| 3 | University of North Carolina | 96 | 3 | University of Pittsburgh | 321 |
| 4 | University of California, Los Angeles | 89 | 4 | Karolinska Institute | 292 |
| 5 | Columbia University | 88 | 5 | University of California, Los Angeles | 280 |
| 6 | Duke University | 85 | 6 | Stanford University | 279 |
| 7 | Yale University | 84 | 7 | Harvard University | 273 |
| 8 | University of Pittsburgh | 83 | 8 | University College London | 269 |
| 9 | Washington University | 82 | 9 | University of California, San Diego | 268 |
| 10 | University of California, San Diego | 81 | 10 | University of Oxford | 266 |

### D. Top Countries (2000-2022)

| Ranked by Degree of Centrality | | | Ranked by Citation Count | | |
|---|---|---|---|---|---|
| | Country | Degree of Centrality | | Country | Citation Count |
| 1 | United Kingdom | 85 | 1 | United States of America | 7497 |
| 2 | United States of America | 79 | 2 | United Kingdom | 2266 |
| 3 | Germany | 68 | 3 | Germany | 1903 |
| 4 | Italy | 65 | 4 | Australia | 1484 |
| 5 | Netherlands | 64 | 5 | People's Republic of China | 1390 |
| 6 | Spain | 63 | 6 | Canada | 1371 |
| 7 | Canada | 60 | 7 | Netherlands | 1221 |
| 8 | France | 58 | 8 | Italy | 1121 |
| 9 | Switzerland | 53 | 9 | France | 875 |
| 10 | Australia | 51 | 10 | Spain | 795 |
